# Site-specific immune remodeling underlies resistance to dual immune checkpoint blockade in metastatic colorectal cancer

**DOI:** 10.64898/2026.10.23.26365904

**Authors:** Iosune Baraibar, Samuel Morabito, Gerard Deuner-Cos, Ginevra Caratù, Juan Luis Melero, Miren Berasategi, Marta Soto, Marta Grzelak, Paula Nieto, Davide Maspero, Francesc Salvà, Nadia Saoudi, Javier Ros, Raquel Pérez-López, Alena Gros, Guillermo Villacampa, Nuria Roson, Javier Carmona, Ariadna García, Arthur Lambert, Nadia Martinez-Martin, Patricia Casbas-Hernandez, Jorge Zeron-Medina, Josep Tabernero, Asaf Rotem, Juan C. Nieto, Elena Élez, Holger Heyn

## Abstract

Immune checkpoint inhibitors (ICIs) have limited efficacy in metastatic colorectal cancer (mCRC) with proficient mismatch repair and microsatellite stability (pMMR/MSS), with liver metastases emerging as a dominant determinant of resistance. Whether this reflects a failure to activate antitumor immunity or a failure to direct it remains unclear. We investigated this in the phase II SERPENTINE trial of dual PD-L1 and CTLA-4 blockade by integrating paired pre-treatment and on-treatment single-cell transcriptomic and T-cell receptor (TCR) profiling (388,196 cells; 95,617 clonotypes) metastatic biopsies with deep TCR sequencing of matched peripheral blood (57,902,418 clonotypes). In MSS mCRC, clinical benefit was restricted to patients without liver metastases, whereas all patients with liver metastases progressed. Dual ICI induced systemic T-cell priming regardless of metastatic site, mobilizing therapy-expanded clones that trafficked to both lung and liver lesions, but the fate of this response was determined locally. In lung metastases, therapy-specific clones focused into tissue-resident, proliferative, tumor-reactive CD8+ T-cell states. In liver metastases, the same activation was dispersed: regulatory T cells and suppressive myeloid populations expanded, TCR repertoires diversified, and exhaustion-associated T-cell gene networks fragmented. Cross-lineage co-expression analysis further revealed that tumor proliferation increased in concert with cytotoxic CD8+ T-cell programs, indicating effector engagement that fails to restrain tumor growth. These findings show that resistance to ICI in liver metastases arises from tissue-specific misdirection of immune activation away from productive antitumor immunity, identifying multicellular coordination of the immune response in the metastatic microenvironment as a determinant of therapeutic efficacy.

## Introduction

Colorectal cancer (CRC) is the third most common primary tumor worldwide and the second in terms of mortality^1^. In unresectable metastatic CRC (mCRC), therapeutic decision-making is currently guided by the tumor’s molecular profile and the individual patient’s clinical condition and preferences. In CRC, microsatellite instability (MSI) status and deficient mismatch repair (dMMR) are currently the primary biomarkers for predicting immunotherapeutic response^2^. This sensitivity to immune checkpoint inhibition (ICI) is attributed to the accumulation of DNA replication errors resulting from the loss of function of one or more DNA mismatch repair enzymes (MSH2, MSH6, MLH1, and PMS2). This defect leads to a widespread genetic instability in short nucleotide repeats, so called microsatellites, giving rise to a hypermutated phenotype^3^. As a consequence, these tumors exhibit high tumor mutational burden (TMB) and an inflamed tumor microenvironment (TME) enriched in tumor infiltrating lymphocytes (TIL), particularly memory cells and cytotoxic T lymphocytes^4^.

In contrast to dMMR/MSI (MSI) CRC, tumors with proficient MMR (pMMR) and microsatellite stability (MSS) are typically poorly immunogenic. Their low tumor mutation burden (TMB) and sparse neoantigen load result in an immune-excluded or ‘desert’ TME characterized by absent or inactive TILs, ultimately leading to resistance to ICIs^5,6^. Efforts to identify biomarkers of immunotherapy response in pMMR/MSS have revealed the specific metastatic site as a critical determinant of resistance. The presence of liver metastases is consistently associated with diminished efficacy of immunotherapy-based combinations in pMMR/MSS mCRC patients^7–12^. This clinical resistance is largely attributed to the liver’s immunosuppressive milieu, characterized by hepatic siphoning of T cells that leads to systemic reduction in CD8+ T cells^13,14^. The high percentage of patients with refractory pMMR/MSS mCRC harboring liver metastases represents a major roadblock to the success of current immunotherapy-based strategies. Yet, the precise molecular mechanisms driving this phenomenon remain poorly understood, partly due to the limited integration of mechanistic analyses in prospective clinical research studies.

Prior CRC immunotherapy studies have largely relied on bulk or cross-sectional profiling of primary or resectable tumors, limiting insight into dynamic immune responses and coordinated adaptation of tumor-resident and systemic compartments. Single-cell RNA sequencing (scRNA-seq) has fundamentally redefined our understanding of tumor immunity, shifting the focus from bulk immune cell abundance towards cell states, spatial organization, and clonal architecture^15–18^. Tumor-reactive CD8+ T cells reside within structured niches shaped by antigen-presenting cells, stromal elements, and metabolic cues. Within these niches, dendritic cells regulate T-cell priming and differentiation, with immune checkpoints such as PD-1 and CTLA-4 playing context-dependent roles in sustaining cytotoxic high-avidity populations^19^. These insights indicate that effective anti-tumor immune responses and clinical benefit from ICIs depend on the presence and preservation of tumor-reactive precursor pools and intact antigen-presentation niches^20^. In metastatic MSS CRC, these conditions are often compromised by immune exclusion and stromal remodeling^21^.

To address these questions in a clinical setting, we designed the SERPENTINE trial (NCT06680739) to interrogate immune remodeling in patients with refractory metastatic pMMR/MSS and dMMR/MSI CRC and endometrial cancer treated with combined PD-L1 and CTLA-4 blockade. Here we present the results focused on patients with CRC. By integrating paired metastatic tumor biopsies and peripheral blood sampling at baseline and on treatment, SERPENTINE enables longitudinal, single-cell–resolved analysis of metastatic tumor–immune ecosystems. By directly contrasting how dual checkpoint blockade reshapes cellular states and tumor-reactive T-cell clonal architectures in liver and lung metastases, this design allowed us to ask whether resistance in this typically immune-refractory disease reflects a failure to activate antitumor immunity or a failure to direct it.

## Results

### Patients and treatment

From 33 patients assessed for eligibility 30 were enrolled in the multicohort SERPENTINE trial (**Figures 1A**, **1B and S1A**). Twenty-four patients were assigned to the pMMR/MSS cohort, and received treatment with durvalumab and tremelimumab, while 6 patients were included in the dMMR/MSI cohort and randomized to durvalumab monotherapy or combination with durvalumab and tremelimumab. From the 24 patients with MSS/pMMR CRC, 6 presented liver metastases (25%) and 18 showed no active liver metastases (75%). Ten out of 18 patients (55.5%) without liver metastases underwent computed tomography-scan guided biopsy of lung metastases, while ultrasound-guided liver biopsies were obtained in the 6 patients with liver involvement (100%, **Figure S1A**). Among patients without liver metastases, objective response rate (ORR) was 12% and disease control rate (DCR) was 41%, while all patients with liver disease presented progression disease (PD) as the best radiological response (**Figure S1B**). The 6-months PFS rate was 17% and 0% for patients without and with liver metastases, respectively (HR = 0.51; 95% 0.18 - 1.42) (**Figures S1C and S1D**). Consequently, the absence of liver metastases and peritoneal disease was associated with improved PFS (HR 0.25, 95%CI 0.09-0.76, p = 0.01) (**Figure S1F**). From the dMMR/MSI cohort, two patients presented liver disease that was biopsied, following ultrasound-guided obtention. All patients in the dMMR/MSI mCRC cohort experienced clinical benefit. Two patients presented complete response (CR, 33%), 1 patient had partial response (16%), and 3 patients presented prolonged stable disease (50%), throwing a DCR of 100% (**Figure S1G**). Median PFS for the pMMR/MSI was not reached.

**Figure 1.**
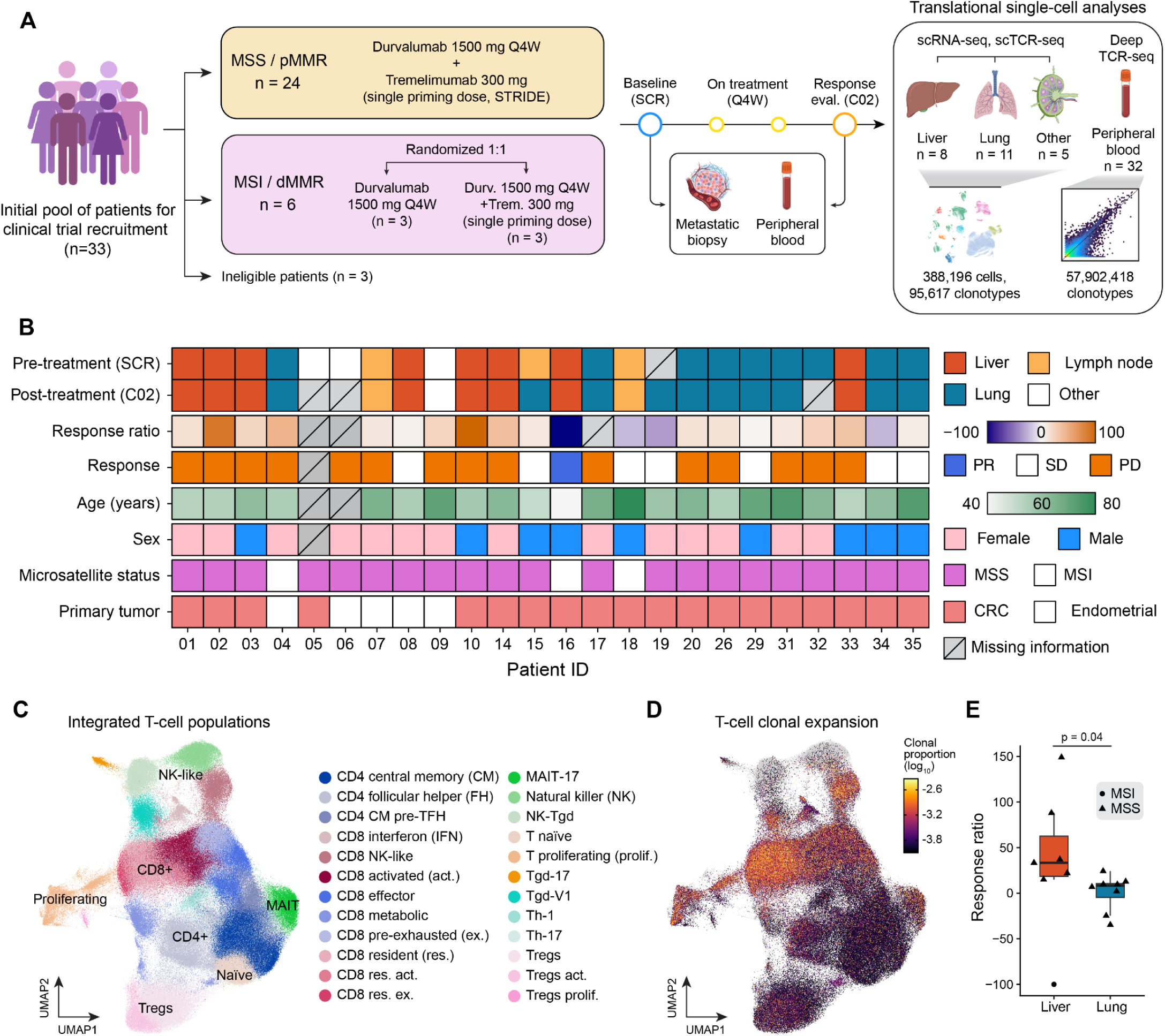
Longitudinal single-cell profiling of metastatic CRC under dual ICI. **A**, Clinical trial design of the SERPENTINE study, and summary of translational single-cell experiments. Patients received dual immune checkpoint inhibition (ICI) with durvalumab and tremelimumab, with metastatic biopsies and peripheral blood collected at baseline (SCR) and at response evaluation (C02). We performed paired single-cell RNA sequencing (scRNA-seq) and single-cell T-cell receptor sequencing (scTCR-seq) in the metastatic tumor microenvironments (TMEs), and we performed deep TCR-seq (Omniscope OS-TCR) in peripheral blood. Illustrations were created with Biorender.com. **B**, Characteristics of patients sequenced in this study (*n*=84 sequencing libraries, *n*=24 patients). Response indicates best radiological response (PR, partial response; SD, stable disease; PD, progressive disease). **C**, Uniform manifold approximation and projection (UMAP) of the T-cell lineage integrated between the SERPENTINE cohort (n=238,458 cells) and primary CRC TMEs from an external dataset^25^ (n=119,642 cells). Datasets were integrated with Harmony^43^ and CellANOVA^61^. Each point represents one cell, colored by their cell-type annotations derived from Leiden clustering^62^ analysis. **D**, T-cell UMAP plot as in (**C**), colored by clonal proportion, log₁₀-transformed (with an added ε = 1×10⁻⁴) to accommodate the skewed distribution of scTCR-seq data. **E**, Distributions of tumor response ratios between patients with liver metastases versus lung metastases (in the absence of liver involvement). P value from a two-sided Wilcoxon rank-sum test is shown. For box-and-whisker plots, box boundaries and lines correspond to the interquartile range (IQR) and median, respectively. Whiskers extend to the lowest or highest data points that are no further from 1.5 times the IQR from the box boundaries.

### Site-specific immune coordination drives divergent clinical responses to ICI

To map the systemic cellular and molecular changes within the mCRC TME induced by dual ICI, we performed paired scRNA-seq and scTCR-seq (10X Genomics Chromium 5’ assay) on fresh longitudinal metastatic tumor biopsies (**Figure 1A**). After quality control filtering, we retained 388,196 cells from a cohort of 24 patients and 84 sequencing scRNA-seq libraries (mean cells/library = 4,621; **Figures 1B and S2**; **Tables S1 and S2**). Unsupervised clustering and marker gene detection identified the major cell compartments of the TME, including T and natural killer (NK) cells (n=238,458), tumor cells (n=84,431), B/plasma cells (n=21,075), myeloid cells (n=29,432), stromal cells (n=12,617), and mast cells (n=2,183; **Tables S3-S6**). To contextualize the metastatic cellular landscape relative to primary disease, we used CellANOVA^22^ to integrate our dataset with an external longitudinal scRNA-seq and scTCR-seq dataset of primary CRC TMEs undergoing a neoadjuvant PD-1 blockade^16^ (58 sequencing libraries from 22 patients). Initial global clustering analysis included all patients enrolled in the trial, while downstream comparative and clinical analyses included only those with CRC. Within the T-cell lineage, we identified 24 distinct cell populations encompassing a broad phenotypic spectrum, including naïve, memory, and various states of exhaustion and activation (**Figure 1C**). Leveraging paired scTCR-seq data, we quantified clonal expansion at single-cell resolution, revealing clonal dynamics of specific T-cell phenotypes and in response to treatment (**Figure 1D**).

In this cohort, we found distinct clinical responses by metastasis sites. Patients with lung metastases in the absence of liver involvement had a significantly more favorable tumor response ratio in comparison to those with liver metastases, suggesting that site-specific TME factors locally constrain the efficacy of dual ICI (two-sided Wilcoxon rank-sum test p=0.04, **Figure 1E**). Hypothesizing that these clinical outcomes reflect divergent remodeling of the immune niche, we employed a neighborhood-based differential abundance test (negative binomial generalized linear model, Milo^23^) to compare post-versus pre-treatment immune landscapes across the T-cells, B/plasma, and myeloid compartments (**Figures S3-S6**). Within the myeloid lineage, the liver was characterized by an expansion of *CCL18*+ resident macrophages and Mono3^24^ (*RETN*, *CSF3R*, *TSPO*) subsets of monocytes, while the lung TME exhibited a phenotypic shift from inflammatory TAMs towards a *PDL1*+ TAMs (**Figure S3**). Our analysis also revealed metastatic site-specific patterns of T-cell remodeling in response to immunotherapy, mirroring the clinical results. In liver metastases, dual ICI induced a robust expansion of cellular neighborhoods mapping to NK, NK-Tγδ, Th-1, and Treg phenotypes, while simultaneously leading to a contraction of CD8+ resident and exhausted populations (**Figures 2A and S4**). In contrast, the lung TME exhibited an enrichment of proliferating and resident exhausted CD8+ T-cells, alongside the depletion of interferon-responsive, central memory, and naïve T-cell pools (**Figures 2B and S5**). While the lung TME exhibited the expansion of tumor-reactive exhausted T-cell pools, the liver TME was characterized by a reciprocal loss of these populations and a concomitant enrichment of Tregs post-treatment. This suggests that dual-ICI in the liver may inadvertently reinforce an immunosuppressive state by favoring regulatory recruitment over effector T-cell persistence.

**Figure 2.**
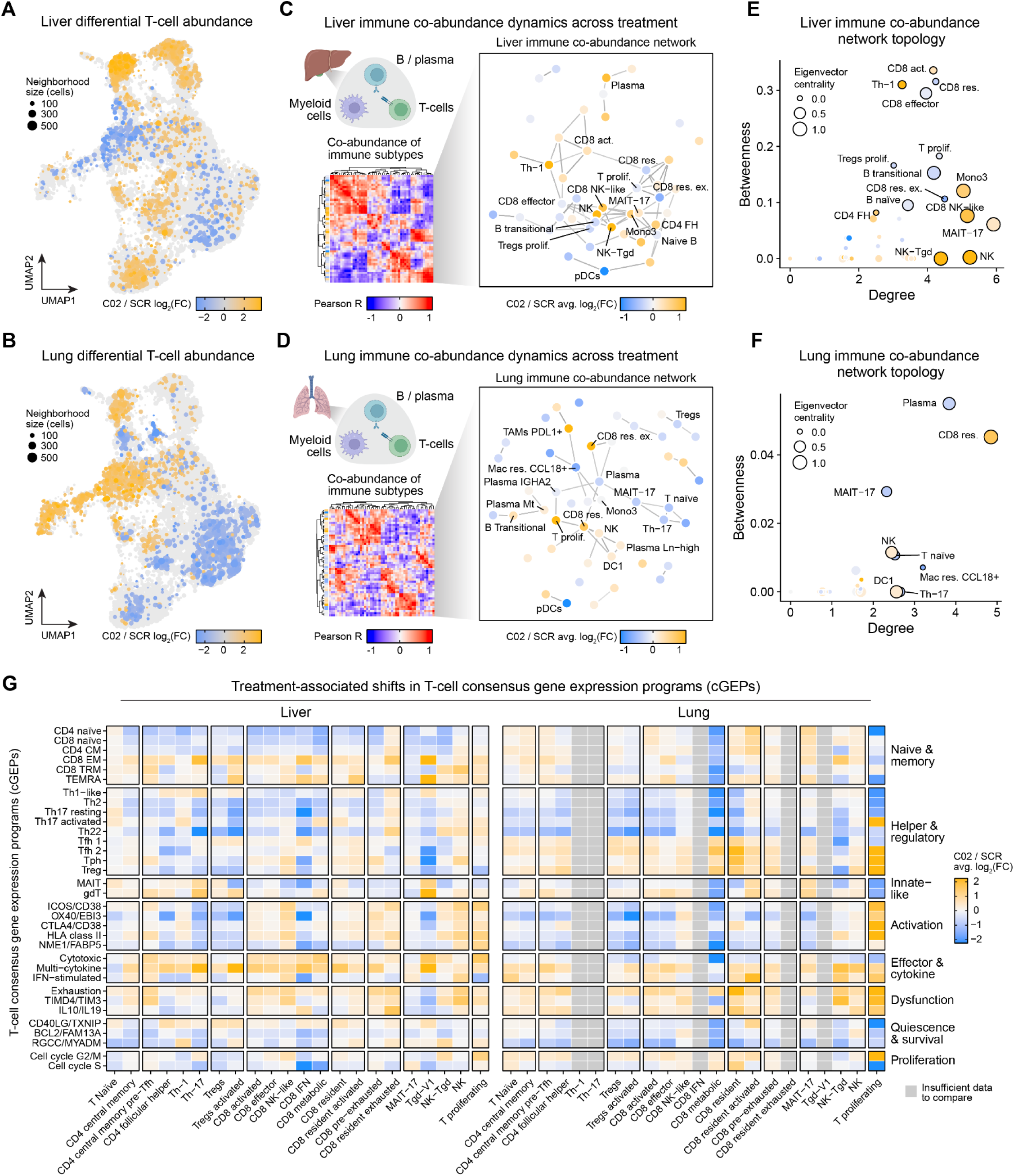
Dual ICI drives divergent immune remodeling in liver and lung metastase. **A-B**, Differential T-cell abundance analysis comparing post-treatment (C02) vs. pre-treatment (SCR) within the liver (**A**) and lung (**B**) TMEs. Points represent cellular neighborhoods from Milo^23^, colored by the effect size of the comparison as log2(fold-change) and point size is scaled by the number of cells per neighborhood. Orange color indicates enrichment of cell populations post-treatment, and blue color indicates enrichment pre-treatment. **C-D**, Systems-level immune co-abundance dynamics across T-cell, B/plasma, and myeloid compartments in liver (C) and lung (D) TMEs. We calculated the change post- vs. pre-treatment (Δ C02 − SCR) of normalized immune subtype abundances (centered log ratios) in each patient, followed by pairwise Pearson correlations across all immune subtypes (heatmaps). Hierarchical clustering of these correlations shows groups of immune cells that respond similarly to treatment. In the network plots, nodes represent immune populations, and edges represent similarity in treatment-induced abundance changes (Pearson’s R > 0.6 and permutation P < 0.05, n = 1,000 permutations). Nodes are colored by the average effect size of the Milo differential abundance test. Selected immune populations are labeled. **E-F**, Topological analysis of the liver (**E**) and lung (**F**) immune co-abundance networks. Node degree (number of direct connections) and betweenness centrality (how often a node bridges otherwise unconnected parts of the network) are compared for each immune population, and node size represents eigenvector centrality (connection to other highly connected nodes). **G**, Heatmap showing treatment-induced shifts in 33 T-cell consensus gene expression programs (cGEPs)^20^ across cell populations, split by liver and lung metastases. Color indicates the average log₂(fold-change) post- vs. pre-treatment (C02/SCR), estimated with a paired limma^46^ model. cGEPs (y-axis) and cell populations (x-axis) are grouped by functional similarity.

We hypothesized that site-specific shifts in immune neighborhoods act as part of a larger coordinated multicellular response to therapy. To identify these synchronized dynamics, we performed a cross-lineage immune co-abundance network analysis, comparing the treatment-induced changes (Δ centered log-ratio, post/pre-treatment) in abundance of each immune subtype across the T-cell, B/plasma, and myeloid compartments (**Figures 2C-D and S7-S9**; see **Methods**). Connections in these networks represent shared dynamics in immune cell abundance in response to treatment. This analysis revealed distinct topological “hubs” and patterns of immune co-regulation in the liver and lung TMEs. In the liver, the network exhibited high global density, where Th-1 and various CD8+ populations (activated, resident, and effector) displayed high betweenness and degree centrality (**Figure 2E**). The central topological position of these effector-phenotype cells within a dense, regulatory-rich network suggests that they are functionally sequestered and constrained by a wide array of non-productive, immuno-suppressive interactions. While NK and NK-Tγδ populations also showed high degree centrality, their low betweenness implicates them as localized hubs, likely driving site-specific innate responses decoupled from a productive systemic response. In contrast, the lung network is notably sparser, centered on a high-centrality axis of CD8+ resident T-cells and plasma cells (**Figure 2F**). The immune co-abundance network in the external dataset of primary tumors following neoadjuvant PD-1 blockade^25^ was also characterized by high connectivity, but the network was topologically centered on CD8+ pre-exhausted and CD4+ follicular helpers (**Figure S10**). This suggests that while the lung TME allows for a well-partitioned effector response, the TME architecture of the liver network promotes a cascade of unintended multicellular feedback. Consequently, the liver’s resistance appears to be a systemic property of its immune coordination network topology, where therapeutic signals are disrupted and dissipated across a highly connected and inhibitory immune landscape.

We next inspected transcriptional changes in a set of T-cell consensus gene expression programs (cGEPs)^20^ which represent different axes of T-cell biology such as programs related to naive/memory, helper/regulatory, innate-like, activation, effector/cytokine, dysfunction, quiescence/survival, and proliferation (**Figure 2G**). In the liver, treatment was broadly associated with reduced naive/memory programs and increased effector/cytokine programs across most populations. In the lung, the shift favored exhaustion programs within CD8+ and resident populations, together with an increase in follicular and peripheral helper programs. This is consistent with the lung-specific expansion of tumor-reactive exhausted T-cell pools we observed by differential abundance. These cGEPs shifts corroborate the site-specific remodeling in the composition of the T-cell compartment at the transcriptomic level, yet reference-derived programs focus on signals already catalogued in existing atlases, leaving programs unique to metastatic disease under dual ICI uncharted.

### Immunotherapy triggers transcriptional network fragmentation in liver metastases

To define the intrinsic gene expression programs governing metastatic T-cell dynamics, we performed high-dimensional weighted gene co-expression network analysis (hdWGCNA^26,27^), constructing separate co-expression networks for each tissue and timepoint. This identified 95 gene co-expression modules across metastatic sites and treatments (**Figures 3A**, **S11-S18**; **Tables S7 and S8**; see **Methods**). To assign biological functions to these modules, we developed **llegir** (**LL**M-**e**nabled **g**ene module **i**nterpretations in **R**; see **Methods**), which orchestrates a suite of downstream analyses to assemble an evidence dossier for each module and then synthesizes it into a biological interpretation using a large language model. Applying llegir to all 95 modules recovered programs related to T-cell activation, antigen presentation, cytotoxicity, metabolic activity, and exhaustion, among others. To compare these modules across contexts, we built a module-module similarity network based on shared expression profiles and member genes, revealing the broader transcriptional architecture of the T-cell compartment in the TME under dual ICI (**Figure 3B**). This similarity network also served as an orthogonal consistency check on the llegir annotations: modules with similar gene sets and expression patterns received thematically related labels. We further contextualized the modules by correlating their expression with T-cell cGEPs. For example, a module related to MHC class II antigen presentation (lung M13, pre-treatment) correlated with the exhaustion cGEP, whereas a module related to NK cell activation (liver M13, pre-treatment) correlated with the cytotoxic cGEP (**Figures 3C and 3D, S19-S21**).

**Figure 3.**
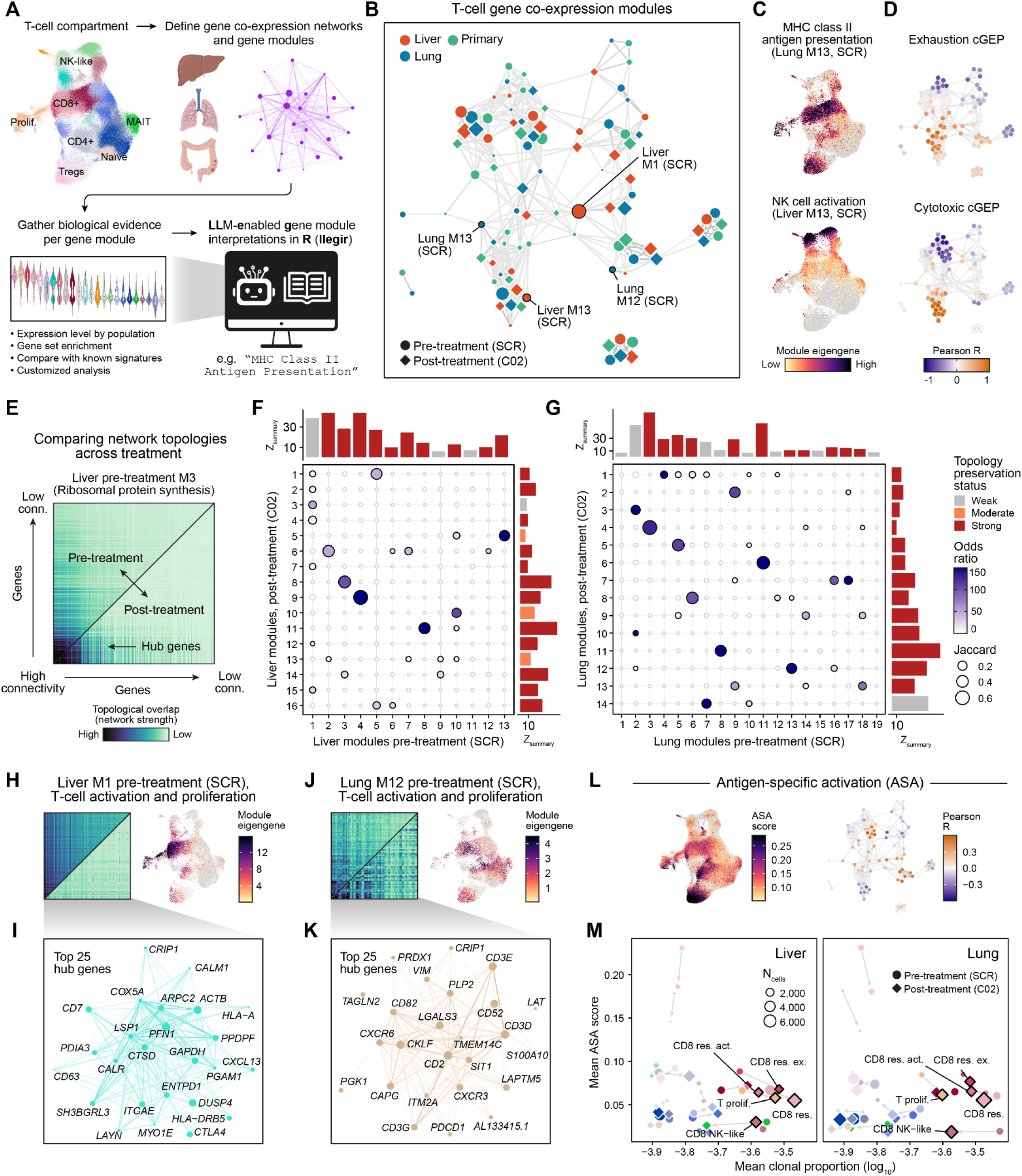
Immunotherapy triggers transcriptional network fragmentation in liver metastases. **A**, Overview of the computational strategy. The T-cell compartment (UMAP, as in Figure 1C) was stratified by tissue (liver, lung, and primary tumor) and by pre- or post-treatment. For each subset, we constructed a gene co-expression network using high-dimensional weighted gene co-expression network analysis (hdWGCNA)^27^ and grouped genes into modules. Biological labels were attributed to these modules by synthesizing a consistent set of evidence per module via **llegir** (**LL**M-**e**nabled **g**ene module **i**nterpretations in **R**. **B**, Network of T-cell gene co-expression modules identified by hdWGCNA. Nodes represent modules, colored by tissue and shaped by timepoint; edges indicate similarity in expression profiles and gene-set overlap (top 10% of edges shown); node sizes are scaled by the number of genes in each module. **C**, UMAP plot of the T-cell compartment colored by expression level (module eigengene, ME) for lung pre-treatment M13 (MHC class II antigen presentation; top) and liver pre-treatment M13 (NK cell activation; bottom). **D**, Network plots as in (**B**), colored by Pearson correlations between the exhaustion (top) and cytotoxic (bottom) co-expression modules and selected consensus gene expression programs (cGEPs)^20^. **E**, Paired triangular heatmaps of gene co-expression network strength (topological overlap) from pre- (upper) and post-treatment (lower) for a module which was strongly preserved across treatment (liver pre-treatment M3, ribosomal protein synthesis). Genes in this heatmap are ordered by their eigengene-based connectivity (kME) in the pre-treatment timepoint (high connectivity genes in the bottom left). **F-G**, Module preservation and overlap analysis of T-cell co-expression modules between pre- (SCR, x-axis) and post-treatment (C02, y-axis) in liver (**F**) and lung (**G**) metastases. Dot color and size indicate the odds ratio and Jaccard index of gene-set overlap (top 100 genes by kME per module); significant overlaps (Fisher’s exact test, FDR < 0.05) are highlighted with a black outline. Marginal bar plots show *Z_summary_* topological preservation scores and preservation categories from NetRep^29^ (see **Methods** for description of the preservation categories). **H**, Network topology and expression level for liver pre-treatment M1 (T-cell activation and proliferation). Left: paired co-expression heatmaps as in (**E**). Right: T-cell UMAP plot colored by M1 expression level. **I**, Hub gene network for the top 25 hub genes by connectivity (kME) in liver pre-treatment. **J**, Network topology and expression level for lung pre-treatment M12 (T-cell activation and proliferation) as in (**H**). **K**, Hub gene network for the top 25 hub genes by kME in lung pre-treatment M12. **L**, Antigen-specific activation (ASA) scores from the T-cell annotator (TCAT)^20^. Left: T-cell UMAP plot colored by cell-level ASA score. Right: co-expression module network plot as in (**B**) colored by Pearson correlation between module expression levels and ASA scores. **M**, Comparison between mean log10(clonal proportion) and mean ASA score across T-cell populations in liver and lung metastases. Each point represents one population at one timepoint, sized by cell number; arrows indicate the trajectory from pre- to post-treatment.

To determine how dual ICI reshapes the architecture of T-cell transcriptional networks, we performed module preservation^28,29^ testing and gene-set overlap analysis between pre- and post-treatment T-cell co-expression networks (**Figures 3E-G**). In this framework, we test for changes across several different aspects of network architecture, where statistical significance identifies preserved regulatory units whose network topology remains stable across timepoints, while a lack of significance indicates substantial treatment-induced remodeling of the module’s internal co-expression structure (see **Methods**). For example, a ribosomal synthesis module (liver pre-treatment M3) exhibited strong one-to-one gene overlap and high topological preservation (Z_summary_ p < 0.05, NetRep^29^ permutation test) across timepoints (post-treatment M8, **Figure 3F**). Direct comparison of network connectivity confirmed that this ribosomal program remains topologically intact following ICI (**Figure 3E**). In contrast, this analysis shows that liver pre-treatment activation and proliferation module (M1) underwent extensive re-modeling (**Figures 3F and 3H**). This module fragmented into seven distinct post-treatment modules, and crucially, module preservation testing revealed that the gene co-expression strength within the M1 network was significantly attenuated post-treatment, indicating a loss of regulatory cohesion (*avg.weight* p = 1.0, NetRep^29^ permutation test). While this module is highly expressed in CD8+ resident exhausted T-cells (**Figures 3I and S13**) and is characterized by hub genes such as *ENTPD1*, *CTLA4*, *PDCD1*, and *CXCL13*, this fragmentation suggests that ICI-induced perturbations in the liver co-expression network leads to the dismantling, rather than productive activation, of this exhaustion-associated T-cell activation program. In sum, we found that three out of thirteen pre-treatment modules in the liver were remodeled (**Figure 3F**), while in the lung eight out of nineteen pre-treatment modules were remodeled (**Figure 3G**). Although a greater number of modules underwent structural shifts in the lung, these changes often represented refinement of pre-existing programs. This is in contrast to the collapse of regulatory hubs as seen in liver M1, which exhibited a near-complete loss of network connection strength post-treatment.

A markedly different trajectory was observed in the lung pre-treatment T-cell activation module M12. Although preservation testing indicated significant remodeling in terms of overall co-expression intensity (*avg.weight* p = 0.301, NetRep permutation test) and expression signatures (*cor.contrib* p = 0.179), we found that its internal co-expression weights remained comparatively robust (**Figures 3J and 3K**), suggesting that the program underwent a functional shift in response to therapy. This program represents a high-avidity axis of productive T-cell exhaustion, characterized by hub genes associated with both T-cell dysfunction and tissue residency, including *PDCD1*, *CDCR6*, *CXCR3*, and *VIM*. Notably, the liver pre-treatment M12 gene set did not significantly overlap with any of the liver pre-treatment modules (Fisher’s exact test, FDR > 0.05), highlighting it as a site-specific transcriptional state. Unlike the extensive fragmentation of liver M1, lung M12 significantly overlapped with only two post-treatment modules (Fisher’s exact test FDR < 0.05), and direct comparison of the network topology revealed that its internal co-expression weights remained robustly active (**Figure 3J**). This suggests that in the lung, the exhaustion-associated gene expression programs are refined rather than dismantled, sustaining a coordinated transcriptional architecture involving core TCR signaling components like *CD3E* and *CD3D*.

To determine whether these divergent networks reflect differences in functional T-cell engagement, we calculated antigen-specific activation (ASA) scores^20^ at single-cell resolution, and compared them with our co-expression modules (**Figure 3L**). Consistent with our co-expression network findings, ASA scores were enriched within the exhaustion- and HLA-associated modules, particularly in those defining the CD8+ resident exhausted T-cell niche. Comparing ASA scores to TCR-derived clonal expansion revealed that while CD8+ resident populations across metastatic sites exhibit a high degree of clonality, their functional activation states remain heterogeneous (**Figure 3M**). This suggests that clonal expansion alone is insufficient for clinical response in the pMMR/MSS mCRC setting, and highlights the need for a deeper investigation into the lineage-specific clonal dynamics and TCR repertoires that characterize these metastatic environments.

### Dual ICI induces site-specific T-cell clonal remodelling in liver and lung metastases

Building on our transcriptional characterization of the liver and lung metastatic TME under dual ICI treatment, we next investigated how these coordinated cellular changes were reflected in the T-cell receptor (TCR) repertoire. By integrating paired single-cell TCR sequencing with transcriptomic profiling, we resolved immunotherapy-induced T-clonal expansion dynamics while simultaneously linking TCR profiles to functional T-cell states. In total, we longitudinally profiled 203,380 tumor-derived TCR+ T cells from 24 patients and 59 V(D)J-T libraries from the SERPENTINE cohort. The MSS CRC Liver/Lung metastatic study subcohort encompassed 115,710 T cells and 58,087 clonotypes from 13 patients with matched pre-treatment (SCR) and post-treatment (C02) samples from liver (n=6) and lung (n=7) metastases (**Figure 4A and S22**).

**Figure 4.**
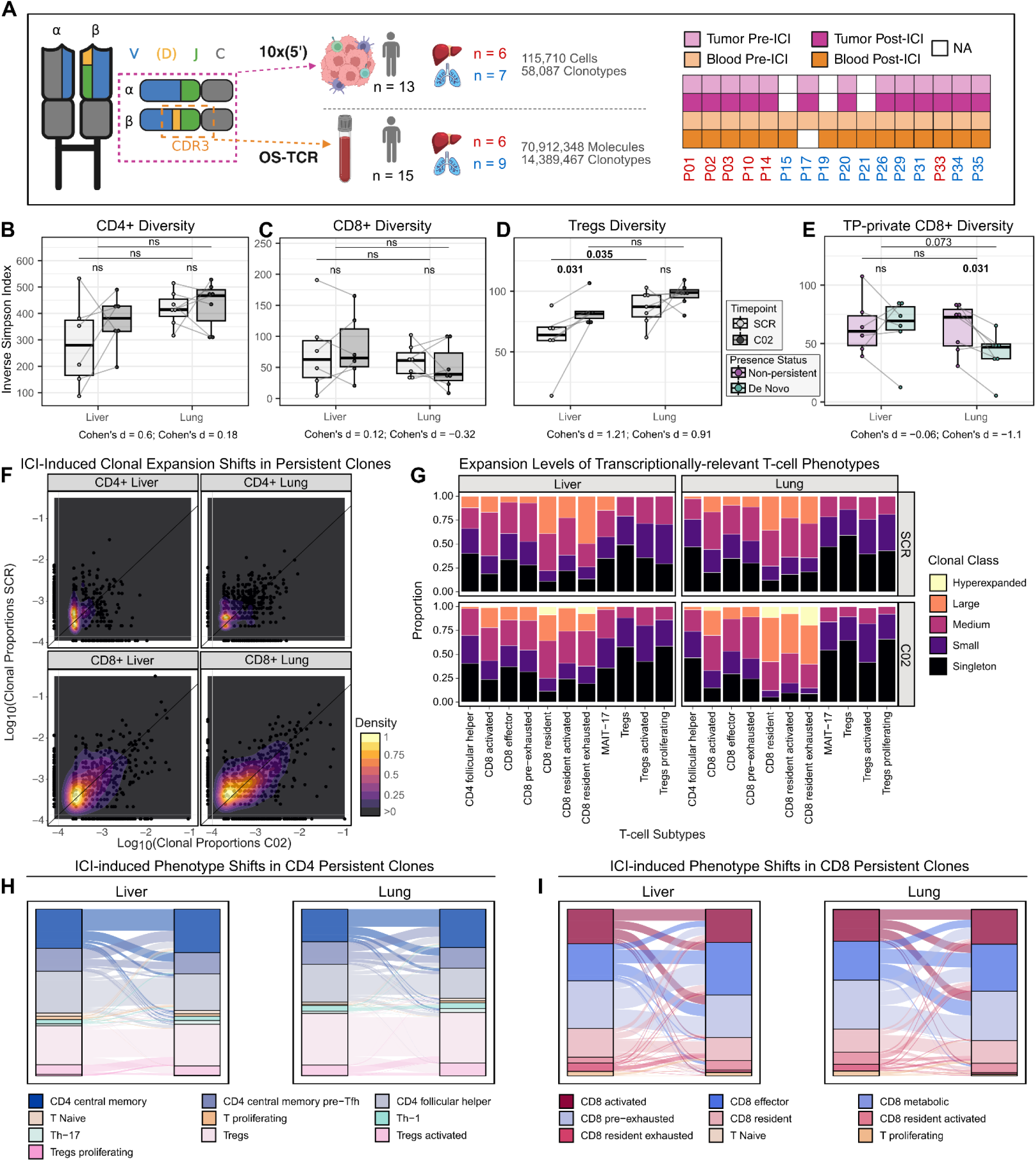
Dual ICI focuses CD8+ clones in lung metastases but diversifies TCRs in liver metastases. **A,** Single-cell TCR sequencing was performed on tumor biopsies (10x Genomics Chromium 5’) and blood samples (OS-TCR, Omniscope Inc.) of matched SCR and C02 time points (13 patients in tumor, 15 in blood). Tumor and blood clonotypes were defined and matched by their β chain CDR3 region. Illustrations were created with Biorender.com. **B-E**, TCR repertoire Inverse Simpson diversity index boxplots stratified by metastatic site across time points. Each dot represents the mean of an individual sample’s (or patient) repertoire diversity with lines connecting the same patient across time points, calculated from 1,000 bootstrap iterations with downsampling to a common depth (10th percentile of sample sizes). Effect sizes are reported as Cohen’s d for each paired comparison; p-values two-sided Wilcoxon signed-rank tests from paired comparisons (SCR vs. C02) and from Wilcoxon rank-sum tests from unpaired (liver vs. lung) comparisons at a given time point are shown. Non-significant comparisons are labeled “ns”; comparisons with p < 0.1 display the exact p-value; significant comparisons (p < 0.05) are shown in bold. Data are shown for CD4+ (**B**), CD8+ (**C**), Treg (**D**), and time point-unique CD8+ TCR repertoires (non-persistent vs. *de novo*; **E**). **F**, Clonal scatter of pre-treatment (SCR; y axis) versus post-treatment (C02; x axis) tumor clonotype expansion levels (clonal proportions), faceted by metastatic site and T-cell lineage. Each point represents a clonotype. Axes are log₁₀-transformed (with an added ε = 1×10⁻⁴) to accommodate the highly skewed distribution of single-cell TCR data. Two-dimensional kernel density estimates with contour overlays are shown on persistent clonotypes, with lighter colors indicating higher clonotype density. **G**, Distribution of global cell-level clonal expansion classes (see **Methods**) across T-cell subsets shown as stacked bar plots faceted by metastatic site and timepoints. **H-I**, Alluvial plots illustrating phenotype transitions of persistent clonotypes across timepoints for CD4+ (**H**) and CD8+ (**I**) conventional T cells. Flows represent aggregated cell type proportions per patient, with the y axis scaled to sum to one at each time point, enabling comparison of relative phenotypic distributions over time.

TME TCR repertoire diversity comparison between SCR and C02 revealed opposing ICI-driven trends between liver and lung metastases (**Figure S23A**), which we disentangled through T-cell lineage stratification, uncovering distinct CD4+ and CD8+ patterns concordant across all diversity metrics examined.

Within the CD4+ compartment, a diversification trend was observed in both liver (Inverse Simpson Index [IS] Cohen’s *d* [d] = 0.6) and lung (IS d = 0.18; **Figure 4B**), with D50 showing the greatest effect size (d = 0.62) and consistently higher values in lung at both timepoints (p = 0.051; **Figure S23B**). Conversely, CD8+ clones showed divergent trends, increasing in liver (IS d = 0.12) and decreasing in lung (IS d = 0.32; **Figures 4C and S23C**). At the effector subset level, these lineage trends were maintained and accentuated, reaching a Cohen’s d of 0.94 (CD4+) and 0.59 (CD8+) in liver, and 0.62 (CD4+) and 0.6 (CD8+) in lung for the most discriminant metric, suggesting a direct immunotherapy-driven effect on the effector pool (**Figures S23D and S24B**). Notably, Tregs showed therapy-induced diversification particularly in liver metastases (IS d = 1.21; two-sided Wilcoxon signed-rank test, p = 0.031), while baseline Treg diversity was significantly greater in lung compared to liver (two-sided Wilcoxon rank-sum test, p = 0.035; **Figures 4D and S24A**), pointing to a pre-existing clonal Treg activity potentially underlying the immunosuppressive nature of the liver TME. Together, these findings indicate a global T cell diversification in liver metastases and a lineage-specific clonal remodeling in lung under immunotherapy.

To further dissect mechanisms underlying divergent CD8+ clonal dynamics, we analyzed clonal persistence, loss, and *de novo* infiltration. Across both sites and both T lineages, persistent clones represented a minor fraction of the repertoire relative to non-persistent and *de novo* clones (two-sided Wilcoxon signed-rank test, p < 0.05; **Figure S25A**). Importantly, in lung metastases, newly infiltrating CD8+ clones exhibited significantly lower diversity (higher clonality) than non-persistent bystander clones (IS d = −1.1; two-sided Wilcoxon signed-rank test, p = 0.031) and liver metastases (two-sided Wilcoxon rank-sum test, p = 0.073; **Figures 4F and S24C**). This links CD8+ specialization to the influx of highly expanded clones and suggests a strong remodeling by newly infiltrating CD8+ clones only in lung lesions. Of note, the diversity of persistent CD8+ clones between SCR and C02 remained unchanged (**Figure S24D**); however, enhanced and contracted expansion of persistent clones was observed in lung and liver metastases, respectively (**Figures 4G and S25B**), suggesting treatment-induced expansion of resident clones in lung lesions.

We next sought to investigate joint T-cell phenotype and clonal ICI-induced dynamics. Within effector populations, expansion patterns were aligned with transcriptomic differential abundance analysis, with tissue-resident CD8+ populations in lung metastases expanding more post-treatment compared to liver lesions (**Figure 4G**). Analysis of phenotypic transitions among persistent clones further underscored the activity of dual ICI treatment, even in the absence of clinical responses. Across metastatic sites, CD4+ naïve and central memory clones transitioned into effector states following treatment, consistent with effective CTLA-4-mediated priming (**Figure 4H**). Additionally, pre-exhausted CD8+ clones acquired activated effector phenotypes, indicative of PD-L1-mediated reinvigoration (**Figure 4I**). Importantly, these therapy-associated differentiation programs were also evident in liver metastases, suggesting that dual ICI engages core T-cell activation pathways within this suppressive niche but fails to translate it into tumor control.

### Systems-level tracking of therapy-specific T-cell clones

Following investigation of local TCR repertoire remodeling, we assessed systemic longitudinal T-cell clonal expansion dynamics in peripheral blood using deep TCR repertoire sequencing (57,902,418 clonotypes; **Figure 5A**), illustrating consistent peripheral ICI-induced T-clonal expansion across the SERPENTINE cohort (**Figure 5B**). In line with the tumor TCR repertoire analyses, we restricted the analysis to MSS CRC patients with (n=6) or without liver metastases but lung metastases (n=9) with matched blood SCR and C02 data (14,389,467 clonotypes). Diversity analysis of blood TCR repertoires showed no differences globally (**Figure S27A**), however evenness increased post-treatment in patients without liver metastases (Pielou’s Evenness; two-sided Wilcoxon signed-rank test, p=0,02; **Figures 5C and S27b**). We next identified event-related differently contracted and expanded (DE) blood clones (see **Methods**); 85,232 clones (1,29%) were identified to be event-related (**Figure 5D**). We found a global significant enrichment of DE clones compared to contracted clones induced by the immunotherapy (two-sided Wilcoxon signed-rank test, p=1,2·10^−4^; **Figure 5E and S27C**), where the majority entered circulation *de novo* after treatment (two-sided Wilcoxon signed-rank test, p<0.05; **Figure 5F and 27D**). These results suggest that dual ICI drives priming of previously absent clones across both metastatic sites, reflecting a systemic T cell mobilization with potential antitumor specificity, while polyclonal broadening is selectively observed in patients with lung metastases, consistent with a broader and more effective T lymphocytic recruitment in response to therapy.

**Figure 5.**
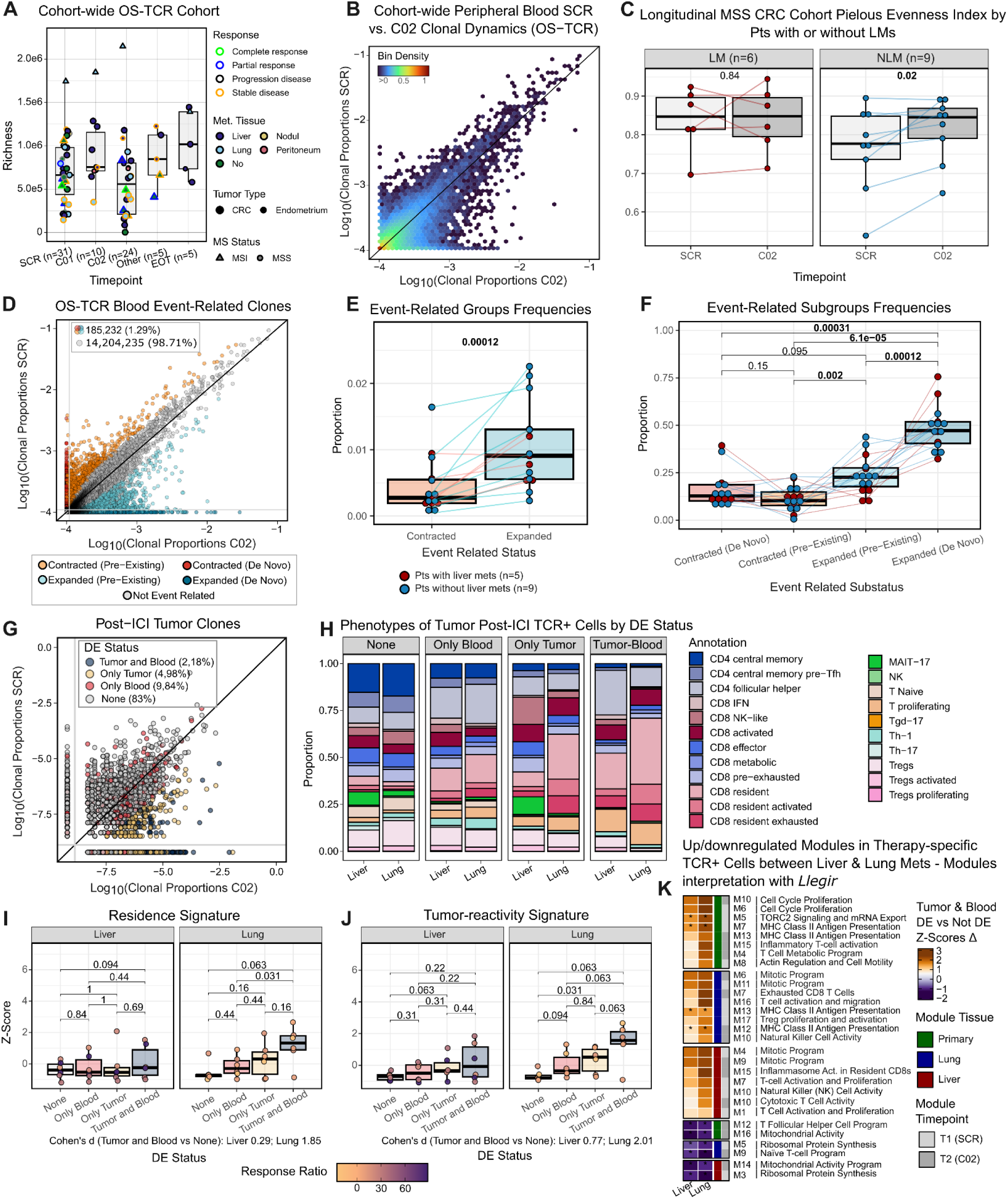
Peripheral blood TCR repertoire dynamics complement tumor-resident clonal responses and enable tracking of therapy-specific clones. **A,** Boxplots of patient blood TCR repertoire sizes (Richness: number of unique clonotypes) across timepoints (SCR: screening; C01: on treatment, 1st cycle; C02: on treatment, 2nd cycle; Other: other on-treatment cycles; EOT: end of treatment) on the entire Serpentine OS-TCR cohort. Dots represent per-patient richness. Inner coloring shows matching tumor sequenced tissue (10x Genomics), edge coloring shows clinical response at C02, size shows primary cancer type, and shape shows microsatellite status. **B,** Cohort-wide clonal scatter of blood pre-treatment (SCR, y axis) and post-treatment (C02, x axis) of blood clonotype expansion levels (clonal proportions) grouped into 50 bins of patients with matched SCR-C02 OS-TCR data. Axes are log₁₀-transformed (with an added ε = 1×10⁻⁴). Bins are colored by the scaled density where red indicates maximal density and dark blue minimal density. **C,** Comparison of blood OS-TCR repertoire Pielou Evenness index between pre-treatment (SCR) and post-treatment (C02) timepoints and split by liver metastasis status: liver (LM) or lung without liver (NLM). Dots represent per-patient diversity values, with lines connecting the same patient across time points. Dots are colored by whether patients have liver (red) or lung metastases (blue). P-values from two-sided Wilcoxon signed-rank tests are shown. **D,** Clonal scatter of blood pre-treatment (SCR, y axis) and post-treatment (C02, x axis) clonotype expansion levels (clonal proportions) of the study cohort patients. Axes are log₁₀-transformed (with an added ε = 1×10⁻⁴). Dots represent clonotypes and are colored by longitudinal event-related status (Omniscope Inc., proprietary noise model, log₂ fold change ≥ 2). **E-F,** Boxplots comparing the proportion of event-related groups (contracted vs. expanded) within the total blood TCR repertoire (**E**), and of event-related subgroups within the event-related pool (considering whether they are pre-existing or *de novo*; **F**). Dots represent per-patient proportions, with lines connecting the same patient across time points. P-values from two-sided Wilcoxon signed-rank tests are shown. **G,** Clonal scatter of tumor pre-treatment (SCR, y axis) and post-treatment (C02, x axis) clonotype expansion levels (clonal proportions) of the study cohort patients. Axes are log₁₀-transformed (with an added ε = 1×10⁻⁴). Dots represent clonotypes and post-ICI (C02) are colored by their compartment-wide differential expansion (DE) status. **H,** Stacked proportions of TCR+ cells’ level 2 annotations stratified by DE status and metastatic site. **I-J,** Z-scores of residence (**I**) and tumor reactivity (**J**) gene signatures computed with AUCell on post-ICI (C02) tumor TCR+ cells grouped by their clonal DE status and split by metastatic site. Dots correspond to signature-specific Z-scaled AUCell scores (scaled across patients and DE groups) and are colored by tumor growth at C02 (response ratio), with dark purple indicating the worst response and light orange the best. P-values of pairwise paired statistical significance assessment through two-sided Wilcoxon signed-rank tests across DE Status groups are shown. **K,** Heatmap showing the delta between Tumor-and-Blood DE and Not DE (None) Status groups of ULM Z-scaled scores of selected Organ-derived GEX co-expression modules computed on the tumor post-ICI (C02) clone TCR+ cells split by metastatic site (x axis). Positive Z-scores delta values shown in orange indicate higher expression of the module in the Tumor and Blood DE Status group, while negative values shown in purple indicate higher expression in the None DE Status group. Modules are functionally annotated with *Llegir* (see **Methods**). Statistical significance of paired differences between both DE Status groups was assessed through two-sided Wilcoxon signed-rank tests and corrected for multiple testing via the Benjamini-Hochberg method. The significance threshold was set at 0.05 and significant differences for each module and tissue are shown with an asterisk (*).

To uncover ICI-induced systemic clonal dynamics, we matched tumor and blood clonotypes to identify site-specific therapy-specific T-cell clones. We found consistently high overlap of repertoires in the tumor and blood across all patients (74.9% at SCR, 72.6% at C02) (**Figure S28A**), guiding our strategy to define therapy-specific clones based on their differential expansion status in both tumor and blood compartments. The majority of the 2,393 (7,15%) tumor DE clones were detectable in peripheral blood post-ICI, with a large fraction of *de novo* and persistent tumor DE clones showing also markedly differential expansion in blood (**Figure S28B-D**). We next stratified systemic therapy-specific clones (DE in tumor and blood, 2.18%) from tDE clones observed only in tumor (4.98%) or blood (9.84%) and bystander clones (83%) (**Figure 5G**). We then examined whether the phenotypic programs of these clones aligned with our previous findings of a coordinated and productive immune response in lung, contrasting with a diffuse and non-specific clonal engagement in the liver. In both sites, we found that systemic therapy-specific T-cells showed a reduction of CD4+ central memory and Tregs phenotypes, and an increase of CD8+ resident and proliferating states compared to their bystander counterparts, an observation more pronounced in lung lesions (**Figure 5H**). Scoring functional T-cell signatures revealed an enrichment of exhaustion, proliferation, and cytotoxicity programs (**Figure S29**), with an increase of residence and tumor-reactivity programs in lung therapy-specific T cells relative to bystanders (**Figures 5I and 5J**). We then scored gene co-expression modules from the primary CRC, lung and liver metastatic tissues (**Figures S30-S32**), functionally annotated with *llegir*, to interpret therapy-specific clonal transcriptional profiles (see **Methods**). In line with prior results, we observe an enrichment of *cell cycle proliferation / mitotic programs, MHC class II presentation* and cytotoxicity-like modules in therapy-specific T cells (therapy-specific vs. bystander Δ; **Figures 5K and S33**). Here, a higher expression of resident-like (Primary-T2 M4, Lung-T1 M15, Liver-T1 M1: modules with *ITGAE* as master regulator) and tumor-reactive-like modules (Lung-T1 M17, Lung-T2 M7, Liver-T2 M15: *CXCL13 and ENTPD1* as master regulators) emerged in lung metastases (liver vs. lung Δ; **Figures 5K and S34**). In contrast, modules associated with *naivety* and central memory (*mitochondrial activity*, *ribosomal protein synthesis*) were downregulated in therapy-specific T cells relative to bystanders (**Figure 5K**)

Altogether, longitudinal TCR repertoire profiling across tumor and peripheral blood, coupled with transcriptional profiling, provides a thorough framework for resolving therapy-induced clonal dynamics and defining therapy-specific clonotypes across metastatic sites. Leveraging this strategy, we uncover a conserved pattern of peripheral differential clonal expansion upon immunotherapy that translates into metastatic site-specific dynamics. In the liver, therapy drives a diffuse local diversification where therapy-specific clones acquire a proliferative, cytotoxic and exhausted phenotype but lack hallmarks of productive antitumor immunity. In contrast, lung metastases exhibit a coordinated peripheral diversification accompanied by a highly clonal CD8+ infiltrate, where therapy-specific clones are further distinguished by a tumor-reactive, tissue-resident program of heightened immunogenic potential, likely underlying the more favorable prognosis of this patient subset. These findings highlight the power of integrating deep multi-compartment clonal tracking with transcriptional profiling to dissect site-specific antitumor immune responses and identify functional determinants of productive immunotherapy in mCRC.

### Immune coordination shapes the trajectory of malignant remodeling

We next investigated the relationship between metastasis site-specific immune microenvironments and the malignant compartment. Using the same approach as the T-cells analysis, we integrated tumor cells from liver and lung metastases with the external primary tumor^25^ dataset, identifying 20 transcriptomically distinct clusters representing a spectrum of CRC transcriptomic states (**Figure 6A**). The distribution of these cells mapping to each malignant cluster varied across patients and metastatic sites, yet the majority of these clusters represent cross-tissue malignant states (**Figure 6B**).

**Figure 6.**
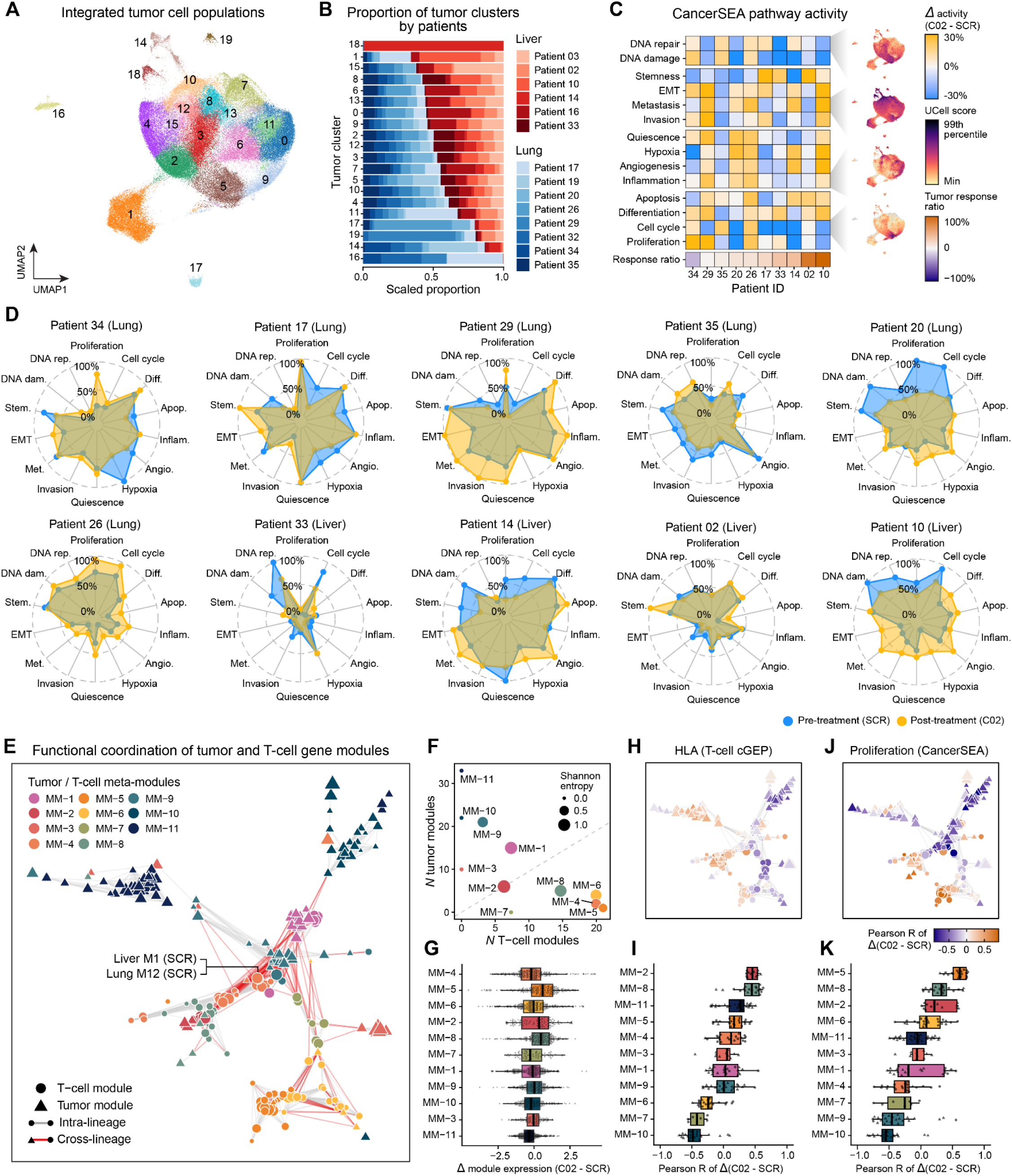
Malignant remodeling under dual ICI is coupled to T-cell dynamics. **A**, Uniform manifold approximation and projection (UMAP) of the integrated tumor cells from the liver and lung metastasis samples of the SERPENTINE cohort (n=40,694 cells) and primary CRC TMEs from an external dataset^25^ (n=72,952 cells). Points are colored by Leiden cluster^62^ assignments. **B**, Stacked bar chart showing the scaled proportion of cells from SERPENTINE liver and lung metastasis patients mapping to each of the 20 tumor cell populations. **C**, Heatmap showing the treatment-induced change (Δ C02 - SCR) in CancerSEA^30^ functional state activity (decoupleR^57^ univariate linear model scores) post-treatment versus pre-treatment. Orange color indicates higher activity post-treatment, and blue color indicates higher activity pre-treatment. Patients are ordered by tumor response ratio (bottom row). UMAP insets show single-cell activity of representative programs (UCell^44^ scores). **D**, Radar plots for individual patients illustrating the functional remodeling of 14 CancerSEA programs from pre-treatment (blue) to post-treatment (orange); scores were min–max scaled across the cohort. For (**C**-**D**), only patients with >75 tumor cells for both timepoints were included in the analysis. **E**, Cross-lineage gene co-expression module coordination network, where nodes represent T-cell (circles, n=95) and tumor (triangles, n=119) modules. Nodes are colored by meta-module (MM) Leiden cluster assignments; node sizes are scaled by the number of genes in each module. Edges combine a weighted combination of the correlation of treatment-induced dynamics (Δ C02 - SCR) between modules, and the overlap in their constituent gene sets (Jaccard index); edges between modules of the same lineage are shown in gray, and cross-lineage edges in red.. Edges were downsampled in the plot for enhanced visual clarity. **F**, Scatter plot comparing the number of T-cell and tumor modules per meta-module; point size indicates lineage Shannon entropy (higher values indicate greater mixing of T-cell and tumor modules). **G**, Distributions of treatment-induced expression dynamics (Δ C02 - SCR) per patient and co-expression module, stratified by cross-lineage meta-modules, ranked by overall variance of the meta-modules and ranked by variance. **H**, Coordination network from panel (**E**), colored by Pearson correlation between the treatment-induced dynamics of each module (Δ C02 - SCR) and that of the HLA class II T-cell consensus gene expression program (cGEP^20^). **I**, The same data shown in (**H**), displayed as box-and-whisker plots stratified by meta-module (each point represents one module). **J**, Coordination network from panel (**E**), colored by Pearson correlation between the treatment-induced dynamics of each module (Δ C02 - SCR) and that of the proliferation signature from CancerSEA^30^. **K**, The same data shown in (**J**), displayed as box-and-whisker plots stratified by meta-module. For box-and-whisker plots, box boundaries and lines correspond to the interquartile range (IQR) and median, respectively. Whiskers extend to the lowest or highest data points that are no further from 1.5 times the IQR from the box boundaries. MM: meta-module.

To describe the molecular response to dual ICI within the malignant compartment of individual patients, we quantified the activity of 14 canonical cancer functional states (CancerSEA^30^, **Figure 6C**). This revealed a diverse array of malignant plasticity, suggesting that therapeutic pressure acts as a selective filter rather than a uniform inhibitor of tumor fitness. For example, in patient 34 (lung metastasis) we observed a down-regulation in hypoxia, metastasis, and stemness programs, along with a simultaneous increase in epithelial to mesenchymal transition (EMT) and proliferation signatures, indicating a high-fitness residual pool (**Figure 6D**). Conversely, in one of the patients with progressive disease (patient 10, liver metastasis) while proliferation was diminished, we observed an increase in EMT, angiogenesis, and metastasis programs (**Figure 6D**). These divergent yet partially-overlapping patterns suggest that in cases of partial response or therapeutic resistance, ICI may act as a selective filter, leaving behind a pool of malignant cells primed for survival and progression in the metastatic niche.

We reasoned that the remodeling of malignant gene expression programs is fundamentally coupled to those of the immune niche. To characterize this relationship, we performed a cross-lineage coordination analysis comparing treatment-induced gene expression changes between T-cell and malignant gene co-expression modules. Similar to our T-cell gene co-expression network analysis, we used hdWGCNA^27^ to identify 119 modules of highly co-expressed genes in the malignant compartment across tissues and treatment, and annotated them with llegir (**Figures S35-S42**; **Tables S8 and S9**). We calculated the change in expression level (Δ C02 - SCR) for all T-cell and malignant modules per patient, and then constructed a cross-lineage coordination network based on the similarity of treatment-induced gene expression dynamics across modules (**Figure 6E**; see **Methods**). Modules were grouped into 11 cross-lineage meta-modules (MMs), seven of which contained both T-cell and malignant modules, identifying them as potential interfaces of immune-malignant crosstalk (**Figure 6F**). The llegir annotations revealed coherent biological themes within each MM. Two cross-lineage MMs captured essential biological programs shared by both lineages, namely translation and ribosomal protein synthesis (MM-1) and mitotic proliferation (MM-2). MM-8 joined T-cell MHC class II antigen presentation and exhausted CD8+ T-cell programs with goblet cell and mucinous secretory programs from the malignant compartment. MM-4 was dominated by T-cell activation modules (liver pre-treatment M1 and lung pre-treatment M12) and proliferating Treg programs, alongside a tumor MHC class I antigen presentation and interferon module. MM-4 also exhibited the highest variance in treatment-induced dynamics, reflecting the high inter-patient heterogeneity in response (**Figure 6G**). Correlation of module dynamics with canonical gene programs (T-cell cGEPs and CancerSEA signatures) revealed which modules and meta-modules have concordant or discordant treatment-induced dynamics with these programs (**Figures S42 and S44**). Although MM-4 encompasses a major axis of T-cell activation and regulatory remodeling, its dynamics were only weakly coupled to the T-cell HLA program (**Figures 6H and 6I**) and were instead anti-correlated with tumor proliferation (**Figures 6J and 6K**). Conversely, tumor proliferation increased in concert with MM-5 (CD8+ metabolic and cytotoxic) programs. This suggests that effector and tumor-reactive T-cell programs can be engaged in these patients without restraining tumor growth, representing potential niches of therapeutic resistance. Collectively, these results suggest that the clinical failure of dual ICI is not merely a consequence of poor immune infiltration, but a breakdown in immune coordination, where T-cell programs fail to synchronize with and reverse malignant progression.

### Modeling patient survival as a function of malignant gene expression programs

We leveraged an interpretable machine learning approach to associate gene modules with patient survival in the TCGA CRC bulk RNA-seq dataset (**Figure 7A**, see **Methods**). We trained an XGBoost regression model on the gene expression signatures of the malignant gene modules and computed SHapley Additive exPlanations (SHAP) values for each module in each TCGA patient, allowing us to rank modules by their contribution to predicted survival. For the top-ranked modules, the relationship between module expression and SHAP values revealed sharp transitions that naturally stratified patients into risk groups. For example, high expression of liver pre-treatment M17 (cell signaling; *ZSWIM6, PPP3CA, SPTBN1*) was associated with higher predicted survival and low expression with lower predicted survival. Conversely, high expression of liver pre-treatment M9 (immediate-early genes and stress response) was associated with lower predicted survival. We used the transition point in each module’s expression-SHAP relationship to define data-driven expression cutoffs and compared overall survival between the resulting high- and low-expression groups (**Figures 7B-D**). For 13 of the 24 top-ranked modules, these cutoffs identified groups with significantly different overall survival (log-rank test, FDR < 0.05). In all three tissue contexts, high expression of the top-ranked module was associated with longer overall survival.

**Figure 7.**
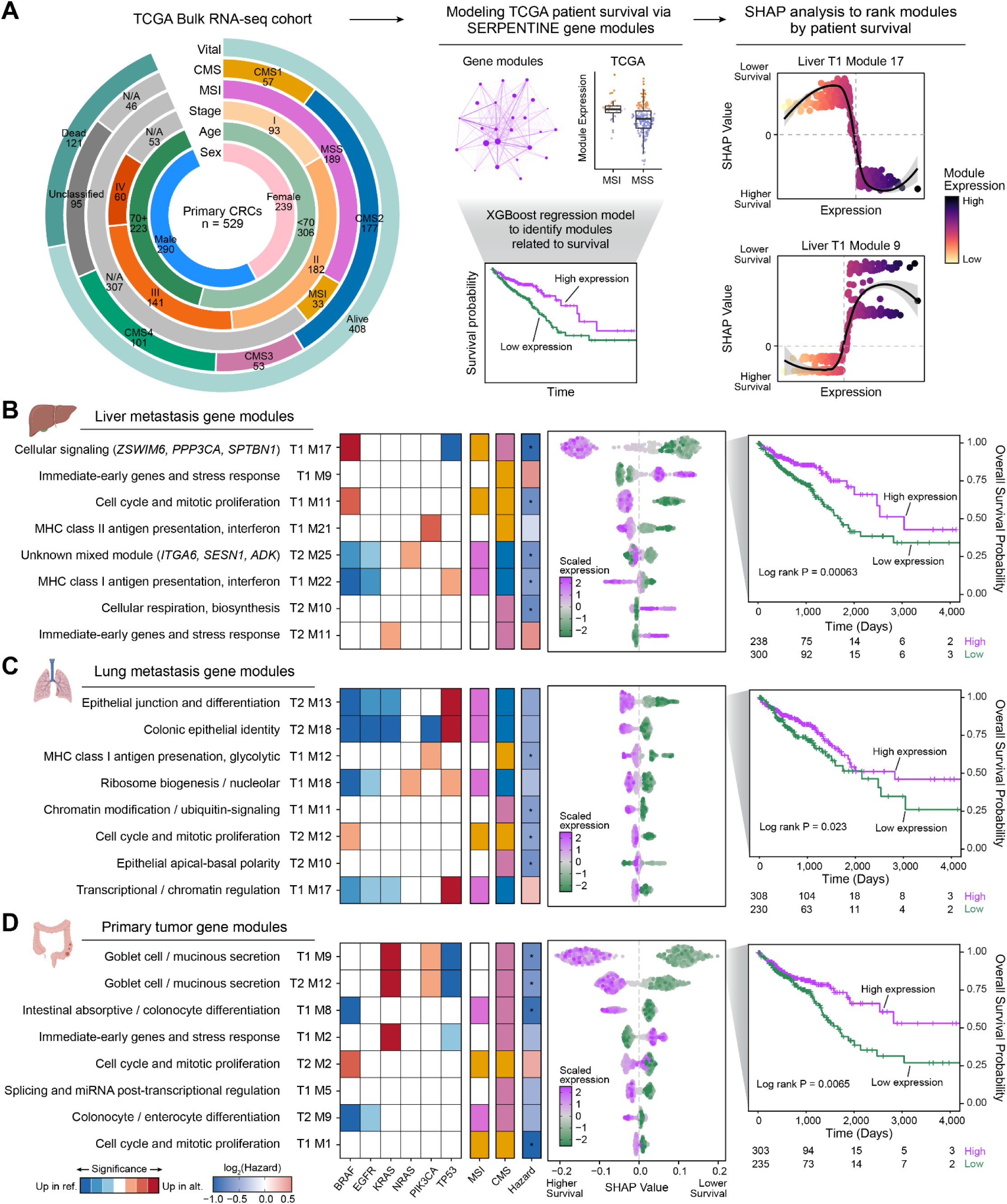
Malignant gene programs recapitulate CRC molecular subtypes and stratify patient survival. **A,** Left: characteristics of the TCGA CRC bulk RNA-seq cohort (n = 538 samples from n = 529 patients) used in this analysis, shown as a circular proportional bar chart. N/A indicates missing data. Middle: overview of the analysis modeling TCGA patient survival using the tumor gene co-expression modules identified in the malignant fraction of the scRNA-seq dataset. For each module, gene expression scores were calculated in the TCGA cohort using decoupleR^57^, enabling comparisons with clinical attributes such as MSI status. An XGBoost model with a Cox proportional hazards objective was trained to predict patient survival using only these gene co-expression modules as input features, and SHapley Additive exPlanations (SHAP) values were calculated to infer feature importance. Right: gene expression vs. SHAP values for two selected gene modules. A positive SHAP value indicates a higher risk of death (lower survival), whereas a negative SHAP value indicates a lower risk of death (higher survival). **B-D**, Layered plots for the eight most informative modules per tissue (ranked by mean absolute SHAP value) (**B**, top: liver metastasis; **C**, middle: lung metastasis; **D**, bottom: primary tumor). Module names indicate the network of origin: T1, pre-treatment (SCR); T2, post-treatment (C02). Left: heatmap showing associations between module expression and clinical attributes: driver mutation status (*BRAF*, *EGFR*, *KRAS*, *NRAS*, *PIK3CA*, *TP53*), MSI status, and consensus molecular subtype (CMS). Mutation status (mutant vs. wild-type) and MSI status (MSI vs. MSS) were compared using two-sided Wilcoxon rank-sum tests, with color indicating the direction of the difference (higher in the reference or alternative group) and intensity indicating significance (Benjamini–Hochberg FDR < 0.05, < 0.01, or < 0.001). CMS associations were tested using the Kruskal–Wallis test, with color indicating the CMS with the highest module expression. P-values were adjusted for multiple testing using the Benjamini–Hochberg procedure (**Table S10**). Hazard ratios were calculated by defining high- and low-expression groups at the characteristic transition in the gene expression vs. SHAP value curve; * indicates log-rank FDR < 0.05. Middle: beeswarm plot showing the distribution of SHAP values, where each dot represents one TCGA sample, colored by scaled gene expression level. Right: Kaplan–Meier curves for the most informative module in each tissue, splitting patients into high- and low-expression groups. Illustrations were created with Biorender.com.

We next examined associations between module expression and additional TCGA patient attributes, including microsatellite status, consensus molecular subtype (CMS)^5^ of the tumor, and mutation status of key CRC driver genes (*BRAF, EGFR, KRAS, NRAS, PIK3CA,* and *TP53*; **Table S10**; see **Methods**). Proliferative and cell cycle modules were enriched in MSI and CMS1 tumors, and in *BRAF*-mutant tumors, supporting the known co-occurrence of these features^5^. Epithelial identity modules from lung metastases (post-treatment M13 and M18) were enriched in *TP53*-mutant, MSS, and CMS2 tumors while depleted in *BRAF*, *EGFR*, and *KRAS-*mutant tumors, consistent with the epithelial phenotype of CMS2. (**Figure 7C**). Goblet cell and mucinous secretion modules from the primary tumor (pre-treatment M9, post-treatment M12) were enriched in *KRAS*- and *PIK3CA*-mutant tumors, depleted in *TP53*-mutant tumors, and associated with CMS3 (**Figure 7D**). Together, these results show that malignant programs resolved at single-cell resolution in a longitudinal clinical setting generalize to more than 500 independent patients, where they recapitulate the established molecular architecture of CRC and stratify overall survival, positioning these gene modules as clinically relevant readouts of tumor biology.

## Discussion

In this phase II clinical trial, we combined dual ICI therapy with longitudinal single-cell and immune repertoire profiling to dissect the mechanisms underlying resistance and response in metastatic pMMR/MSS colorectal cancer. By integrating clinical outcomes with high-resolution analyses of immune coordination, transcriptional network dynamics, and T-cell clonal remodeling across metastatic sites, the SERPENTINE trial provides a systems-level framework to understand why immunotherapy remains largely ineffective in this disease and why this failure is not uniform across metastatic niches. Our data indicate that in liver metastases, therapeutic resistance reflects a failure to direct antitumor immune response.

Immunotherapy has so far failed to demonstrate clinical benefit in metastatic pMMR/MSS CRC, and clinical trials exploring immunotherapy-based combinations have achieved only modest results. The STELLAR-303 study was the first phase III clinical trial to show a statistically significant improvement in overall survival with an immunotherapy-based regimen zanzalintinib–atezolizumab, albeit the magnitude of benefit was not clinically meaningful^31^. The C-800-01 phase I trial evaluated botensilimab, a multifunctional Fc-enhanced anti-CTLA-4 antibody, in combination with balstilimab, a high affinity antiPD-1, based on the premise that dual PD-L1/CTLA-4 blockade may enhance T cell priming and clonal diversity^11^. Objective responses observed in the trial occurred in patients with non MSI-H/-dMMR that did not present active liver metastases. These results prompted the design of the phase II trial C-800-25 (NCT05608044), which selectively enrolled patients with refractory pMMR/MSS CRC without active liver metastases to receive botensilimab monotherapy or in combination with balstilimab. Despite these encouraging signals, the mechanistic basis by which liver metastasis constrains immunotherapy efficacy remains ill-defined, largely owing to the lack of integrated longitudinal, high-resolution translational analyses into clinical studies that enable an in-depth interrogation of cellular states, spatial organization, and clonal architecture upon immunotherapy treatment^15–17^.

The SERPENTINE trial was designed to address this gap by combining dual PD-L1 and CTLA-4 blockade with paired metastatic biopsies and longitudinal blood sampling, enabling prospective, single-cell-resolved interrogation of metastatic tumor-immune microenvironments over time. Clinically, all patients with pMMR/MSS mCRC with liver metastases experienced disease progression as best response, whereas a subset of patients without liver metastases achieved sustained disease control for up to nine months. Clinical benefit was pronounced in the population lacking liver disease, underscoring the concept that the site of metastasis critically determines immunotherapy responsiveness in this patient population.

At the single-cell level, dual ICI treatment induced extensive immune remodeling across metastatic sites, but with sharply divergent architectures. Lung metastases exhibited expansion of proliferative and tissue-resident CD8⁺ T cells, consistent with effective local immune engagement. In contrast, liver metastases exhibited enrichment of regulatory T cells, NK/NKT populations, and suppressive myeloid subsets, suggesting preferential reinforcement of immunosuppressive programs^32^. These differences were further reflected at the systems level. Immune co-abundance network analysis revealed sparse, modular networks in lung metastases centered on tissue-resident CD8⁺ T cells, consistent with productive immune coordination. By contrast, liver metastases displayed dense, highly interconnected networks enriched for regulatory and innate immune populations. In this context, effector T-cell populations occupied central but constrained positions, suggesting functional sequestration within an inhibitory multicellular network. These findings suggest that resistance induced by liver metastasis results from maladaptive immune network topology rather than absence of immune activation. Transcriptional network analysis reinforced this distinction. In lung metastases, an exhaustion-associated T-cell activation program was refined rather than dismantled following treatment, retaining robust internal co-expression and coordinated expression of checkpoint and TCR signaling genes. In contrast, the analogous program in liver metastases underwent extensive fragmentation, with loss of coordinated regulation of key genes including *CTLA4*, *ENTPD1*, *CXCL13*, and *ITGAE*. This fragmentation may help explain why immune activation signals in the liver fail to translate into sustained effector function^33^.

Analysis of TCR repertoires revealed site-specific clonal remodeling in response to immunotherapy. Lung metastases showed clonal focusing with expansion of highly clonal, tissue-resident and newly infiltrating tumor-reactive CD8⁺ T cells, whereas liver metastases exhibited a trend towards broad CD4⁺ and CD8⁺ repertoire diversification, most pronounced among regulatory T cells, driven by influx of weakly expanded clones and contraction of pre-existing expanded populations. Peripheral blood TCR profiling demonstrated robust systemic immune activation across metastatic sites, with therapy-expanded clones readily detectable in circulation and overlapping with tumor repertoires. Yet these therapy-specific clones acquired tissue-resident and tumor-reactive programs predominantly in lung metastases. These findings indicate that resistance in liver metastases does not reflect impaired systemic priming, but rather local reshaping or neutralization of incoming immune responses, uncoupling systemic activation from tumor control^19^. Integration of immune and malignant gene co-expression programs further showed that tumor cell states are coupled to immune dynamics: tumor proliferation increased together with cytotoxic CD8⁺ T-cell programs, indicating effector engagement that fails to restrain tumor growth. At the patient level, malignant remodeling was heterogeneous, with induction of EMT, angiogenesis, and metastasis programs in individual patients with progressive liver disease^34^. Notably, these malignant programs generalized beyond our cohort, recapitulating established CRC molecular subtypes and stratifying overall survival in more than 500 patients from TCGA in an analysis guided by an interpretable machine learning approach.

This study has limitations. First, metastatic site distribution and biopsy accessibility constrained tissue sampling, as liver and lung metastases are the most accessible lesions for percutaneous biopsy due to their comparatively lower procedural risk^35^. Consequently, our study primarily compares hepatic with non-hepatic disease, and additional metastatic sites in these patients remained uncharacterized at the tissue level. Second, as is inherent to prospective trials with longitudinal biopsies, the number of patients with matched pre- and post-treatment samples was determined by enrollment and biopsy feasibility. Therefore we sought to support our findings by converging evidence from complementary data modalities (cellular composition, gene networks, and tumor and blood TCR repertoires) and by assessing the clinical relevance of malignant programs in an independent cohort of more than 500 patients. Finally the external primary tumor dataset used for comparison was treated with neoadjuvant PD-1 blockade rather than dual PD-L1 and CTLA-4 blockade, which limits direct comparison with the metastatic cohort. Nevertheless, the analysis of longitudinal tumor immune microenvironment, tumor cell plasticity and systemic T-cell clonal dynamics in tumor and peripheral blood mitigated this limitation by providing deep, high-resolution insights and dependencies across cell types, states and metastatic sites.

Collectively, these data support a unifying model in which resistance to immunotherapy in pMMR/MSS mCRC, particularly in liver metastases, arises from maladaptive immune coordination across cellular, transcriptional, and clonal scales. Dual checkpoint blockade engages core immunological programs even within the liver, but these responses are redirected into fragmented transcriptional states, diluted clonal repertoires, and suppressive multicellular networks that fail to achieve effective tumor control^36^. These findings suggest that future immunotherapy strategies in pMMR/MSS colorectal cancer may need to move beyond intensifying immune activation and instead focus on rewiring immune coordination within suppressive metastatic niches such as the liver. Targeting regulatory feedback loops, macrophage-mediated suppression, or spatial immune organization, potentially in combination with liver-directed therapies, may be required to convert immune remodeling into durable clinical benefit^21^. More broadly, immune coordination may represent a generalizable framework for understanding and overcoming resistance to immunotherapy in immune-refractory solid tumors. Methodologically, this study also introduces **llegir**, an open-source software package which assembles traceable evidence for data-driven gene modules and uses a large language model to draft biological annotations, easing a key bottleneck in interpreting co-expression networks from complex single-cell studies.

These findings indicate that successful checkpoint blockade is determined less by the magnitude of immune activation than by its organization. Whereas lung metastases channel systemic immune priming into coordinated tumor-reactive CD8⁺ responses with clonal focusing, liver metastases redirect this activation toward broad inflammatory remodeling involving innate and regulatory populations, uncoupling immune activation from tumor control. These findings reframe organ-specific resistance as a failure of immune coordination rather than immune induction and provides a foundation for site-adapted immunotherapy strategies.

## Materials & Methods

### Clinical trial design and participants

The SERPENTINE trial is an open-label, unicentric multicohort phase II study performed in a tertiary universitary hospital located in Barcelona (Spain) in patients with advanced colorectal or endometrial cancer. Eligible patients were aged 18 years or older. In cohort 1, patients had confirmed metastatic adenocarcinoma of the colon or rectum or histologically confirmed endometrial cancer, not amenable to surgery, with known tumor MSI or dMMR status according to local standard practice, regardless of prior chemotherapy or targeted therapy. For cohort 2, patients had confirmed pMMR/MSS metastatic colorectal adenocarcinoma that had progressed on, or was refractory or intolerant to previous therapy with fluoropyrimidine, irinotecan, and oxaliplatin, with or without an anti-VEGF monoclonal antibody and, if *RA*S wild-type, an anti-EGFR monoclonal antibody; or pMMR/MSS endometrial cancer that had progressed during or after platinum- and taxane-based therapy.

Measurable disease, determined by investigators using Response Evaluation Criteria in Solid Tumours (RECIST) version 1.1, and an Eastern Cooperative Oncology Group performance status of 0–1 were required. Previous treatment with PD-L1, PD-1 or CTLA-4 targeting immune checkpoint inhibitors was not allowed. To characterize the intrinsic and baseline characteristics and the changes that arise on and after immunotherapy that could be modulating the immune response, sequential tumor biopsies were performed, including prior to treatment initiation and at C1D28 timepoints, when safely feasible, as well as at the time of maximal response and at disease progression. Liquid biopsies prior to treatment initiation and before each cycle were obtained to characterize peripheric changes. This trial was approved by the institutional review board and conducted in accordance with requirements of the regulatory authorities and provisions of the Declaration of Helsinki and Good Clinical Practice guidelines of the International Council for Harmonisation. Patients provided written informed consent. The trial is registered with the identifier NCT06680739. The full trial protocol and the Statistical Analysis Plan can be accessed in the Supplementary Material.

Patients with dMMR/MSI mCRC or mEC were randomized to either cohort 1A (durvalumab 1500 mg via IV infusion Q4W starting at Week 0) or cohort 1B (durvalumab 1500 mg plus tremelimumab 300 mg×1 dose at Week 0, followed by durvalumab 1500 mg monotherapy Q4W) using computer generated-block randomisation schedules at 1:1 ratio, while all patients in cohort 2 received durvalumab 1500 mg plus tremelimumab 300 mg×1 dose at Week 0, followed by durvalumab 1500 mg monotherapy Q4W until progression disease, unacceptable toxicity or completion of treatment for 24 months. Primary endpoint was objective response rate (ORR) according to Response Evaluation Criteria in Solid Tumors (RECIST) v1.1, while clinical secondary endpoints included progression free survival (PFS), defined as time from treatment initiation to progressive disease, and toxicity assessment, graded according to the National Cancer Institute Common Terminology Criteria for Adverse Events (NCI CTCAE) v5.0. Translational endpoints included characterization at a single cell resolution of the transcriptome of the tumors at baseline and upon treatment with immunotherapy, also including at the time of maximal response and at progression if feasible and correlation of the antitumor activity of durvalumab and durvalumab plus tremelimumab with other biomarkers on tumor tissue and in blood samples.

After inclusion of 5 patients with EC (4 pMMR/MSS, 1 dMMR/MSI), enrollment of patients with this tumor type was finalized due to lack of activity and the recruitment was limited to mCRC. Given the growing evidence of the presence of liver metastases as a biomarker of immune resistance, a subgroup analysis of the population with liver metastases versus non-active liver metastases was planned for patients with CRC. This report is based on a planned analysis (data cutoff January 2, 2026). The trial continues to the final overall survival analysis. Progression-free survival was estimated using the Kaplan–Meier method and log-rank test was used for statistical comparison. Univariable Cox proportional-hazard models were used to obtain hazard ratios (HRs) with 95% CIs. Two-sided 95% exact CIs were used to describe objective response and disease control in each group. The median follow-up time was calculated using the reverse Kaplan-Meier method.

### scRNA-sequencing

Fresh human metastatic colon cancer core needle biopsies were transferred immediately after obtention from Vall d’Hebron Hospital to the Single Cell Laboratory at CNAG, both located within Barcelona (Spain). Tissue fragments were placed on a pre-chilled Petri dish on ice, washed three times with cold PBS (Thermo Fisher Scientific), and minced with a pre-chilled razor blade until well homogenized. Minced tissues were collected from the plate using 3 ml of pre-warmed digestion medium (2.5 mg/ml Liberase TM [Roche] and 0.1 mg/ml DNase I [Worthington Biochemical] in RPMI medium without FBS) and transferred with a wide-bore pipette tip into a 15 ml Falcon tube. Samples were digested for 30–45 min at 37 °C in a horizontal shaker, pipetting occasionally with a wide-bore P1000 to mechanically dissociate larger fragments. After digestion, samples were kept on ice and assessed under the microscope using Trypan Blue. They were then filtered through a 70 µm filter into a new 15 ml Falcon tube. RPMI supplemented with 10% FBS (Thermo Fisher Scientific) was used to wash the tube and filter to a final volume of 10 ml. Samples were centrifuged at 400 × g for 7 min at 4 °C, and the pellet was resuspended in 100 µl of PBS/1% BSA (Miltenyi Biotec) and prepared for FACS isolation. To separate the immune fraction from the non-immune/tumoral fraction, cells were stained with PerCP/Cy5.5 anti-human CD45 (BioLegend #304028) for 20 min at 4 °C in the dark. Red blood cell lysis was subsequently performed using 1 ml of 1× RBC Lysis Solution (Miltenyi Biotec) for 3 min at room temperature. Cells were centrifuged at 350 × g for 10 min at room temperature, the supernatant was discarded, and the pellet was resuspended in 1 ml of cold PBS/1% BSA, followed by filtration through a 40 µm filter. After DAPI addition, live cells were FACS-sorted for CD45⁺/⁻ populations using the Melody flow cytometer (Becton Dickinson, Franklin Lakes, NJ). After sorting, cells were concentrated by centrifugation at 400 × g for 10 min at 4 °C and counted using the Luna-FL cell counter (Logos Biosystems), before proceeding to 10x Genomics 5′ single-cell RNA sequencing. Depending on the number of cells sorted for each fraction, samples were either loaded twice as a pool containing 50% CD45⁺ and 50% CD45⁻ cells or loaded once per fraction targeting maximal recovery (10,000 cells per reaction) when possible. Cells were partitioned into Gel Bead-In-Emulsions (GEMs) using the Chromium Next GEM Single Cell 5′ Reagent Kit v2 (10x Genomics, PN-1000263) and library preparation was performed according to the manufacturer’s protocol (CG000331). Briefly, amplified full-length cDNA, enriched T cell receptor (TCR) cDNA, and final sequencing libraries were quality-checked and quantified using the High Sensitivity DNA chip on an Agilent Bioanalyzer (Agilent Technologies). Sequencing was performed on an Illumina NovaSeq 6000 system, targeting approximately 40,000 read pairs per cell for gene expression (GEX) libraries and 10,000 read pairs per cell for TCR libraries. Sequencing conditions were as follows: Read 1 – 28 bp; i7 index – 10 bp; i5 index – 10 bp; Read 2 – 90 bp.

### Preprocessing single-cell sequencing data

scRNA-seq and scTCR-seq reads were processed with Cell Ranger multi (version 7.0.0), which uses Cell Ranger count to map scRNA-seq reads to the human reference transcriptome provided by 10X Genomics (refdata-gex-GRCh38-2020-A) and Cell Ranger vdj to map scTCR-seq reads to the human V(D)J reference provided by 10X Genomics (refdata-cellranger-vdj-GRCh38-alts-ensembl-5.0.0). Doublet scores, representing the likelihood of cellular barcode collisions, often referred to as were calculated separately for each scRNA-seq library using Scrublet^37^ (version 0.2.3) with default settings. We merged the resulting gene expression matrices from each scRNA-seq replicate into a single AnnData^38^ object for downstream analysis, totaling 486,410 barcodes and 36,601 genes from 84 sequencing libraries before additional quality control (QC) filtering.

We applied an initial dataset-wide filter to retain barcodes with 600 or greater unique molecular identifiers (UMI) and 50,000 or fewer UMIs, 600 or greater genes expressed, and fewer than 50% mitochondrial reads, yielding 456,653 barcodes. At this stage it is important to use a relaxed threshold to avoid removing tumor cells, which generally have increased mitochondrial activity compared to immune and stromal cells in these tissues and we perform additional filtering on mitochondrial reads in our lineage-specific clustering analyses. For interoperability between R and Python bioinformatic ecosystems, we formatted our datasets as both anndata objects and Seurat^39^ objects.

#### Initial clustering analysis grouping single cells into major lineages

The goal of this initial clustering analysis was to identify the major lineages of the TME: T cells, B and plasma cells, myeloid, stromal, and tumor/epithelial. We performed clustering analysis of our scRNA-seq dataset and the primary tumor dataset using SCANPY^38^ (version 1.9.8) UMI counts were library-size normalized and log-transformed using the functions `sc.pp.normalize_total()` and `sc.pp.log1p()`. Highly variable genes were identified using `sc.pp.highly_variable_genes()`. We performed principal component analysis (PCA) on the normalized expression matrix subsetted by the highly variable genes using the function `sc.tl.pca()`. A cell-cell neighborhood graph was computed based on the first 30 principal components using sc.pp.neighbors, followed by Leiden clustering (resolution = 1.0), resulting in 43 clusters. For data visualization purposes, the cell neighborhood graph was projected into two dimensions using UMAP with the function `sc.tl.umap()`. We assigned cell lineage annotations to each of these clusters based on gene expression patterns in a panel of canonical lineage-specific marker genes.

### Analysis of publicly available colorectal cancer single-cell datasets

We collected the Chen *et al*. 2024 dataset^25^ for integrative analysis with our own dataset. We downloaded the gene expression counts matrix and patient-level metadata from Gene Expression Omnibus (GEO) accession GSE236581. This dataset includes data from Primary CRC tissue samples, adjacent healthy tissue samples, and peripheral blood mononuclear cells (PBMCs), but for the purpose of this analysis we only include cells from the primary CRC tumor (279,886 cells). This expression matrix was already filtered using the same UMI, genes, and mitochondrial read thresholds as our dataset, and similarly we applied Scrublet to detect barcode collisions in this dataset. We applied a basic SCANPY-based clustering analysis to this dataset, similar to the analysis that we performed in our own dataset. We used the broad cell lineage labels provided by the original Chen *et al*. 2024 dataset to subset cells for lineage-specific analyses.

### Subclustering and annotating the T cell lineage

We performed a secondary clustering analysis in order to identify fine-grained cell populations of the T cell lineage. In this analysis, we integrated T cells from all metastatic tumors samples in our dataset, and with the primary tumor dataset from Chen *et al*. 2024. We first created a merged anndata object of all T-cell lineage cells from the two datasets, with 390,892 cells in total. We applied a dataset-wide filter to remove cells with greater than 5% mitochondrial reads, and we removed genes that were expressed in fewer than 3 cells, retaining 363,792 cells and 30,548 genes. When integrating single-cell datasets, it is essential to strike a balance between correcting batch effects and retaining biological variation^40^. With this in mind, we sought to define a set of features that would accurately represent the spectrum of T cell populations present in these distinct tissue contexts. For each combination of tissue + timepoint (i.e., liver metastasis at the screening timepoint) in our dataset, we performed highly variable feature selection using the Python package BigSur^41^. This approach calculates modified corrected Fano factors and their p-values for each gene in a UMI counts matrix, providing a meaningful ranking for each gene and identifying sets of genes that are significantly variable within a dataset, in contrast to more standard feature selection approaches in SCANPY and Seurat which rely on an arbitrarily selected number of features^42^. For downstream clustering analysis, we retained all genes that were considered highly variable by the BigSur algorithm in three or more tissue + timepoint combinations (2,130 genes). We performed linear dimensionality reduction, batch correction, and biological signal recovery using Harmony^43^ (harmonypy Python package version 0.0.10) and CellANOVA (version 0.1.0). For each tissue, we defined the pre-treatment samples as the “pool of controls” for CellANOVA, and the post-treatment samples as the condition, in order to construct a cellular embedding that retains the biological signal related to ICI treatment. CellANOVA outputs an adjusted gene expression matrix for the given set of cells and variable genes, and we used this CellANOVA-corrected matrix for PCA and Leiden clustering within SCANPY. Several Leiden clustering resolutions between 1 and 3 were tested, and we moved forward with resolution = 3 for downstream annotations (43 Leiden clusters). We projected the data into two dimensions using UMAP with the function `sc.tl.umap()`for data visualization.

We annotated these clusters using both a panel of canonical T cell marker genes, and data-driven marker gene discovery in each cluster. Cluster-specific marker genes were identified using a two-sided Wilcoxon rank sum test via the SCANPY function `sc.tl.rank_genes_groups()` and we focused on marker genes with FDR-corrected p-values ≤ 0.05 and a log2(fold change) ≥ 1.0. Furthermore, we inspected the distributions of quality-control metrics (number of UMI detected per cell, number of genes detected per cell, percentage of mitochondrial reads, and doublet scores from Scrublet^37^) in each cluster to identify additional outliers. In total we removed 5 out of 43 clusters due to outlier status regarding quality-control metrics, or evidence of potential doublets like contradictory gene expression patterns. The remaining clusters were annotated into 24 functional T cell categories based on marker gene expression patterns: CD4+ central memory, CD4+ follicular helper, CD8+ resident, CD8+ activated, CD8+ effector, CD8+ activated, CD8+ metabolic, CD8+ interferon, CD8+ pre-exhausted, CD8+ exhausted, regulatory T cells (Tregs), naive T cells, proliferating T cells, natural killer (NK) cells, mucosal-associated invariant T (MAIT) cells, and gamma-delta T cells. The original numeric cluster identities were also retained to add further granularity within these T cell populations throughout our analysis when appropriate.

### Integrated subclustering of each cell lineage

Similar to the T cell lineage, we performed secondary integrated clustering analyses on the following cell compartments in our dataset: myeloid cells, B and plasma cells, stromal cells, and tumor cells. We followed the same analytical pipeline as previously described for the T cell compartment to integrate the data from each of these compartments with the corresponding data from the Chen *et al*. 2024 primary tumor dataset. For all cell compartments except the tumor compartment, we removed cells with greater than 5% mitochondrial reads. In each analysis, we removed additional low-quality Leiden clusters indicated by doublet scores, QC metrics, and conflicting marker genes. Except in the tumor compartment, Leiden clusters were annotated by inspecting the gene expression distributions of canonical marker genes, and via unbiased cluster marker gene analysis (two-sided Wilcoxon rank sum test, **Tables S3-S6**).

For the myeloid compartment, we retained 37,229 cells after QC filtering (56,534 cells initially), grouped into 20 Leiden clusters (resolution = 1.0). Based on marker gene expression, these clusters were broadly annotated into the following functional categories: classical monocytes, non-classical monocytes, resident macrophages, tumor-associated macrophages (TAMs), lipid-associated macrophages (LAMs), and dendritic cells (DCs). For the B cell compartment, we retained 67,043 cells after QC filtering (83,302 cells initially), grouped into 15 Leiden clusters (resolution = 1.0). We annotated four B-cell populations (activated, memory, transitional, and memory), and five plasma cell populations (long non-coding RNA high plasma cells, mitochondria-high plasma cells, IGHA2 plasma cells, IGHG1 plasma cells, and other plasma cells). For the stromal compartment, we retained 24,215 cells after QC filtering (32,689 cells initially), grouped into 18 Leiden clusters (resolution = 1.0), which were coarsely categorized into endothelial cells, fibroblasts, and pericytes. For the tumor epithelial cell compartment, we performed subclustering analysis only using the cells from the liver metastases, lung metastases, and the primary tumor dataset. After filtering for these tissues, and after further QC, we retained a total of 113,646 cells (168,835 cells initially across all tissues), grouped into 23 Leiden clusters (resolution = 1.0). Due to the inherently heterogeneous nature of tumor cells, for this compartment we did not provide further annotations beyond the Leiden cluster identities, rather we sought to characterize the functional states of these cells with cell-level signature scoring of 14 canonical cancer pathways from the CancerSEA^30^ database (R package version 0.1.0) using the UCell^44^ algorithm (R package version 2.10.1).

### Differential cell abundance analysis

We compared the relative abundance of cell states between clinical groups using Milo^23^ (R package version 2.0.0). This method first aggregates transcriptomically similar cells into “cell neighborhoods”, based on a *k*-nearest neighbors (KNN) graph computed on a low-dimensional embedding of the transcriptomic data; here we used the 30-dimensional embedding from CellANOVA^22^ with Harmony^43^. Differential abundance within cell neighborhoods was modeled using a negative binomial generalized linear model implemented via edgeR^45^ (R package version 4.2.1). Differential cell abundance analysis was performed separately for each of the three immune cell compartments in our dataset: T cells, myeloid cells, and B / plasma cells. In our dataset, we compared cell abundances post- versus pre-treatment for liver and lung metastases separately. In the external primary CRC tumor dataset, we compared the second timepoint to the baseline timepoint, and excluded the additional timepoints.

### Cross-lineage immune coordination networks

To identify synchronized changes in cell abundance across the distinct lineages, we performed a cross-lineage co-abundance network analysis. First, we calculated the relative proportion of each cell state for every patient both at the pre-treatment (SCR) and post-treatment (C02) timepoints. To account for the compositional nature of single-cell data, the raw proportions were transformed using the centered-log-ratio (CLR). We then calculated the treatment-induced shift (ΔCLR) for each cluster by subtracting the baseline CLR from the post-treatment value. We performed a pairwise Pearson correlation of the ΔCLR values for each immune cluster across all patients, stratified by tissue (liver metastasis, lung metastasis, and primary tumor). These Pearson correlation coefficients form the basis of the cross-lineage immune coordination networks. Hierarchical clustering was performed on the raw Pearson correlation matrix, to identify concerted groups of immune cells with shared dynamics, and we visualized the hierarchical clustering results using the ComplexHeatmap R package (version 2.22.0). We performed a permutation test (*n*=1,000) by randomly shuffling patient identifiers, allowing us to assess the significance of these connections by comparing to the null distribution of correlation coefficients. Co-abundance networks were constructed where nodes represent individual immune clusters, and edges represent shared treatment-induced dynamics (*pperm* < 0.05 and R > 0.6). Network topology metrics, including degree centrality, betweenness centrality, and eigenvector centrality, were calculated to identify central “hub” immune subtypes driving the coordinated response. All network analyses were implemented using the igraph (version 2.1.4), tidygraph (version 1.3.1), and ggraph (version 2.2.2) packages in R.

### T-cell consensus gene expression programs

To complement the marker-gene based annotations attributed to cell clusters in the T-cell compartment, we investigated a set of T-cell consensus gene expression programs (cGEPs) from the T-cell annotator (TCAT) tool. Cell-level TCAT scores were calculated using the starCAT^20^ Python package (version 0.0.0), and pseudo-bulk level scores were generated via decoupleR ULM. We correlated the pseudo-bulk gene expression signatures between our co-expression modules and each of the TCAT cGEPs. Treatment-associated changes in cGEP activity were estimated per T-cell populations and metastatic site using a paired limma^46^ model (∼ patient + timepoint + log10(nCells)) on the pseudobulk program scores. Given the limited statistical power of this dataset after grouping by pseudobulk samples across cell populations, this comparison was designed to be descriptive, and we report effect sizes (model coefficients) rather than significance levels.

### High-dimensional weighted gene co-expression network analysis

We performed high-dimensional weighted gene co-expression network analysis (hdWGCNA) within the integrated T cell lineage and tumor / epithelial cell datasets (cells from metastatic tumor samples + primary tumor samples from Chen *et al*. 2024) using the R package hdWGCNA^27^ (version 0.4.7; WGCNA version 1.73). For this analysis, we performed network analysis separately for key tissue + treatment timepoint combinations: liver metastasis pre-treatment, lung metastasis pre-treatment, liver metastasis post-treatment, lung metastasis post-treatment. We also ran the same network analysis pipeline on the Chen *et al.* 2024 dataset (primary tumor pre-treatment and post-treatment), resulting in six co-expression networks in total for the T-cell lineage. Each of the individual network analyses are comprised of the following steps: i) subset the dataset to the group of interest (i.e., treatment timepoint + tissue), ii) filter out genes expressed in less than 5% of cells, iii) aggregate cells into metacells, iv) perform a parameter sweep for suitable soft-power threshold values, v) construct the weighted gene co-expression network, and vi) apply hierarchical clustering and dynamic tree cutting^47^ (R package version 1.63.1) to the co-expression network to group genes into co-expression modules. We calculated cell-level gene expression scores (module eigengenes, MEs) for each of these gene modules using the hdWGCNA function `ModuleEigengenes()`, and we calculated the eigengene-based connectivity (kME) of each gene using the hdWGCNA function `ModuleConnectivity()`. To ensure consistency and reproducibility across these different network analyses, we implemented this procedure as a Snakemake^48^ pipeline. Furthermore, we also correlated our gene modules with the TCAT Antigen-Specific Activation (ASA) score. To establish the relationship between each module and the T-cell populations, we first visualized the cell-level gene expression (MEs from hdWGCNA) for each population as a dot plot. We next performed a one-versus-all “marker module” test using Seurat’s FindAllMarkers function, employing the MAST^49^ model on cell-level MEs to identify which T-cell clusters significantly expressed each module at a higher level relative to the rest of the T-cell population. We also used the same analytical pipeline to perform a separate co-expression network analysis on the malignant fraction.

### T-cell gene co-expression module similarity networks

To directly compare the T-cell gene co-expression modules identified across these six independent networks, we constructed a module-module similarity network. We first quantified the gene set overlap between all pairs of modules using the GeneOverlap R package (version 1.42.0), with significance determined by Fisher’s exact test. Critically, to focus on the most functionally important genes in these gene overlap tests, we only considered the top 100 genes in each module, ordered by intramodular connectivity strength (kME). We calculated pseudo-bulk co-expression module activity scores (patient x timepoint x T-cell cluster) using the univariate linear model (ULM) framework in the R package decoupleR (version 2.12.0), and we calculated pairwise Pearson correlations for these pseudo-bulk expression scores. A composite network edge weight was derived by averaging the Jaccard index of the gene-set overlap, and the Pearson correlation coefficient (50/50 weighting, after 0-1 scaling). The final module similarity network, as shown in Fig. 3a, was assembled by retaining the top 10% of network edges, where nodes represent individual co-expression modules and edges represent shared biological identity (gene set similarity) and expression patterns (pseudo-bulk ULM score correlations).

### Assessing the topological preservation of T-cell co-expression modules

To assess the degree of network-level remodeling induced by the treatment, we performed a series of topological comparisons and module preservation analysis using the NetRep^29^ (version 1.2.7) framework. Specifically, we tested whether the network topology of co-expression modules identified at the baseline state before administration of dual-ICI was significantly preserved in the post-treatment state, and vice versa, for liver and lung metastases separately. NetRep utilizes a permutation-based approach (n=2,500) to determine whether the observed topological properties of the “discovery” network modules are more similar to the “test” network than random chance, via the null distribution generated by permuting the network labels. We evaluated a set of seven key module preservation statistics: i) ‘cor.cor’, which measures the concordance of the correlation structure; ii) ‘avg.cor’, the average magnitude of correlation coefficients; iii) ‘avg.weight’, the average magnitude of network edge weights; iv) ‘cor.degree’, the concordance of the scaled weighted degree (identifying if the most strongly connected ‘hub’ nodes remain central in the test dataset); v) ‘cor.contrib’, the concordance of node contribution to the module expression profile (pseudo-bulk decoupleR ULM score); vi) ‘avg.contrib’, the average magnitude of node contributions; and vii) ‘coherence’, the proportion of variance explained by the module summary profile in the test dataset. Modules were categorized into preservation classes based on the number and identity of significant permutation p-values (threshold < 0.05): strong, all 7 metrics significant; moderate, four or greater metrics significant, including the essential structural metrics ‘coherence’ and ‘avg.weight’; weak, at most one significant metric, or insignificance in one of the aforementioned essential metrics. To provide a global measure of preservation strength, we calculated a Z-summary score for each module. For each of the seven statistics, a Z-score was calculated as the observed value subtracted by the mean of the null distribution, divided by the standard deviation of the null distribution. The final Z-summary is an average Z-score across each of the seven metrics, providing a global effect size for how significantly a module’s topology deviates from the null expectation.

### An automated intelligence system to summarize the biological context of gene co-expression modules

There exist many data-driven approaches to group genes together based on similarity in expression patterns across a given set of observations, for example including gene co-expression modules from hdWGCNA^27^ as we present in this manuscript or alternative approaches like non-negative matrix factorization. These methods are generally motivated by the systemic coordination of individual genes into functional biological pathways, wherein a gene co-expression module serves as a model of a pathway or even a set of pathways which have similar expression patterns across the given set of observations. However, the biological identity of these modules is not immediately clear after obtaining these results, and researchers typically perform a suite of downstream analyses to build a biological understanding of each module. For example, a typical way that researchers annotate co-expression modules is by manually inspecting the set of hub genes in each module, or perhaps performing gene ontology enrichment analysis of these gene sets. The module annotation process may also require more in-depth analysis like differential expression of each module across a given set of experimental conditions. Ultimately this process may be different for each dataset depending on its inherent qualities and the goals of the study. To date this is largely a manual process that requires careful analysis and curation. To aid in the automated annotation of the gene co-expression modules, we developed the open-source software package **llegir** (**LL**M-**e**nabled **g**ene module **i**nterpretations in **R**; https://github.com/smorabit/llegir).

For a given dataset and a set of modules, llegir first constructs a `ModuleSet`, a lightweight adapter containing gene-module membership, summarized expression levels of the module (e.g., module eigengenes, decoupleR scores, UCell scores), and relevant meta-data via a uniform interface agnostic of the algorithm used to identify the modules (hdWGCNA, cNMF, literature-derived gene lists, etc). The user also supplies a brief text description of the dataset to provide necessary context to frame the biological setting (species, tissue, cell compartment, assay, experimental conditions, additional notes). An *evidence packet* is then assembled for each module by running a uniform suite of analysis tools: identifying hub genes per module, gene-set overlap enrichment with databases such as Gene Ontology or MSigDB, module-level and gene-level differential expression across conditions, module-level expression with respect to different cell populations. Individual tools produce *evidence fragments*, which are uniformly formatted via a JSON schema, enabling a smooth handoff to an LLM for summarizing the whole evidence packet per module. Further, since llegir organizes these tools into a registry, users can add their own tools for custom analyses, which in turn could be easily shared among other users of the package. Critically, these evidence fragments can be collected at several resolutions within one evidence packet (cell-level, sample-level, or pseudobulk-level).

To interpret a module, llegir assembles an LLM prompt based on a template prompt along with the top findings from each evidence fragment, including key statistics like effect sizes and significance levels where applicable. The LLM itself is accessed through the `ellmer` R package, which lets the user choose any supported LLM provider and model, including open-source models running on local hardware. Rather than free-writing prose, the model then fills in a structured *interpretation object* with typed fields: a proposed label, one-line summary, dominant biology, supporting claims (each citing the specific fragments that back it, with direction and strength), and, where supported, cell state, condition dynamics, metadata associations, plus a confidence value and review flags. A human-readable summary is produced by a deterministic, versioned template with no further model call. This process is repeated separately for each module, where the LLM’s context is fully refreshed in between calls to reduce potential confusion. For joint review across all modules, llegir compiles a single self-contained HTML report pairing the LLM-drafted narrative for each module with its underlying evidence packet, so the user may read the interpretation side-by-side with the metrics which support it.

A central challenge in applying LLMs to this kind of analysis is the tension between the flexibility of an autonomous, agentic system and the rigor and reproducibility expected of a standard bioinformatics method. To reconcile the two, llegir enforces that every claim in a module interpretation references evidence fragments that exist in that module’s evidence packet. Because the evidence packets, prompt templates, and tool and software versions are all hashed and recorded in a provenance log, any given interpretation can be traced back to the exact evidence and package state that produced it, even though the LLM’s wording is not itself deterministic. Above all, llegir is built around a human-in-the-loop philosophy: it is not meant to be a fully autonomous annotator, but rather to accelerate expert curation by drafting interpretations and flagging the modules (insufficient evidence, conflicting tools, likely artifacts, or low-specificity labels) that most warrant a closer look. Finally, for users who wish to explore beyond this bounded workflow, llegir offers an “agentic mode”, which exports completed run to a self-contained workspace where an external coding agent can be launched to facilitate further analysis and interrogation of the data, in addition to module annotations similar to the stand. In this manuscript, co-expression modules in the T-cell lineage were annotated using the standard llegir approach with the open-weights Qwen2.5-7B-Instruct as the underlying model, while the tumor modules were annotated using the llegir agentic mode with Anthropic’s Claude Opus 5 as the underlying model.

### Single cells V(D)J-T assignment

High-level V(D)J annotations of high-confidence, cell-associated productive contigs from Cell Ranger’s (version 7.0.0) *filtered_contig_annotations.csv* output from 59 V(D)J-T libraries (24 patients; SCR, CO2 and EOT time points) were used to perform per-cell V(D)J chain assignment, yielding the complementarity-determining region 3 (CDR3) V(D)JC gene, amino acid and nucleotide sequences of the TCR α and β chains associated with each cell barcode. This was achieved individually for each library using *scRepertoire’s*^50,51^ *CombineTCR* function (version 2.9.0). Cells with single or paired α and β chains assigned were considered (*remove.na = FALSE*); for cells with more than one α or β chain assigned, the chain with highest UMI counts was selected to mitigate potential multiplets and technical artifacts (*remove.multi = FALSE, filter.multi = TRUE*). This resulted in a total of 208,530 TME T cells for which TCR V(D)J data were recovered, and 203,380 cells from which β chain contigs were assigned.

### Clonotypes definition

Clonotypes were defined on the basis of the β chain CDR3 junctional nucleotide sequence in combination with the associated V and J gene segments. This allowed for maximal cell recovery given higher expression of β chains (compared to α; **Figure S22A**), maximal antigen specificity, preservation of rearrangement origin enabling clonotype tracking, and further integration with matched peripheral blood bulk β chain V(D)J profiles (OS-TCR, Omniscope Inc.). T cells with identical clonotypes were grouped into clones, from which clonal frequencies and proportions (frequencies normalized by sample size) were quantified using *scRepertoire’s clonalBin* function at the sample (patient + timepoint) level; technical replicates were aggregated to reflect the actual sample composition. This captured a total of 95,617 tumor clonotypes (**Figure S22B**). Clonotypes were classified based on their clonal sizes: singletons were defined as clonotypes observed exactly once (clonal frequency = 1). All non-singleton clonotypes were considered expanded and were subcategorized based on their relative frequency within the repertoire into: rare (X <= 1e-04), small (1e-04 < X <= 1e-03), medium (1e-03 < X <= 1e-02), large (1e-02 < X <= 1e-01), and hyperexpanded (1e-01 < X <= 1) sizes. Clonotypes were further classified based on their longitudinal presence status into non-persistent clonotypes: detected only before treatment (SCR samples); *de novo* clonotypes: detected only after treatment (C02 samples); and persistent clonotypes: detected both before and after treatment.

### Matching TCRs with GEX profiles

V(D)J-T-library T cells were matched with high-quality preprocessed GEX-library annotated T cells by their cell barcodes to enable joint T-cell clonal expansion and functional characterization (171,788 out of 206,945 cells from CD45+ GEX libraries; 83,01% of GEX T cells had productive TCRs; **Figure S22C**). This was performed on the T-cell/NK compartment and filtering out clusters annotated as NK, to restrict the overlap to TCR+ T-cell populations. First-level (lv1) annotations (CD4+, CD8+; T Naïve/CM, Tregs, non-Treg effector CD4+ T cells and effector CD8+ T cells) and second-level (lv2) annotations representing higher resolution T-cell states and phenotypes were used for downstream analyses. T cells were classified into CD4+ and CD8+ lineages using their lv1 and lv2 annotations. Because the Naïve and Proliferating clusters contained a mixture of both lineages, cells in these clusters were instead assigned based on log-normalized expression of *CD4* relative to *CD8A* and *CD8B*. For clonotype-level analyses, the most abundant phenotype within each clonal population was selected, in contrast to cell-level analyses, where each TCR-GEX matching T cell retains its associated phenotype. T-cell phenotype (lv2) transitions between SCR and C02 time points were visualized through alluvial plots using the *ggalluvial* R package (version 0.12.6).

### Cohort-wide tumor TCR repertoire quality and diversity assessment

Diversity metrics are inherently sensitive to sample sizes and experimental technical factors, especially for richness-weighted indices. Because larger samples recover a greater proportion of the low-frequency rare clonotype tail^52^, conventional metrics that incorporate or heavily weigh richness are artificially inflated independent of the true underlying diversity^53^. Although all libraries in this study were sequenced at comparable depth, the number of T cells and clonotypes recovered per sample varied substantially, reflecting both technical variability (i.e. CD45+ cell sorting, sample quality) and potentially biological variability in T-cell abundance across patients and timepoints. The oligoclonal structure across the full SERPENTINE cohort was first described, dominated by a small fraction of expanded clonotypes with a long tail of low-frequency clonotypes (**Figure S22D**). We then quantified per-sample Hill numbers of order q=0 (richness), 1 (Shannon diversity), 2 (Inverse Simpson Index), to investigate the association between diversity and sample size as a function of q (increasing q placing greater weight on clonotype evenness and reduced sensitivity to rare clonotypes). These were computed using the scRepertoire’s *clonalDiversity* function with *skip.boots* = *TRUE* such that downsampling was not done. Consistent with this expected bias, raw Hill number diversity estimates of q=1 and 2 correlated with the number of cells recovered per sample (Spearman correlation; **Figure S22E**), confirming that uncorrected diversity comparisons would be confounded by differences in sampling depth rather than reflecting clonal structure alone.

To address this, downstream diversity comparisons (reported in subsequent analyses) standardize samples to a common sequencing depth via bootstrapped downsampling, isolating differences in clonal diversity from differences in cells recovered. To evaluate robustness of this approach, sample-level rarefaction and extrapolation curves were generated using the *clonalRarefaction* function from scRepertoire, which implements the Hill-number framework of Chao et al. (2014)^54^ via the iNEXT^55^ package (version 3.0.2), computed at q=1 with 95% bootstrap confidence intervals (*n.boots* = 20, default). For most samples, confidence intervals remained narrow (stable, low-variance estimates) at the observed depth even without full saturation, supporting the reliability of bootstrapped downsampling for downstream comparisons (**Figure S22F**).

### Study cohort subsetting for TCR analyses

To specifically interrogate immune remodeling induced by dual ICI in MSS CRC, and to directly compare clonal dynamics between liver and lung metastases, all downstream TCR repertoire analyses were restricted to MSS CRC patients with either matched SCR and C02 liver or lung metastatic tumor biopsies. This selection strategy minimized confounding effects arising from MSI, involvement of other metastatic sites, and differences in cancer origin by excluding the EC histology. The resulting subset comprised 13 patients (6 liver: P01, P02, P03, P10, P14, P33; 7 lung metastases: P17, P20, P26, P29, P31, P34, P35), each profiled at SCR and C02. In total, 115,710 TCR+ T cells, corresponding to 58,087 clonotypes, were recovered across both time points. Of these, 101,449 TCR+ T cells (87,68%) were successfully matched to high-quality GEX profiles, enabling integrated analysis of clonal architecture and transcriptional cell states.

Clonal scatters (scatterplots) comparing clonal proportions at SCR and CO2 timepoints were used to visualize longitudinal clonal dynamics (stratified by metastatic site and T-cell lineage). Clonal proportions were log₁₀-transformed, with a pseudocount (ε = 1×10⁻⁴) added to accommodate the highly skewed distribution characteristic of single-cell TCR data. To specifically investigate clonal dynamics of persistent clones, two-dimensional kernel density estimates were computed to visualize shifts in clonal occupancy (*geom_density_2d_filled* with *contour_var = “ndensity”*), informing clonal expansion and contraction shifts. Additionally, stacked barplots were used to visualize proportions of clonal classes per Lv2 annotations and presence status groups at the TCR+ cell level. For these, one outlier clonotype (TRBV2_TRBJ1-1_GCCAGGGGGGATGATCCGAAGGGGCGGGGGACTGAAGCTTTC) was excluded from patient P10 at SCR, as it accounted for a disproportionate clonal frequency (n = 2,969 cells; 31.8% of the sample’s repertoire) that would have masked actual clonal dynamics and overloaded the hyperexpanded clonal class.

### Longitudinal diversity assessment of tumor TCR repertoires

Building on the diversity bias assessment described above, we performed a longitudinal comparison of tumor TCR repertoire diversity between SCR and C02 timepoints across multiple levels of resolution: the global TCR+ repertoire, the lineage level (CD4+ and CD8+ repertoires), and the sublineage level (effector non-Treg CD4+, Treg, and effector CD8+ repertoires). We additionally assessed diversity among CD8+ timepoint-unique clones (non-persistent vs. *de novo*), as well as among persistent clones alone, comparing diversity between SCR and C02 in each case. To avoid biasing our interpretation of diversity dynamics toward any single metric, we evaluated multiple diversity indices for each comparison available under the scRepertoire ecosystem:

– Shannon entropy: quantifies the joint richness and evenness of clonotypes, providing an overall measure of diversity that is more sensitive to rare clonotypes than dominance-based indices. Higher values indicate the presence of more distinct clonotypes and a more even clonal distribution [0, ∞].
– Inverse Simpson index: represents diversity weighted towards dominant clones and is comparatively less sensitive to rare species. Low values indicate dominance by one or few highly expanded clones, while high values suggest that diversity is spread across multiple equally-abundant clones [1, ∞].
– D50: number of top clones constituting 50% of the repertoire. Offers a measure of clonal dominance that is less influenced by the long tail of rare clonotypes. Low values indicate that top clones are highly dominant while higher values indicate lower dominance and a more evenly distributed repertoire [1, ∞].
– Pielou’s evenness index: calculated as Shannon entropy normalized by the natural logarithm of the sample’s richness (inverse of clonality). Isolates the evenness component of diversity independent of richness, allowing evenness to be compared directly across samples with differing numbers of unique clonotypes. Values approaching 1 indicating a maximally even clone-size distribution [0, 1].

Diversity metrics were computed using scReperoire’s *clonalDiversity* function at the sample level (patient + timepoint) stratified by metastatic site (liver or lung) with 1,000 bootstrap iterations, downsampling (with replacement) all samples to a common depth, to enable fair comparison of diversity across samples. The downsampling depth was set to the 10th percentile of the corresponding subset’s sample sizes; the minimum sample size was not used given it was consistently driven by a low-number single sample (P03-SCR) so it would increase the probability of losing underlying clonal structure through random subsampling. For each sample, the mean diversity score across bootstrap iterations was used as the point estimate, and 95% confidence intervals were calculated as the 2.5th and 97.5th percentiles of the corresponding bootstrap distribution, providing a non-parametric assessment of the robustness of each sample-level diversity metric. Given the limited statistical power (small sample size), Cohen’s *d* effect sizes were used as a robust estimate of the magnitude of change in diversity across timepoints independent of statistical significance. Cohen’s *d* values were reported for paired SCR versus C02 comparisons, with statistical significance assessed using two-sided Wilcoxon signed-rank tests. Comparisons of diversity between metastatic sites at a given timepoint were assessed using two-sided Wilcoxon rank-sum tests. Inverse Simpson index was selected as the representative diversity metric for its robustness to singleton-driven biases inherent to the small sample sizes and low sequencing depth of single-cell TCR sequencing.

### OS-TCR (Single-molecule TCR sequencing, Omniscope)

The osTCR assay was used to characterise the TCR repertoire of the SERPENTINE trial patients from peripheral blood before treatment and on-treatment timepoints when available. Cryopreserved PBMC were thawed by immediate immersion of the vials in a 37°C water bath, and then washed in complete media. After washing PBMC in PBS, RNA isolation and subsequent concentration were performed using the RNeasy Plus Mini Kit (QIAGEN) and the RNeasy MiniElute Cleanup Kit (QIAGEN). The isolated RNA served as the input material to perform the proprietary Omniscope (Omniscope Inc., Barcelona, Spain) osTCR assay to generate sequencing libraries.

### Peripheral blood deep TCR repertoires

Peripheral blood bulk β OS-TCR sequencing data was available for 32 patients across the entire SERPENTINE trial cohort at multiple timepoints including SCR (31 patients), C01 (10 patients), C02 (24 patients), other specific treatment cycles (C03: 1 patient, C04: 1 patient, C06: 1 patient, C14: 1 patient, LTI: 1 patient) and EOT (5 patients; also referred as DISC for one patient). This comprised a total of 75 libraries consisting of 224,675,952 single molecules and 57,902,418 clonotypes (**Figures S27A and S27B**). Cohort-wide longitudinal clonal dynamics (SCR: pre-treatment vs. C02: post-treatment) were represented through richness-descriptive boxplots and clonal scatters from patients with matched SCR and C02 samples, where clonotypes were grouped into bins to visualize treatment-induced expansion density shifts. Downstream peripheral blood OS-TCR clonal dynamics were restricted to the MSS CRC cohort of patients with either liver or lung metastases and to the SCR and C02 timepoints, in line with the 10x tumor single-cell TCR sequencing data.

Within this subcohort, OS-TCR data from SCR and C02 timepoints were available for 15 patients, including 6 patients with liver metastases and 9 patients without liver metastases but with lung metastases. Importantly, 12 of the 13 patients of the tumor 10x TCR dataset had a complete SCR-C02 match with the blood OS-TCR dataset (patients 01, 02, 03, 05, 10, 14, 20, 26, 29 31, 33, 34, 35; 6 liver and 6 lung); for patient 17, OS-TCR data was only available at SCR. Additionally, SCR and C02 matched OS-TCR data from 3 patients were included for blood-only-based analyses for whom tumor 10x data were not available (patients 15, 19 and 21); For patient 15, OS-TCR data were not available at SCR but at C01 (first treatment cycle); since this sample was collected before immunotherapy administration and is immunologically equivalent to a baseline, it was treated as SCR and included accordingly in all analyses. Overall, the peripheral blood OS-TCR MSS CRC study cohort comprised a total of 15 patients, 30 libraries, 70,912,348 single molecules and 14,389,467 clonotypes. OS-TCR contig files were used as inputs for downstream analyses and clonotypes were defined by identical contig IDs (V gene + J gene + CDR3 nucleotide sequence).

### Peripheral blood T-cell clonal diversity

Longitudinal peripheral blood clonal diversity between SCR and C02 time points was assessed via richness (number of clonotypes), Shannon entropy, Inverse Simpson Index, Pielou’s evenness and D50. These diversity indices were computed using the *diversity* (*index = shannon, invsimpson*) and *specnumber* (for richness) functions from *vegan*’s^56^ R package (version 2.7.5). Pielou’s evenness was computed as the samples’ Shannon entropy divided by the natural logarithm of their richness, and D50 as the minimum number of clonotypes whose cumulative sum of sorted frequencies surpassed 0.5 of the summed library frequency. A bootstrapping approach was not applied given computational constraints inherent to the OS-TCR data dimensions and given the absence of an association between number of cells sequenced and Hill numbers on OS-TCR repertoires used in downstream analyses (**Figures S27C and S27D**), which highlights the capacity of deep TCR sequencing in capturing a good representation of the samples’ clonal structure regardless of sample size. Paired diversity differences between timepoints were statistically assessed via two-sided Wilcoxon signed-rank tests.

### Circulating differentially expanded clonotypes post immunotherapy

To identify differentially expanded (DE) or contracted clonotypes between pre-treatment (SCR) and post-treatment (C02) samples in peripheral blood (OS-TCR), a proprietary noise model (Omniscope Inc., Spain) was applied. The model generates a null distribution for each clonotype to account for sampling and technical variability, with a tunable parameter controlling stringency. Significance (event-relatedness) was determined by the noise model, which computes p-values under the null hypothesis of no change, using an absolute log₂ fold change ≥ 2 on clonal frequencies. Clonotypes uniquely present in one sample were assigned a count of zero in the absent sample. Clonotypes showing statistically significant expansion in the second sample (C02) relative to the first (SCR) were labeled “event-related, expanded”, whereas clonotypes significantly reduced in the second sample were labeled “event-related, contracted”. Event-related clonotypes were further classified as pre-existing if detectable in both samples, or *de novo* if they appeared only in a single sample. The proportion of event-related contracted versus expanded clonotypes among OS-TCR repertoires, and the proportion of each event-related status subgroups (contracted *de novo*, contracted pre-existing, expanded pre-existing, expanded *de novo*) among the event related subset were computed at the patient level and statistically assessed for differences via two-sided Wilcoxon signed-rank tests.

### Clonotype tracking of therapy-specific clones across compartments

Having tumor-and-blood-matched pre- and post-treatment samples enabled tracking of clonotypes across time points and compartments, and the identification of therapy-specific clones: clones that are differentially expanded after treatment in both tumor and peripheral blood. Tumor 10x clonotypes were matched with blood OS-TCR clonotypes by clonotype id (β chain V gene + J gene + CDR3 junctional nucleotide sequence) excluding the first and last codons of 10x clonotypes. This was performed for patients with complete SCR-C02 overlap across compartments (6 liver, 6 lung). Overlap of tumor clonotypes in blood from patients with matched SCR and C02 tumor and blood samples was quantified using raw percentages.

Post-treatment tumor enriched (DE) clonotypes were defined as those exhibiting a log2 fold change ≥ 2 in normalized clonal frequency between post-treatment (C02) and pre-treatment (SCR) samples. For *de novo* clonotypes, which are not detected at baseline, direct fold-change calculations are not possible. To address this, we used the 90th percentile normalized clonal frequency of non-persistent clones from the same patient as a baseline. This baseline value serves as a reference for typical, non-tumor-specific clonal expansion, allowing us to identify *de novo* clonotypes whose expansion exceeds that of bystander clones. Clonotypes meeting this criterion were classified as enriched and retained for subsequent functional analyses, highlighting clones likely expanded in response to therapy rather than reflecting baseline clonal activity. Post-treatment tumor clonotypes were further classified based on their blood event-related status (as described in the *Circulating differentially expanded clonotypes post immunotherapy* section above). Clonotypes were grouped into the following differential expansion status (DE Status) categories: i) therapy-specific clones, DE in both tumor and blood (Tumor and Blood); ii) Only DE in tumor (Tumor-only); iii) Only DE in blood (Blood-only); and iv) not DE in any compartment (None).

### Functional characterization of therapy-specific clones

After classifying tumor post-treatment clones based on their tumor and blood differential expansion statuses, we assessed for T-cell functional differences between the distinct DE status groups to identify metastatic site-specific profiles of therapy-specific clones. We first quantified lv2 annotations of post-treatment TCR+ cells stratified by their clonal DE status groups and metastatic sites, and visualized them through stacked barplots to phenotypically characterise therapy-specific T cells. We then scored post-treatment TCR+ cells based on different T-cell function-related gene set signatures with AUCell using DecoupleR’s^57^ *run_aucell* function (version 2.6.0) on the log-normalized counts, and then averaged them per DE status group at the patient-level. AUCell scores were then scaled into Z-scores across all patients and DE status groups for each signature independently, enabling comparisons of relative signature activity across DE status groups and between metastatic sites. We defined the following gene signatures for scoring:

– Activation: CD27, CD82, TNFRSF4, TNFRSF9, CXCR3, IL2RA, CD69, CD74, IL2RB, ICOS, CD28
– Cytotoxicity: *IFNG, NKG7, PRF1, GZMK, GZMA, GZMB, GZMH, FASLG, KLRG1, TNF, FGFBP2, LAMP1, FAS*
– Residence: *ITGAE, ZNF683, CD44, ITGA1, CD69*
– Tumor reactivity: *41BB, CXCL13, ENTPD1*
– Migration and adherence: *ITGA4, CCR5, ITGAL, CD44, S1PR1, CX3CR1*
– Exhaustion: *PDCD1, HAVCR2, LAG3, TIGIT, CTLA4, CD244, CD160, BTLA*
– Proliferation: *MKI67, TOP2A, PCNA, CDK1, CCNB1, CCNB2, CCNA2, CDC20, CDC25C*
– Stemness and memory: *TCF7, LEF1, IL7R, CCR7, SELL, KLF2, FOXO1*

To further functionally characterize therapy-specific clones, we scored T-cell driven hdWCGNA gene co-expression modules from the Primary, Lung and Liver compartments (see *High-dimensional weighted gene co-expression network analysis* section above) on the different DE status groups. To achieve this, pseudobulks were created by replicate and DE status using scuttle’s^58^ (version 1.20.0) *aggregateAcrossCells* function (*use.assay.type = “counts”, statistics = “sum”*), and further log-normalized using *logNormCounts* function (*use.assay.type = “counts”, pseudo.count = 1*) to correct for library size differences. Modules were ULM-scored using decoupleR’s *run_ulm* function and further scaled (Z-scores) across patients and DE groups. Within-tissue and inter-tissue scores differences across DE status groups were statistically assessed via two-sided Wilcoxon signed-rank and Wilcoxon rank-sum tests, respectively. Additionally, the Δ of liver vs lung Z-Scores across DE status groups and the Δ of therapy-specific (Tumor and Blood) vs not DE (None) TCR+ across metastatic sites were reported as heatmaps for easier interpretation of module expression differences between DE status groups and metastatic sites. P-values from Δs comparisons were corrected for multiple testing via the Bonferroni-Hochberg method when visualized through heatmaps.

### Immune-malignant coordination network analysis

To characterize the functional state of the malignant compartment, we first quantified the activity of 14 canonical cancer gene expression programs from the CancerSEA^30^ database. Single-cell expression scores for these programs were calculated using UCell, while patient-level pseudo-bulk activity scores were calculated via the decoupleR ULM framework. For the simultaneous visualization of multi-program shifts in malignant states, pseudo-bulk CancerSEA scores were min-max scaled (0-1) across the entire cohort to ensure comparability between pathways of different dynamic ranges on the final radar plots. For the pseudo-bulks used in this analysis, we excluded patients with fewer than 75 cells in either lineage, and we only used patients with sufficient cell counts in both the pre- and post-treatment timepoints.

To investigate the cross-lineage relationships in treatment-induced dynamics between the T-cell and tumor compartments, we constructed a cross-lineage module coordination network. We first calculated the treatment-induced shift in module activity (Δ post - pre) for all T-cell and tumor co-expression modules, at the pseudo-bulk patient level. A module-module similarity matrix was constructed by calculating pairwise Pearson correlations of these Δ values across the cohort. Similar to our previous analysis, we constructed a network where edge weights were a composite of the gene-set overlap (Jaccard) and similarity in module activity, however, the critical difference is that here we use the Pearson correlation in the post-treatment minus pre-treatment (Δ) module expression values, rather than the comparison of the static expression values. The strongest 10% of the edges were retained for the final network. We identified 11 clusters, or “meta-modules” (MMs), using the Leiden algorithm (resolution = 2.5). To quantify the degree of cross-lineage integration within these MMs, we calculated the lineage Shannon entropy, where high entropy values indicate high mixing of T-cell and tumor modules within the same meta-module. We also sought to identify which canonical T-cell and tumor expression programs have concordant or discordant dynamics with these data-driven co-expression modules. Therefore, we calculated the Δ expression values of the TCAT cGEPs and CancerSEA signatures at pseudo-bulk level, and correlated these Δ values with the Δ expression values of our co-expression modules.

### Modeling patient survival as a function of CRC tumor gene co-expression modules in TCGA

To facilitate downstream analyses of the malignant gene expression signatures identified in our cohort in a border context, we leveraged bulk RNA-seq datasets from the cancer genome atlas (TCGA)^59,60^. We downloaded processed RNA-seq data of CRC cases within TCGA using the R package TCGAbiolinks (project IDs “TCGA-COAD” and “TCGA-READ”). Since the majority of TCGA CRC RNA-seq samples were from primary tumors, we removed metastatic and recurrent tumor samples before downstream analysis. We also removed 13 formalin-fixed paraffin-embedded (FFPE) samples, yielding 634 bulk RNA-seq samples of primary CRC tumors. Similar to the previously described analysis in pseudobulk scRNA-seq samples, we calculated the gene expression level for each hdWGCNA tumor gene co-expression module using decoupleR ULM scores.

We leveraged extreme gradient boosting trees (XGBoost) to model the patient’s risk of death as a function of these tumor co-expression modules. As a feature selection step to remove the least informative modules, we first fit a simple univariate Cox regression model for each co-expression module, retaining the top 10 modules per network (tissue + timepoint) ordered by ascending p-values (59 / 119 tumor modules retained). We selected the Cox proportional hazards model as the objective function for the XGBoost algorithm, utilizing a negative log-likelihood evaluation metric. To ensure optimal model performance, we performed a random hyperparameter search over 500 iterations, evaluating the model performance calculated as the Harrell’s Concordance index (C-index) in a 5-fold cross-validation scheme. The final model with the best 5-fold cross-validation performance had a testing C-index of 0.629 and a training C-index of 0.841, demonstrating generalizable predictive performance on unseen data. To add interpretability to the underlying predictions of this model, we calculated SHapley Additive exPlanations (SHAP values), revealing an underlying ranking of which co-expression modules were most informative in modeling a patient’s risk of death. Under this setup, a positive SHAP value corresponds to an increased predicted likelihood of death, whereas a negative SHAP value corresponds to an increased predicted likelihood of survival.

We next sought to test whether these co-expression modules could be used to split patients into high and low-risk groups. The relationship between the module’s SHAP value (explainability) and the module’s expression level (decoupleR ULM score) was used to programmatically determine a cutoff to split patients into these two populations by fitting this data to a LOESS regression curve, and using the expression level with the steepest slope on the curve to define the cutoff. We then fit Kaplan-Meier curves using the R packages `survival` and `survminer`, and assessed whether these curves were significantly stratified patients into high and low risk groups using a Log-rank test.

## Data availability

All raw and processed single-cell sequencing data will be made publicly available upon final publication. Additional single-cell datasets from published studies were obtained via the following accessions: the Chen et al 2024^25^ scRNA-seq + scTCR-seq dataset was collected from the Gene Expression Omnibus at accession GSE236581. The cancer genome atlas (TCGA) CRC bulk RNA-seq dataset was collected using the R package TCGAbiolinks.

## Code availability

All data analysis code used for this study is available on GitHub at the following link: https://github.com/Single-Cell-Genomics-Group-CNAG-CRG/SERPENTINE. The open-source llegir software package is available on GitHub at the following link: https://github.com/smorabit/llegir under an MIT license.

## Acknowledgements

We acknowledge AstraZeneca Partners of Choice. SM was supported by a European Molecular Biology Organization (EMBO) postdoctoral fellowship (ALTF-834-2024). GDC was supported by a Joan Oró FI predoctoral fellowship of the Department of Research and Universities of the Generalitat de Catalunya (2026 FI-1 01103) and the co-financing of the European Social Fund Plus. We acknowledge funding from the European Union’s H2020 research and innovation program (grant agreement 848028) and from the European Research Council (ERC) under the European Union’s Horizon 2020 Research and Innovation Program (grant agreement 810287) awarded to H.H. Further support to H.H. was received from the Ministerio de Ciencia e Innovación (MCI) under grant agreements PID2020-115439GB-I00, PID2023-148687OB-I00 and PID2024-161652OB-I00 and PLEC2021-007654, and by ERA-NET Neuron/Ministerio de Ciencia e Innovación (MCI) under grant agreement PCI2022-133012. H.H. has received additional funding from the European Union’s Horizon Europe Research and Innovation program (grant agreement 101072891) and from LaCaixa Foundation (grants HR22-0031, HR22-0172 and HR25-00861). We are also thankful to the Generalitat de Catalunya through the Suport Grups de Recerca AGAUR (2021-SGR2021-01216 to H.H.). This work was also supported by an ASPIRE Award from The Mark Foundation for Cancer Research and the Scientific Foundation of the Spanish Association Against Cancer (CATALYST 0000000014). AGros was supported from Comprehensive Program of Cancer Immunotherapy and Immunology II (CAIMI-II) (53/2021) supported by the BBVA Foundation, Instituto de Salud Carlos III (MS15/00058, PI17/01085), Asociación Española Contra el Cáncer (AECC) (IDEAS197PORT), La Fundació la Marató de TV3 (201919-30), and Ministerio de Ciencia, Innovación y Universidades (PID2020-118529RB-I00). RPL is supported by the Miguel Servet Programme (ISCIII), co-funded by the European Union (grant CP25/00092), Asociacion Española Contra el Cancer (PRYCO211023SERR) and FERO Foundation. NS would like to acknowledge ESMO for supporting her fellowship at IFOM. Any views, opinions, findings, conclusions, or recommendations expressed in this material are those solely of the authors and do not necessarily reflect those of ESMO.

## Competing interests

**IB** reports accommodation and travel expenses from Amgen, Merck, Sanofi, and Servier; and personal financial interests as Honoraria, Consulting or Advisory Role from Agenus, Amgen, AstraZeneca, Sai-Med and Servier. **HH** is co-founder and Chief Scientific Officer of Omniscope, Founder of Codex Insights, scientific advisory board member of Nanostring, Bruker and MiRXES, consultant to Moderna and Singularity and received an honorarium from Genentech. **EE** reports personal financial interests as Honoraria, Consulting or Advisory Role, and Speakers’s Bureau: Agenus, Amgen, Bayer, Bristol Myers Squibb, Boehringer Ingelheim, Cure Teq AG, GlaxoSmithKline, Hoffman La Roche, Janssen, Johnson&Johnson, Lilly, Medscape, Merck Serono, MSD, Nordic Group BV, Novartis, Organon, Pfizer, Pierre Fabre, Repare Therapeutics Inc., RIN Institute Inc., Rottapharm Biotech, Sanofi, Seagen International GmbH, Servier, and Takeda. **JCN** is a scientific consultant to Omniscope and scientific advisory board member of Biocompare. **JT** reports personal financial interest in form of scientific consultancy role for Accent Therapeutics, Alentis Therapeutics, AstraZeneca, Boehringer Ingelheim, Bristol Myers Squibb, Carina Biotech, Cartography Biosciences, Chugai, Daiichi Sankyo, F. Hoffmann-La Roche, Genentech, Johnson & Johnson/Janssen, Larkspur Biosciences, Lilly, Marengo Therapeutics, Menarini, Merus, MSD, Novartis, Ono Pharma USA, Peptomyc, Pfizer, Pierre Fabre, Quantro Therapeutics, Scandion Oncology, Scorpion Therapeutics, Servier, Sotio Biotech, Syntelios AG, Taiho, Takeda Oncology and Tolremo Therapeutics. Stocks: Alentis Therapeutics, Oniria Therapeutics, 1TRIALSP and Pangaea Oncology. **AGros** received funding from Merck for the submitted work. In addition, A. Gros has a patent for E-059-2013/0 licensed and with royalties paid from Intima Bioscience Inc., Intellia Therapeutics, Inc., Tailored Therapeutics, LLC, Cellular Biomedicine Group, Inc., and Geneius Biotechnology, Inc.; a patent for E-085-2013/0 licensed and with royalties paid from Intima Bioscience Inc., Intellia Therapeutics, Inc., and Geneius Biotechnology, Inc.; and a patent for E-149-2015/0 licensed and with royalties paid from Intima Bioscience Inc., Intellia Therapeutics, Inc., and Tailored Therapeutics, LLC. **AGarcia** reports accommodation and travel expenses from Merck. **GV** has received speaker’s fee from MSD, Pfizer, GSK, PharmaMar, Giliead, AstraZeneca and Pierre Fabre, and received consultant fees from Reveal Genomics. **FS** reports personal financial interests, honoraria for advisory role, travel grants, research grants (past 5 years) from Sanofi Aventis, Amgen, Merck Serono, Servier, Bristol-Myers Squibb, Terumo, MSD, Takeda. **JR** reports personal speaker honoraria from Sanofi, Pfizer, Takeda, BMS, Merck and Amgen; and accommodation expenses from Pierre Fabre, Servier, Takeda, Amgen, and Merck. **NS** reports travel and accommodation expenses from Merck and Amgen. Speakers’ Bureau: From AMGEN and Medistream OncoBites. **RPL** reports receiving research funding from AstraZeneca PLC (global) and Roche Farma S.A. (Spain). RPL also declares a consulting/advisory role with AstraZeneca PLC (global). Additionally, she holds non-remunerated positions as a member of the RECIST Working Group; head of the Cancer Core Europe Imaging Task Force; head of Oncological Imaging for the Spanish Society of Medical Imaging (SERAM); and head of the Research Committee of the European Society of Radiology (ESR). **AR**, **JZM**, **AL**, **NMM** and **PCH** are employees of AstraZeneca and have equity in AstraZeneca. **JLM**, **MS**, and **MG** are employees of Omniscope.

## Author contributions

EE., JT., IB., AR., JZM., JCN and HH. conceived, designed the study and provided biological and translational insights. AR., AL and JZM., provided biological and translational insights. EE., IB., AG., AG., JR., FS., NSG., AG., MB., GC., were involved in sample collection and data acquisition. SM., GDC., GC., PN., DM., JLM., MS., MG., performed experiments, analysis and interpretation of sequencing data. IB., NMM., PCH were involved in the project and study management. SM., GDC., and GV, performed the data and statistical analyses. RPL., and NR performed the imaging and radiological analyses. SM., GDC., GC., PN., DM., JLM., performed the bioinformatic analyses of the single cell data and other computational analyses. SM., GDC., JCN and HH created the main and supplementary figures. IB., SM., GDC., AR., JC., JCN., EE and HH., wrote the manuscript, and IB., SM., GDC., AR., JC., JT., JCN., EE and HH participated in the review and editing.

## Notes

### Clinical Trial

NCT06680739

### Author Declarations

The Hospital Universitari Vall d'Hebron Research Ethics Committee gave ethical approval for this work (approval number: PR(AG)144/201).

