## Supplementary Material for "Site-specific immune remodeling underlies resistance to dual immune checkpoint blockade in metastatic colorectal cancer"

| | Cohort 1a | Cohort 1b | Cohort 2 | |
|---|---|---|---|---|
| | dMMR/MSI | | pMMR/MSS | |
| | Durvalumab | Durvalumab + Tremelimumab | Durvalumab + Tremelimumab | Durvalumab + Tremelimumab |
| | | | **Liver metastases** | **Non active liver mestastases** |
| | n = 3 | n = 3 | n= 6 | n= 18 |
| | Durvalumab | Durvalumab + Tremelimumab | Durvalumab + Tremelimumab | Durvalumab + Tremelimumab |
| **Sex, n (%)** | | | | |
| Female | 0 (0%) | 2 (66.7%) | 3 (50%) | 8 (44.4%) |
| Male | 3 (100%) | 1 (33.3%) | 3 (50%) | 10 (55.6%) |
| **Age** | | | | |
| Median (range) | 55 (36 - 69) | 60 (35 - 83) | 56 (51 - 63) | 60 (48 - 77) |
| **Ethnicity, n (%)** | | | | |
| White | 2 (66.7%) | 3 (100%) | 6 (100%) | 17 (94.4%) |
| Others | 1 (33.3%) | 0 (0%) | 0 (0%) | 1 (5.6%) |
| **ECOG, n (%)** | | | | |
| 0 | 3 (100%) | 1 (33.3%) | 3 (50%) | 10 (55.6%) |
| 1 | 0 (0%) | 2 (66.7%) | 3 (50%) | 8 (44.4%) |
| **Primary tumor location, n (%)** | | | | |
| Right colon | 2 (66.7%) | 2 (66.7%) | 1 (16.7%) | 8 (44.5%) |
| Left colon | 1 (33.3%) | 1 (33.3%) | 3 (50%) | 6 (33.3%) |
| Rectum | 0 (0%) | 0 (0%) | 2 (33.3%) | 4 (22.2%) |
| **Primary tumor resected, n (%)** | | | | |
| Yes | 3 (100%) | 1 (33.3%) | 6 (100%) | 15 (83.3%) |
| No | 0 (0%) | 2 (66.7%) | 0 (0%) | 3 (16.7%) |
| **Molecular profile, n (%)** | | | | |
| *RAS/BRAF* wild type | 1 (33.3%) | 1(33.3%) | 1 (16.7%) | 8 (44.4%) |
| *KRAS* mutant | 0 (0%) | 0 (0%) | 5 (83.3%) | 10 (55.6%) |
| *NRAS* mutant | 0 (0%) | 0 (0%) | 0 (0%) | 0 (0%) |
| *BRAF* mutant | 2 (66.7%) | 2 (66.7%) | 0 (0%) | 0 (0%) |
| **Number of metastatic sites, n (%)** | | | | |
| 1 | 1 (33.3%) | 0 (0%) | 0 (0%) | 0 (0%) |
| 2 | 1 (33.3%) | 2 (66.7%) | 3 (50%) | 8 (44.4%) |
| >2 | 1 (33.%) | 1 (33.3%) | 3 (50%) | 10 (55.6%) |
| **Peritoneal disease, n (%)** | | | | |
| Yes | 2 (66.7%) | 2 (66.7%) | 2 (33.3%) | 7 (38.9%) |
| No | 1 (33.3%) | 1 (33.3%) | 4 (66.7%) | 11 (61.1%) |
| **Time to liver metastases appearance, n (%)** | | | | |
| At disease debut | 1 (33.3%) | 1 (33.3%) | 3 (50%) | NA |
| At progression to first line therapy | NA | NA | 1 (16.7%) | NA |
| At progression to second line therapy | NA | NA | 0 (0%) | NA |
| At progression to third line therapy | NA | NA | 2 (33.3%) | NA |
| **Prior lines of therapy, n (%)** | | | | |
| 0 | 1 (33.3%) | 2 (66.7%) | 0 (0%) | 0 (0%) |
| 1 | 2 (66.7%) | 1 (33.3%) | 0 (0%) | 1 (5.5%) |
| 2 | 0 (0%) | 0 | 3 (50%) | 5 (27.8%) |
| 3 | 0 (0%) | 0 | 0 (0%) | 7 (38.9%) |
| >3 | 0 (0%) | 0 | 3 (50%) | 5 (27.8%) |
| **Previous systemic therapies for mCRC, n (%)** | | | | |
| Previous fluoropyrimidine, irinotecan, and oxaliplatin | 0 (0%) | 0 (0%) | 6 (50%) | 18 (100%) |
| Any previous anti-VEGF antibody | 0 (0%) | 0 (0%) | 6 (50%) | 16 (88.9%) |
| Any previous anti-EGFR antibody | 0 (0%) | 0 (0%) | 1 (16.7%) | 8 (44.4%) |

**Table 1. Baseline patient and tumor characteristics**

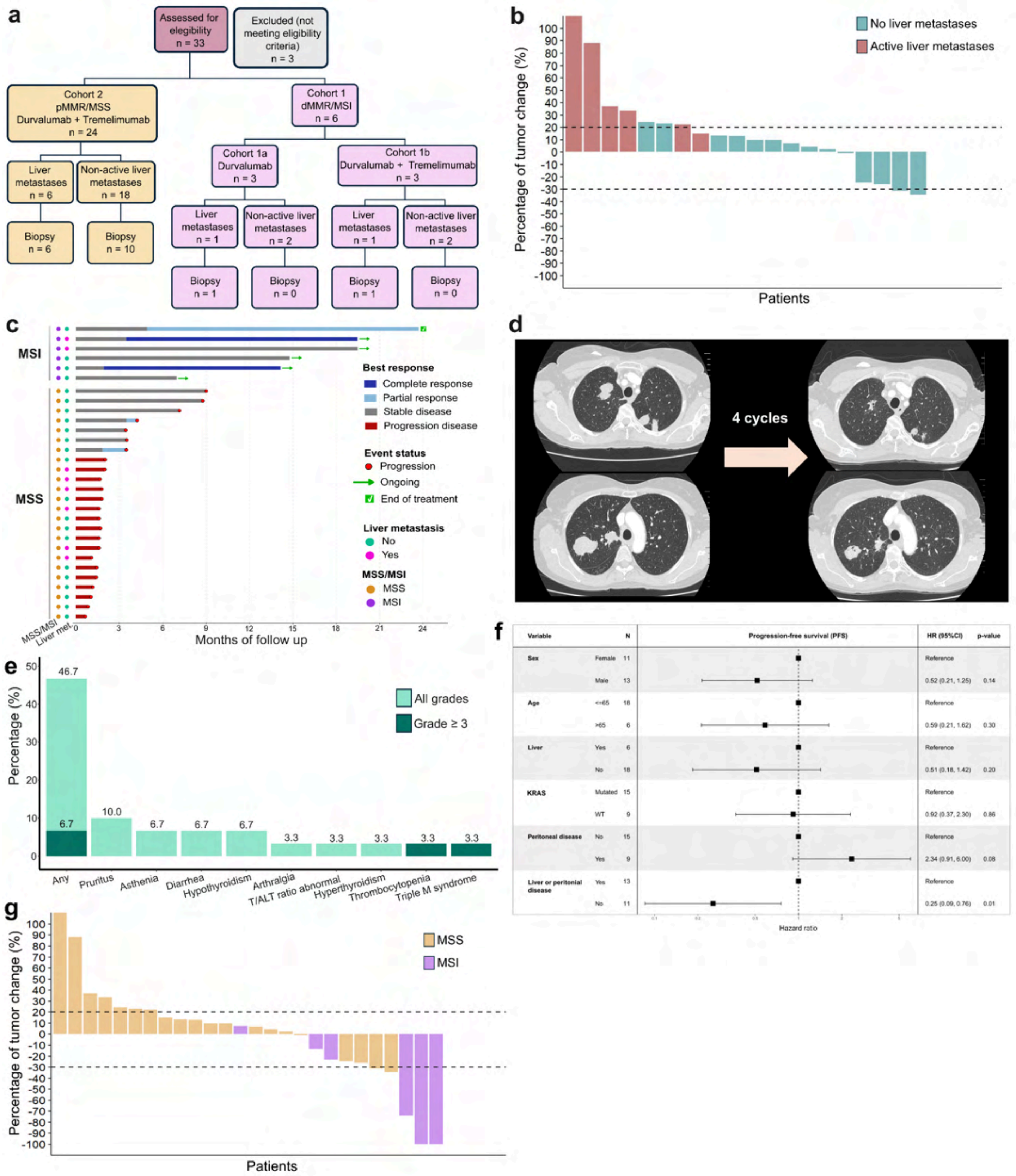


**Figure S1. Clinical trial design and clinical outcomes**

Figure caption on the next page.

**Figure S1. Clinical trial design and clinical outcomes**

**a**, Patient disposition. Flow diagram showing patient recruitment, randomization, and obtention of biopsy specimens. Three patients with mCRC in cohort 2 (pMMR/MSS mCRC) were found to be ineligible (2 due to clinical deterioration unfit for treatment and 1 due to analytical alterations). **b,** Waterfall plot for tumor changes in cohort 2 (pMMR/MSS mCRC) in patients with liver metastases (red) and those without liver metastases (pine green). Three patients presented clinical progression before first radiological evaluation and 1 patient died due to respiratory infection prior to radiological evaluation and was considered not evaluable. **c,** Swimmer plot showing clinical outcomes of all patients in the clinical trial, according to the assigned cohort and presence or not of liver metastases. **d,** Patient with pMMR/MSS refractory mCRC, KRAS G61H, with bilateral lung lesions at baseline, who receivedDurvalumab + Tremelimumab as fifth line. Computed tomography after 4 cycles of treatment showed partial response. **e,** Treatment-related adverse events. Light green shows all grades of toxicity, while dark green represents grade 3 or higher. **f**, Univariate analysis for progression-free survival in patients with pMMR/MSS CRC. **g**, Waterfall plot for tumor change in the overall cohort in patients with pMMR/MSS (yellow) and dMMR/MSI (purple) those without liver metastases (pine green). Three patients presented clinical progression before first radiological evaluation and 1 patient died due to respiratory infection prior to radiological evaluation and was considered not evaluable, all of them had pMMR/MSS mCRC.

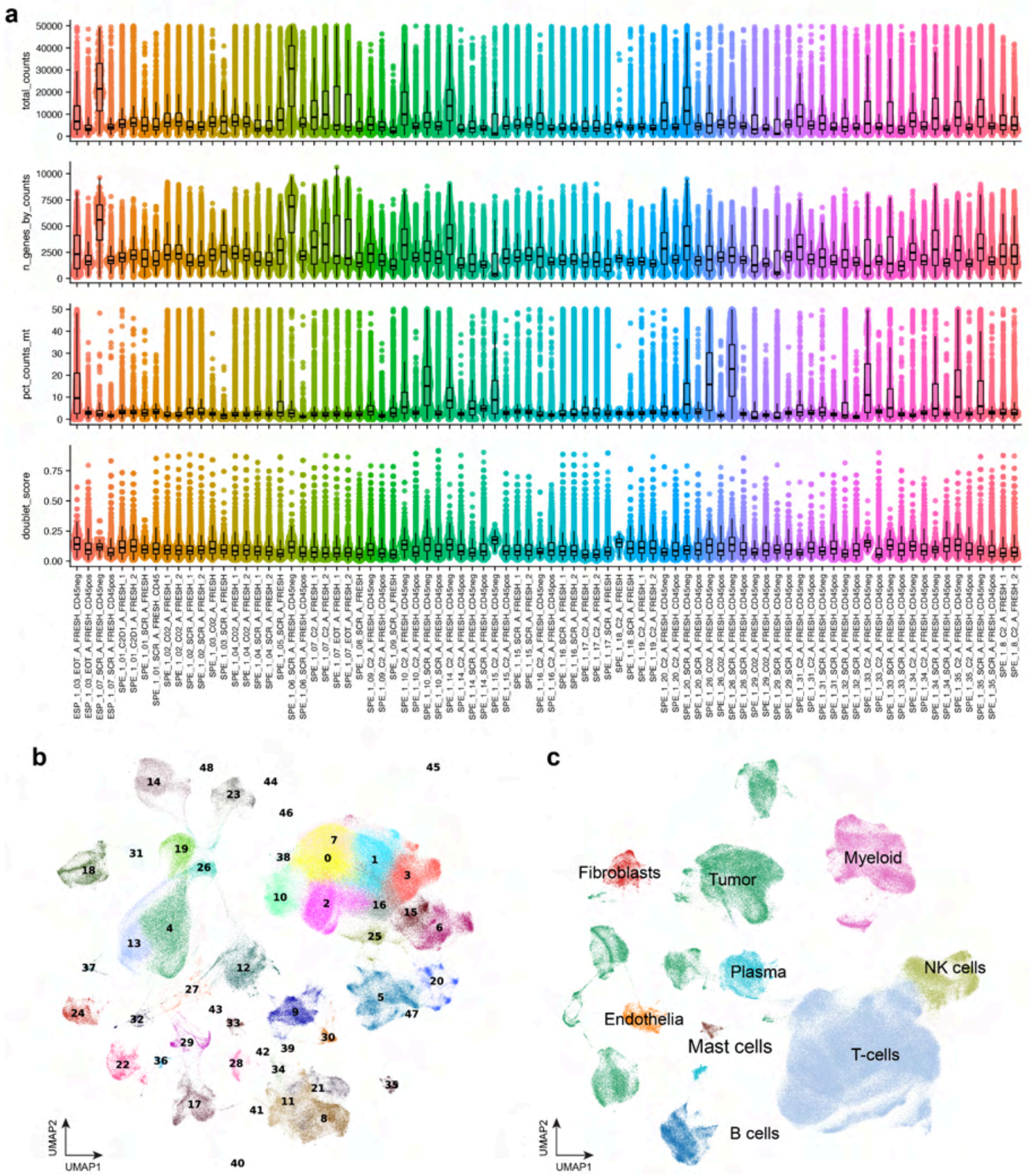


**Figure S2. single-cell RNA-seq initial clustering and quality control**

**a**, Distributions of quality control metrics by sequencing library. Doublet scores were calculated using Scrublet[1]. For all box-and-whisker plots, box boundaries and lines correspond to the interquartile range (IQR) and median, respectively. Whiskers extend to the lowest or highest data points that are no further from 1.5 times the IQR from the box boundaries. **b**, Uniform manifold approximation and projection (UMAP) of 486,410 cells from all sequencing libraries from the SERPENTINE cohort, before additional quality control (QC) filtering. The UMAP is based on the top 30 principal components, without any additional batch correction. Cells are colored by leiden clustering assignment. **C**, UMAP plot of 456,653 after initial QC filtering, colored by major cell lineages.

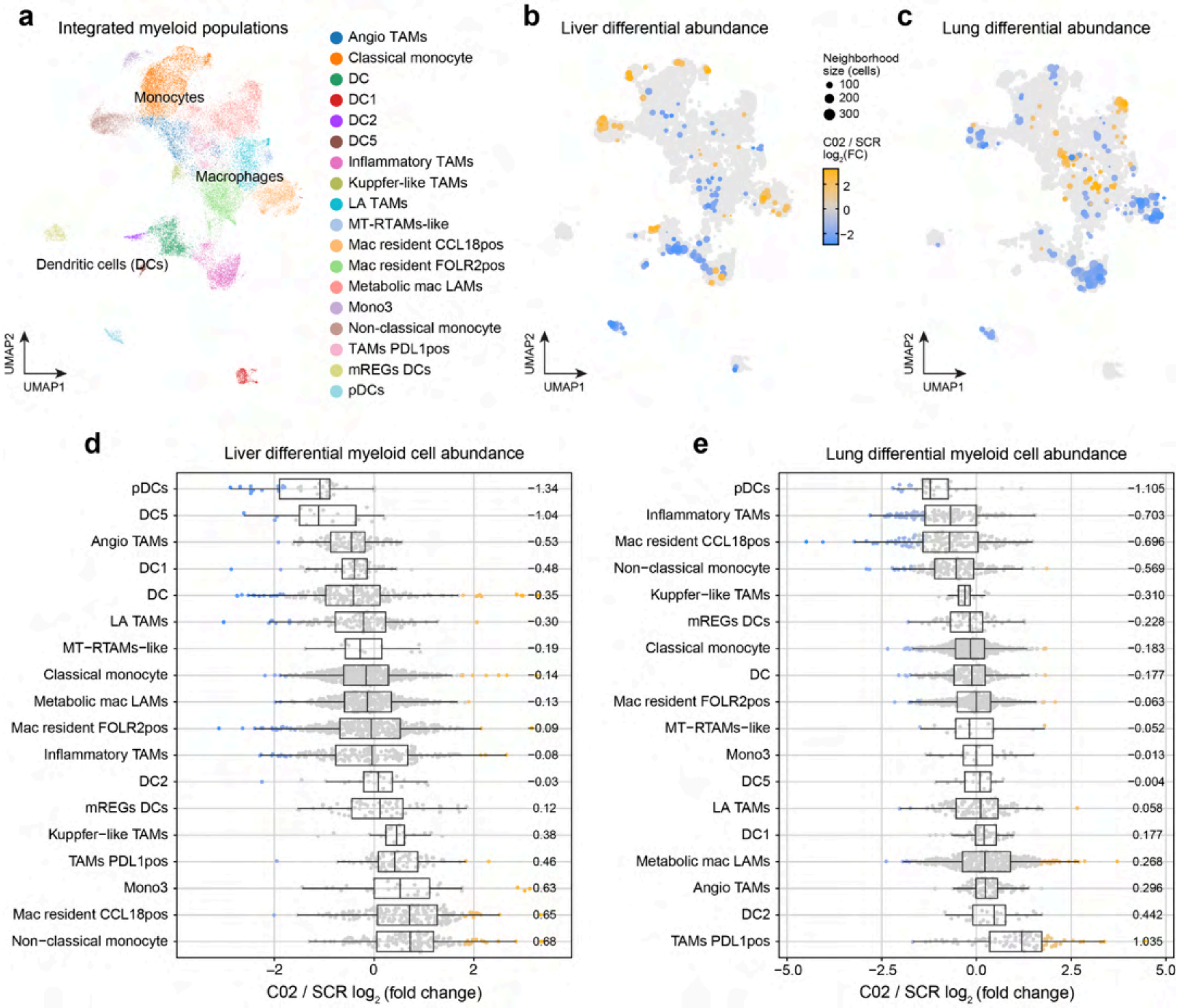


**Figure S3. Myeloid cell clustering and differential cell abundance analysis**

Uniform manifold approximation and projection (UMAP) of 37,229 Myeloid lineage cells integrated between the SERPENTINE cohort and primary CRC TMEs from an external dataset[2]. Datasets were integrated with Harmony[3] and CellANOVA[4]. Each point represents one cell, colored by their cell-type annotations derived from Leiden clustering[5] analysis. **b-c**, Differential myeloid cell abundance analysis comparing post-treatment (C02) vs. pre-treatment (SCR) within the liver (**b**) and lung (**c**) TMEs. Points represent cellular neighborhoods from Milo[6], colored by the effect size of the comparison as log2(fold-change). Orange color indicates enrichment of cell populations post-treatment, and blue color indicates enrichment pre-treatment. **d-e**, Milo differential abundance results shown as box-and-whisker plots for each cell type. Median effect sizes are annotated on the right side. For box-and-whisker plots, box boundaries and lines correspond to the interquartile range (IQR) and median, respectively. Whiskers extend to the lowest or highest data points that are no further from 1.5 times the IQR from the box boundaries.

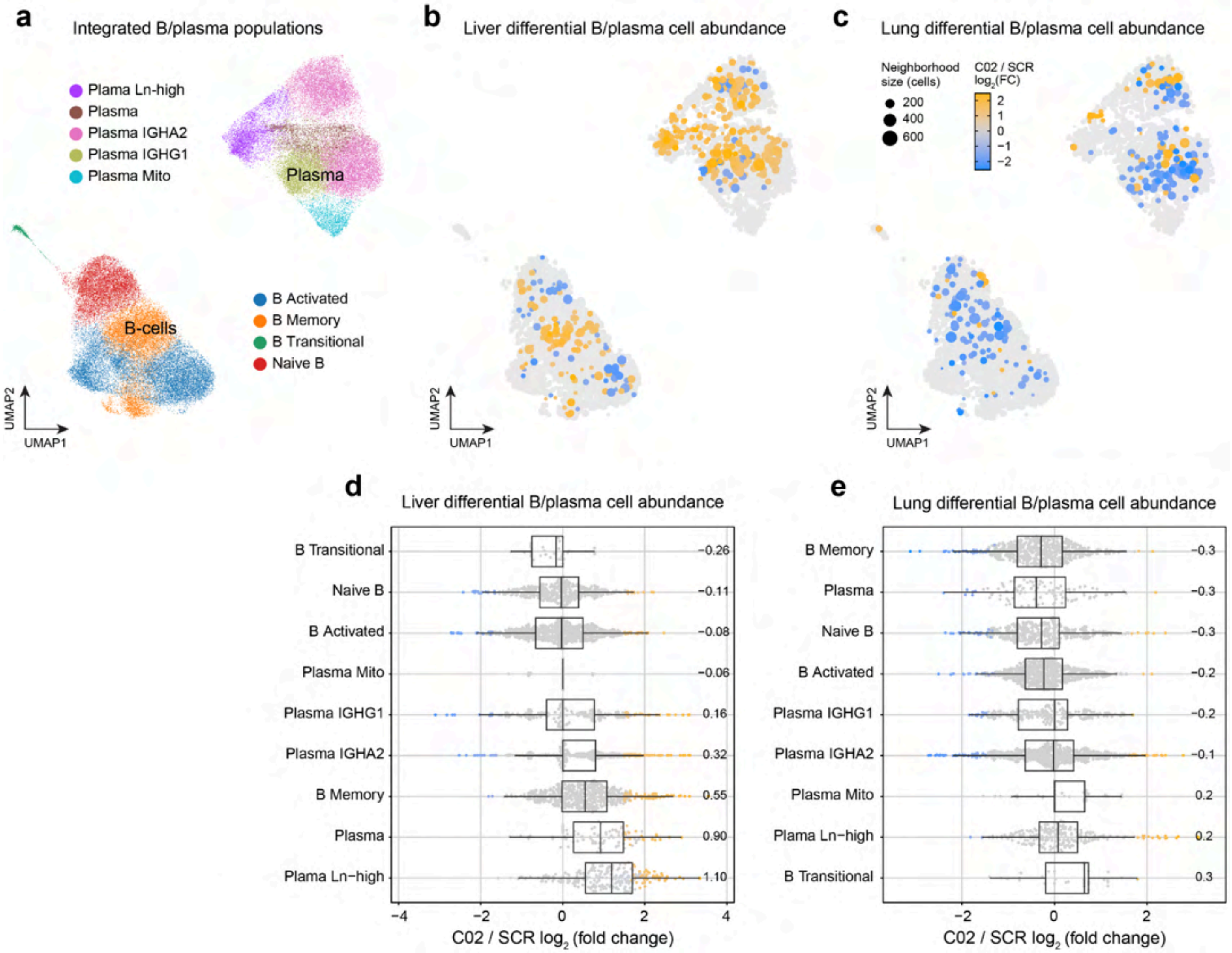


**Figure S4. B and plasma cell clustering and differential cell abundance analysis**

Uniform manifold approximation and projection (UMAP) of 67,043 B/plasma cells integrated between the SERPENTINE cohort and primary CRC TMEs from an external dataset[2]. Datasets were integrated with Harmony[3] and CellANOVA[4]. Each point represents one cell, colored by their cell-type annotations derived from Leiden clustering[5] analysis. **b-c**, Differential B/plasma cell abundance analysis comparing post-treatment (C02) vs. pre-treatment (SCR) within the liver (**b**) and lung (**c**) TMEs. Points represent cellular neighborhoods from Milo[6], colored by the effect size of the comparison as log2(fold-change). Orange color indicates enrichment of cell populations post-treatment, and blue color indicates enrichment pre-treatment. **d-e**, Milo differential abundance results shown as box-and-whisker plots for each cell type. Median effect sizes are annotated on the right side. For box-and-whisker plots, box boundaries and lines correspond to the interquartile range (IQR) and median, respectively. Whiskers extend to the lowest or highest data points that are no further from 1.5 times the IQR from the box boundaries.

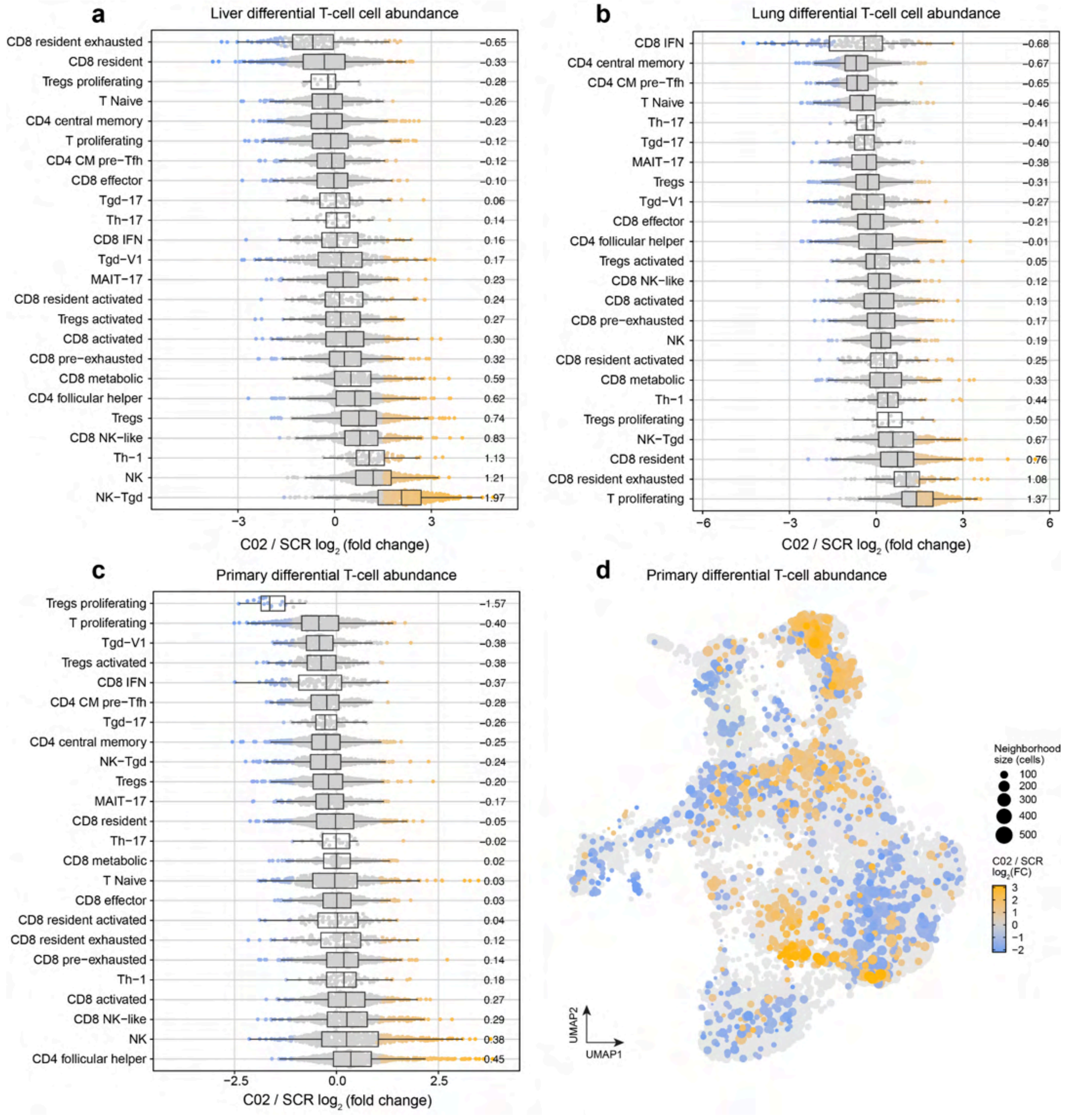


**Figure S5. T-cell differential cell abundance analysis**

**a-c**, Milo differential abundance cell abundance analysis comparing post-treatment (C02) vs. pre-treatment (SCR) shown as box-and-whisker plots for each tissue. Median effect sizes are annotated on the right side. For box-and-whisker plots, box boundaries and lines correspond to the interquartile range (IQR) and median, respectively. Whiskers extend to the lowest or highest data points that are no further from 1.5 times the IQR from the box boundaries. **d**, The same differential abundance results as in panel (**c**) shown as a UMAP plot.

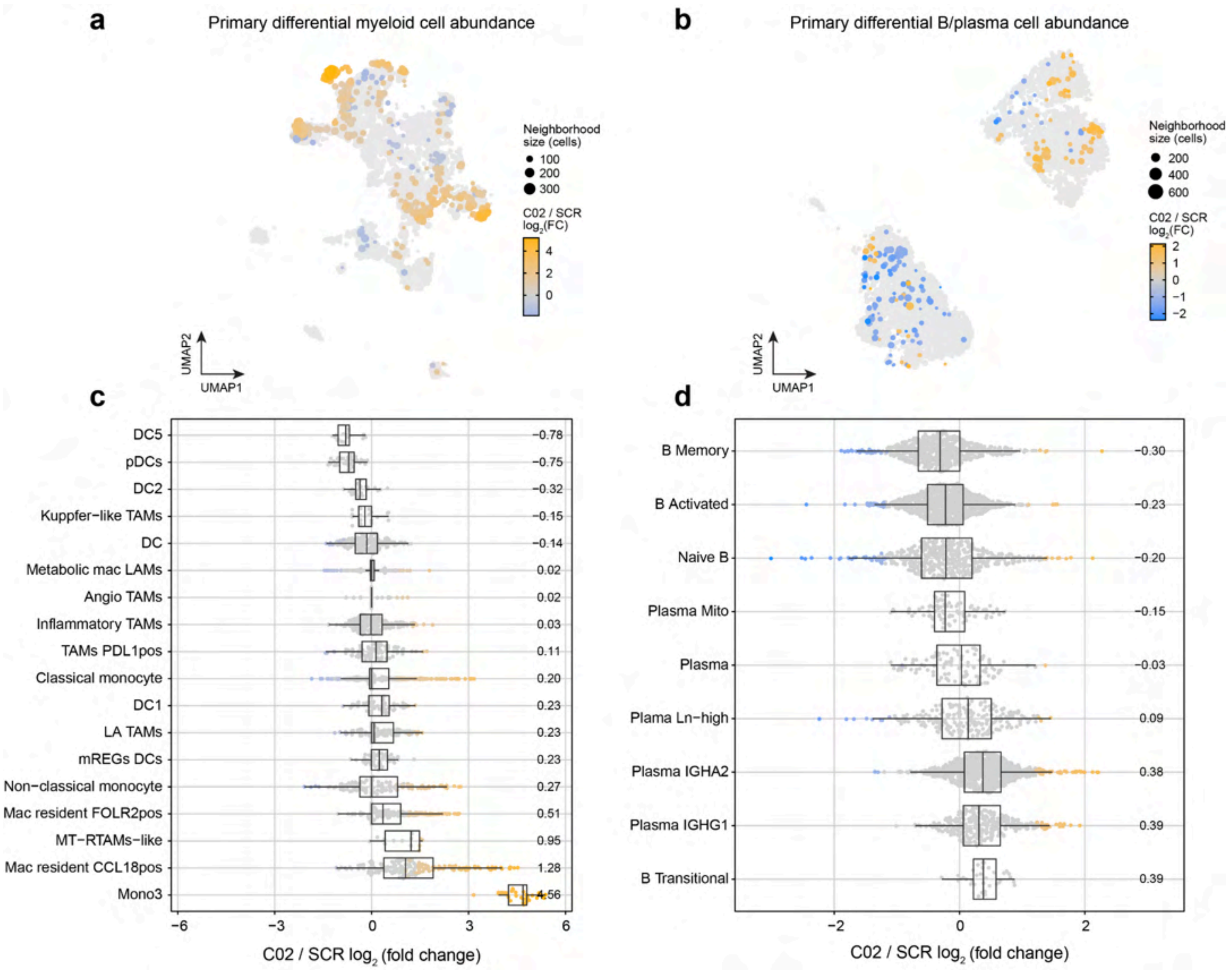


**Figure S6. Primary tumor differential cell abundance analysis**

**a-b**, Differential cell abundance analysis comparing post-treatment (C02) vs. pre-treatment (SCR) within the primary tumor dataset for the myeloid lineage (**a**) and the B/plasma lineage (**c**). Points represent cellular neighborhoods from Milo[6], colored by the effect size of the comparison as log2(fold-change). Orange color indicates enrichment of cell populations post-treatment, and blue color indicates enrichment pre-treatment. **c-d**, Milo differential abundance results shown as box-and-whisker plots for each cell type. Median effect sizes are annotated on the right side. For box-and-whisker plots, box boundaries and lines correspond to the interquartile range (IQR) and median, respectively. Whiskers extend to the lowest or highest data points that are no further from 1.5 times the IQR from the box boundaries.

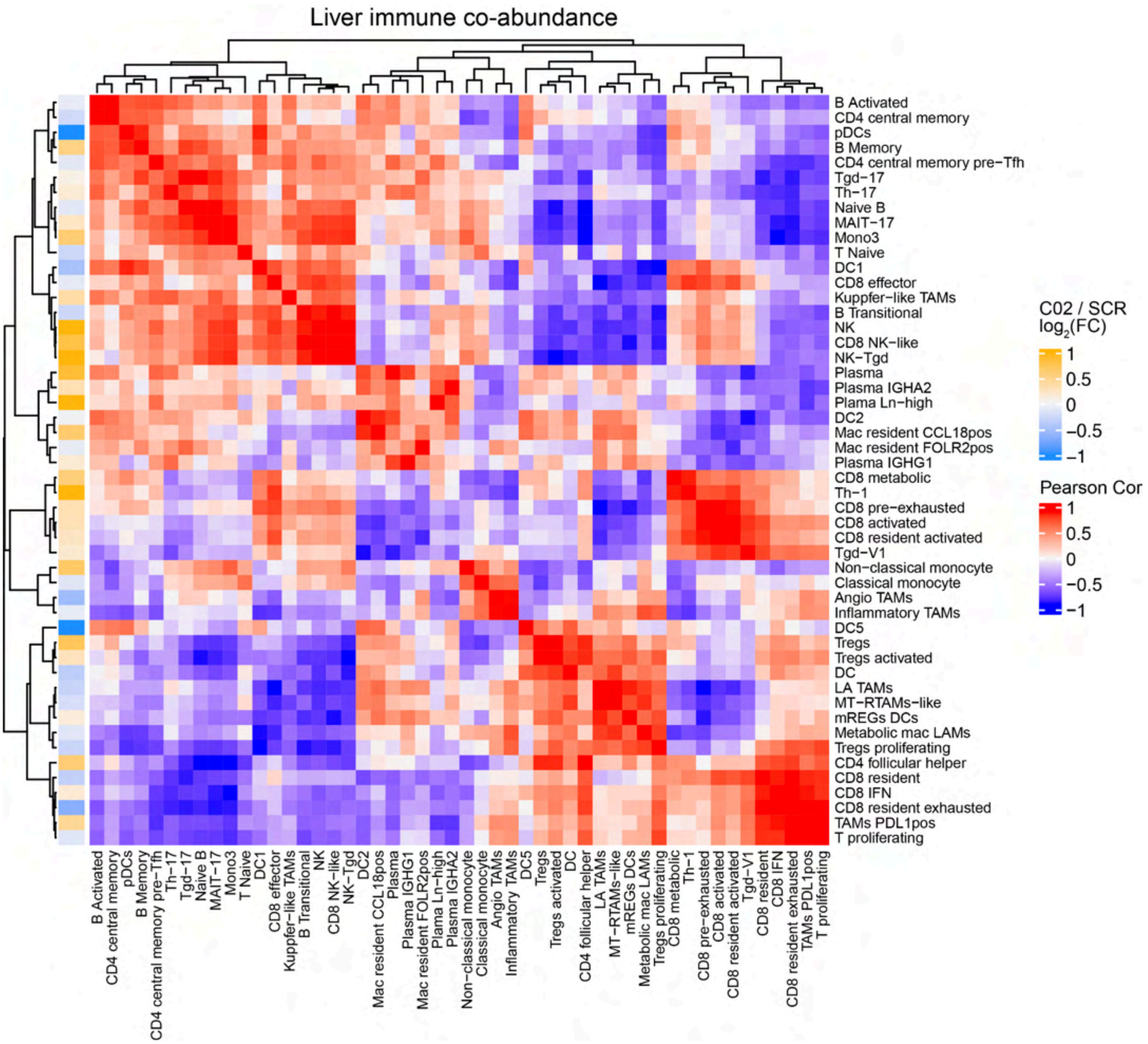


**Figure S7. Liver metastasis immune co-abundance heatmap**

Hierarchical clustering of immune cell populations in liver metastasis based on the similarity of their treatment-induced abundance dynamics. The main heatmap displays the pairwise Pearson correlation coefficients calculated from the treatment-induced shift in centered log-ratio transformed proportions (ΔCLR; post-treatment vs. pre-treatment). The left annotation bar indicates the average effect size in differential abundance between post-treatment (C02) and screening (SCR) timepoints from the previously described Milo analysis.

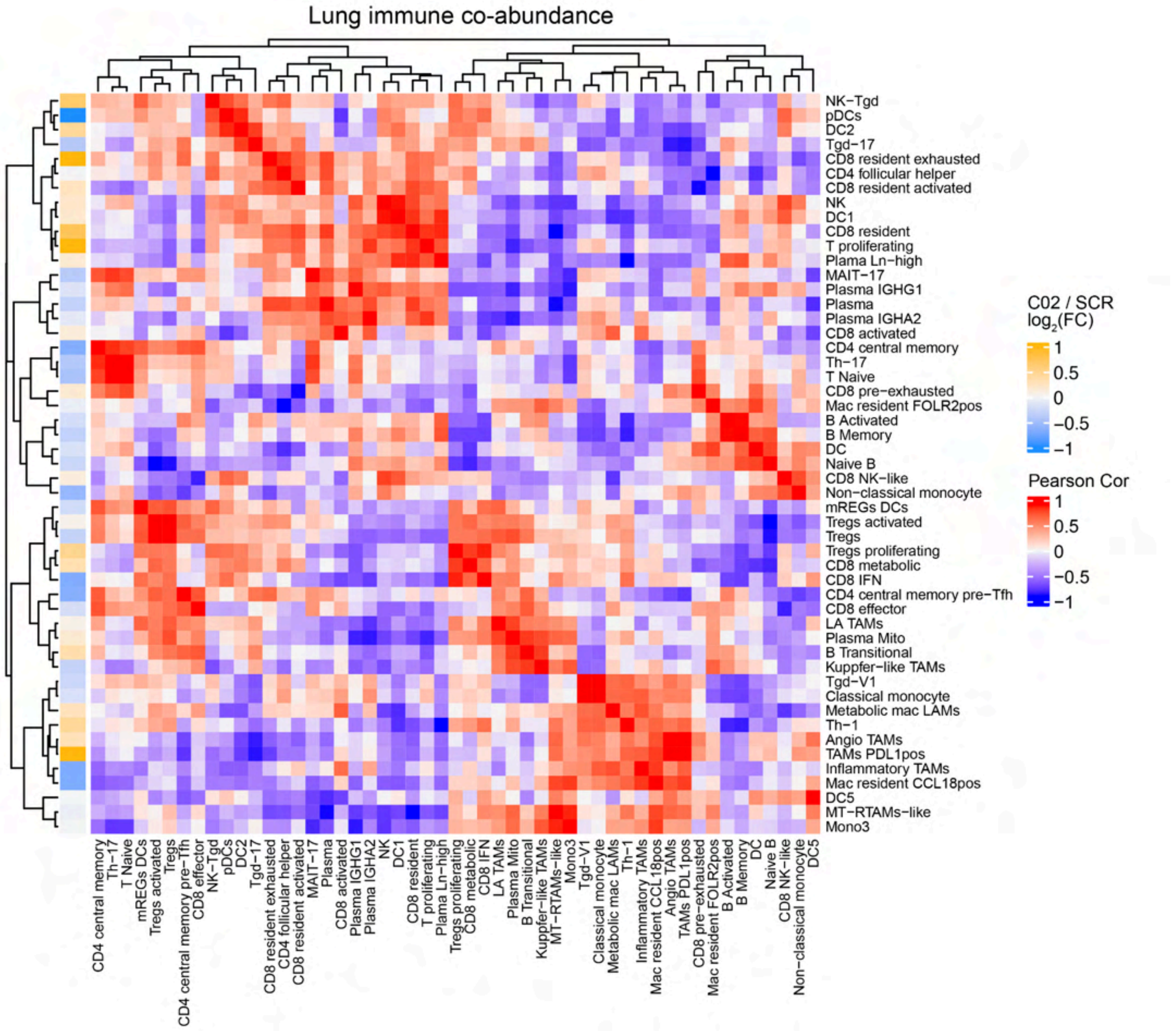


**Figure S8. Lung metastasis immune co-abundance heatmap**

Hierarchical clustering of immune cell populations in lung metastasis based on the similarity of their treatment-induced abundance dynamics. The main heatmap displays the pairwise Pearson correlation coefficients calculated from the treatment-induced shift in centered log-ratio transformed proportions (ΔCLR; post-treatment vs. pre-treatment). The left annotation bar indicates the average effect size in differential abundance between post-treatment (C02) and screening (SCR) timepoints from the previously described Milo analysis.

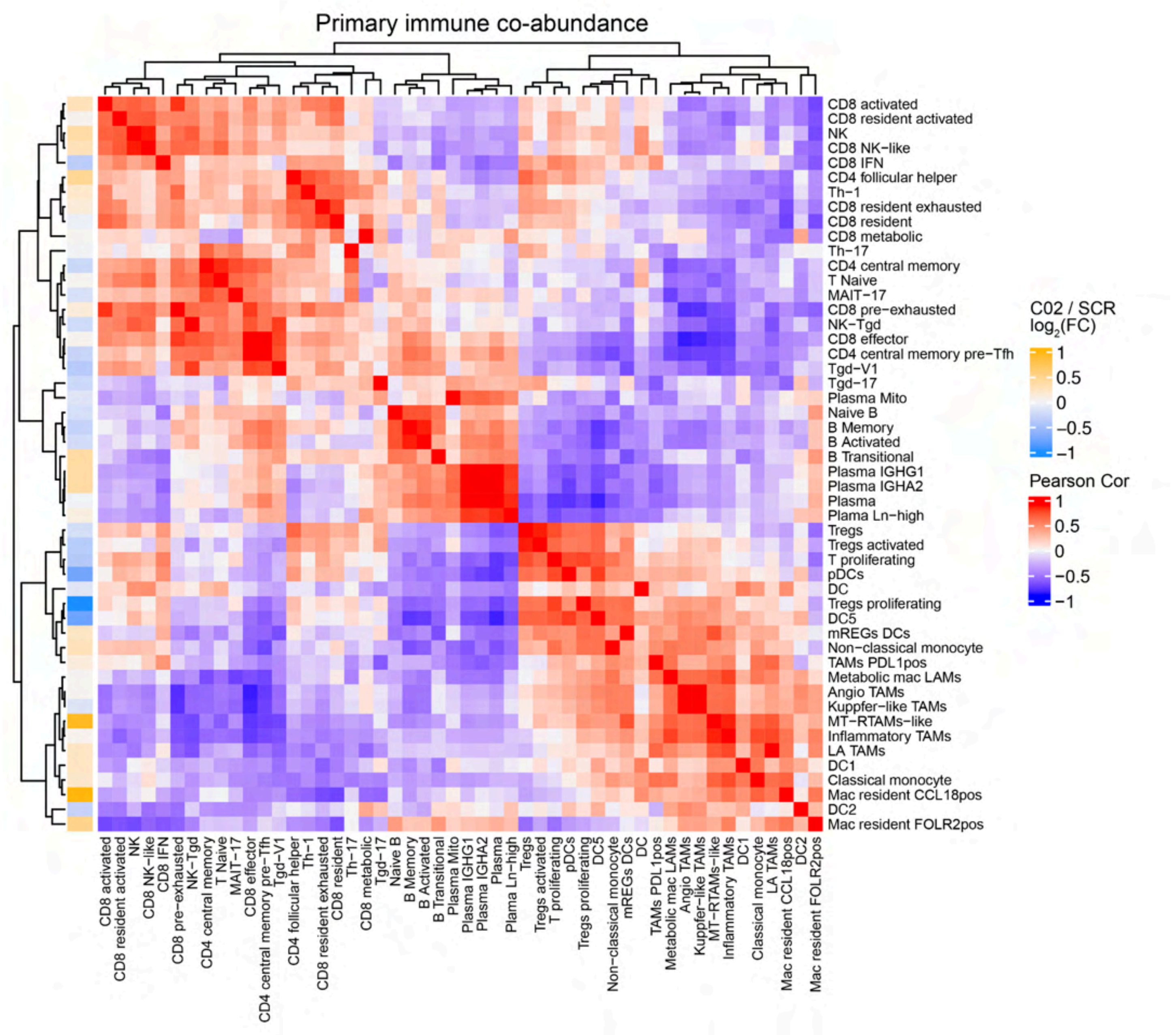


**Supplementary Fig. 9 | Primary tumor immune co-abundance heatmap.** Hierarchical clustering of immune cell populations in the primary CRC TME based on the similarity of their treatment-induced abundance dynamics. The main heatmap displays the pairwise Pearson correlation coefficients calculated from the treatment-induced shift in centered log-ratio transformed proportions (ΔCLR; post-treatment vs. pre-treatment). The left annotation bar indicates the average effect size in differential abundance between post-treatment (C02) and screening (SCR) timepoints from the previously described Milo analysis.

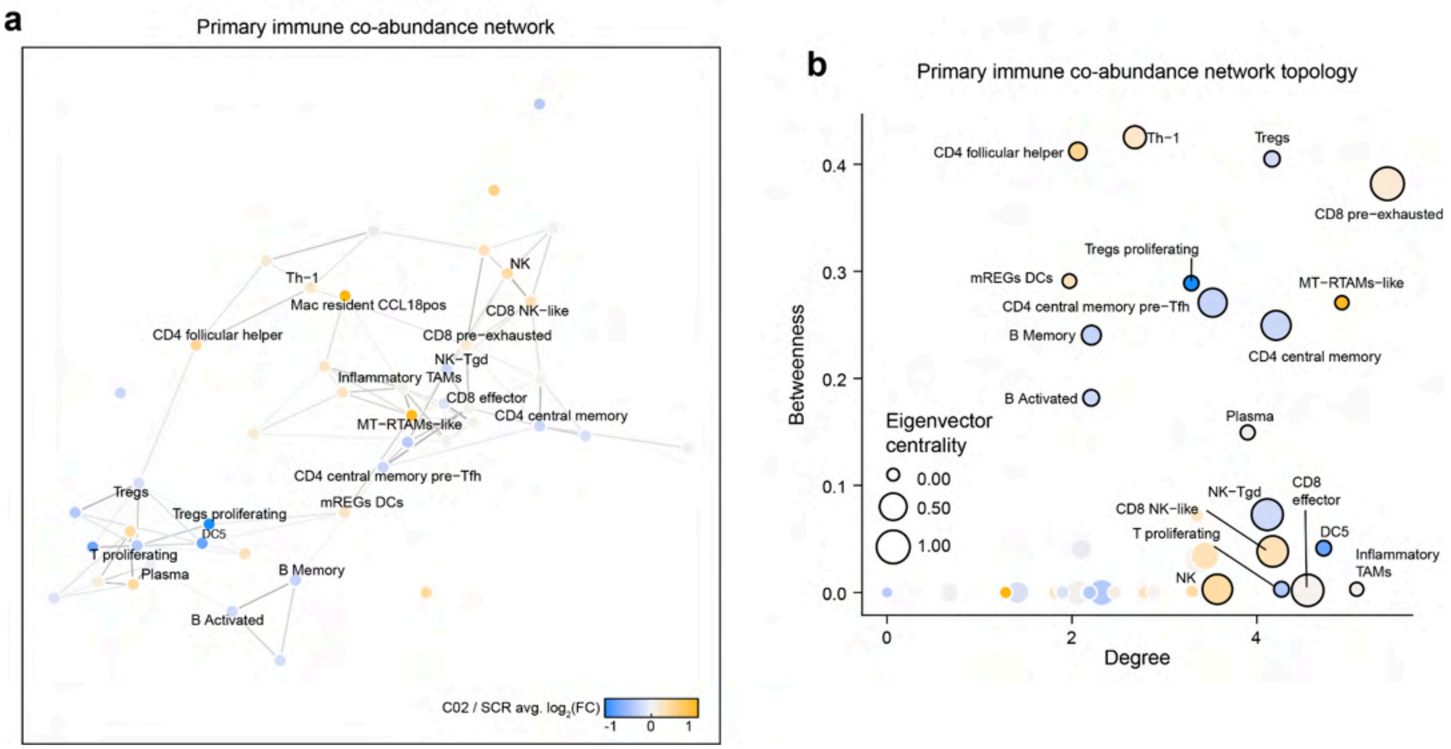


**Figure S10. Primary tumor immune co-abundance network**

**a**, Systems-level immune co-abundance dynamics in the primary CRC TME. Nodes represent immune populations, and edges represent similarity in treatment-induced abundance changes (Pearson's $R > 0.6$ and $p_{perm} < 0.05$). Nodes are colored by the average effect size of the Milo differential abundance test. Selected immune populations are labeled. **b** Topological analysis of the primary CRC TME immune co-abundance networks. Node degree (local connectivity) and betweenness centrality (global bridge capacity) are compared for each immune population, and node size represents eigenvector centrality (prestige).

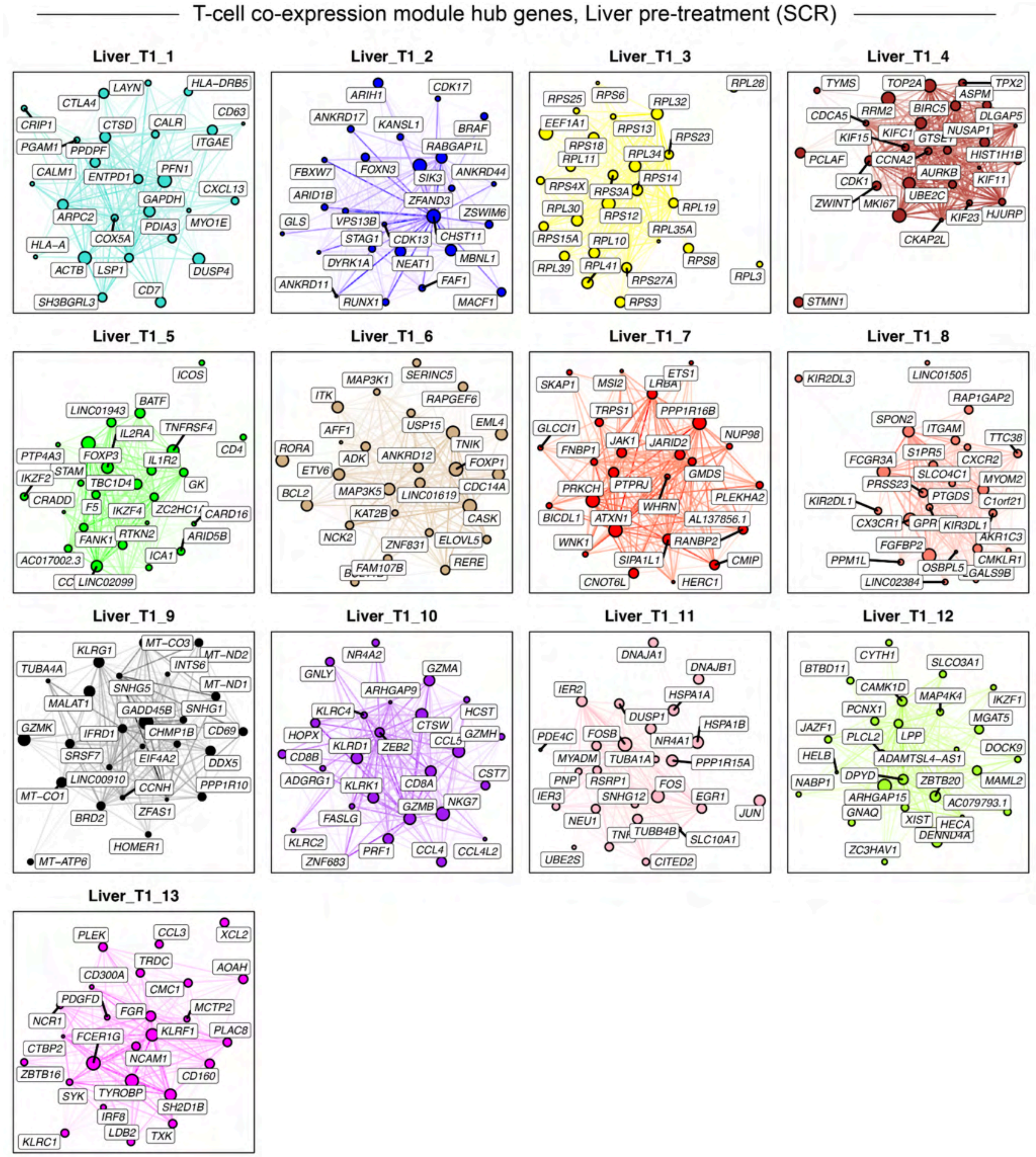


**Figure S11. T-cell gene co-expression module hub genes, liver metastasis pre-treatment**

Hub gene network for each of the 13 Liver pre-treatment (SCR) T-cell lineage gene co-expression modules identified via hdWGCNA[7]. Nodes represent genes, and edges represent co-expression links (topological overlap). The top 25 hub genes ranked by eigengene-based intramodular connectivity (kME) are shown, and the top 250 edges per module are shown.

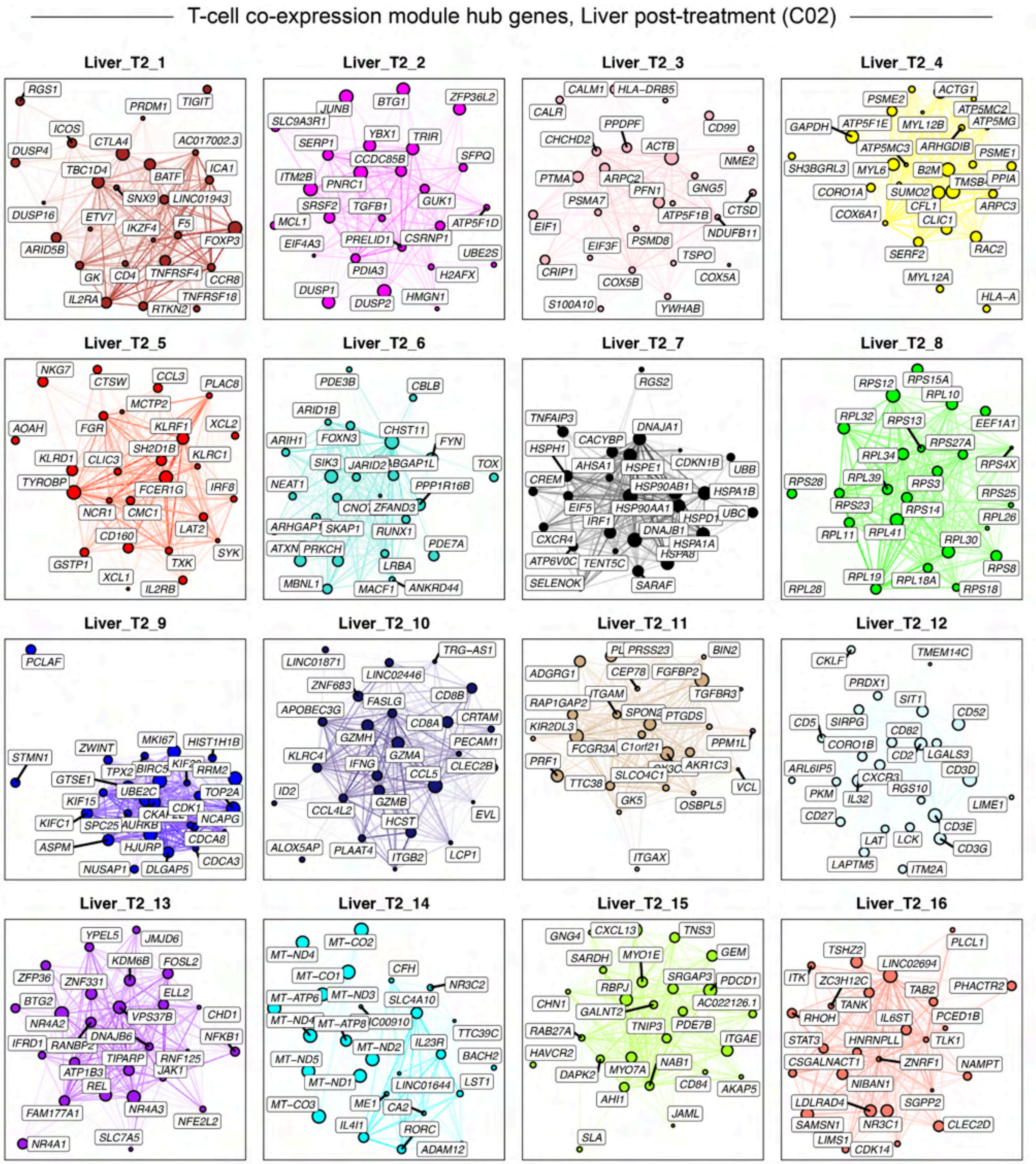


**Figure S12. T-cell gene co-expression module hub genes, liver metastasis post-treatment**

Hub gene network for each of the 16 Liver post-treatment (C02) T-cell lineage gene co-expression modules identified via hdWGCNA[7]. Nodes represent genes, and edges represent co-expression links (topological overlap). The top 25 hub genes ranked by eigengene-based intramodular connectivity (kME) are shown, and the top 250 edges per module are shown.

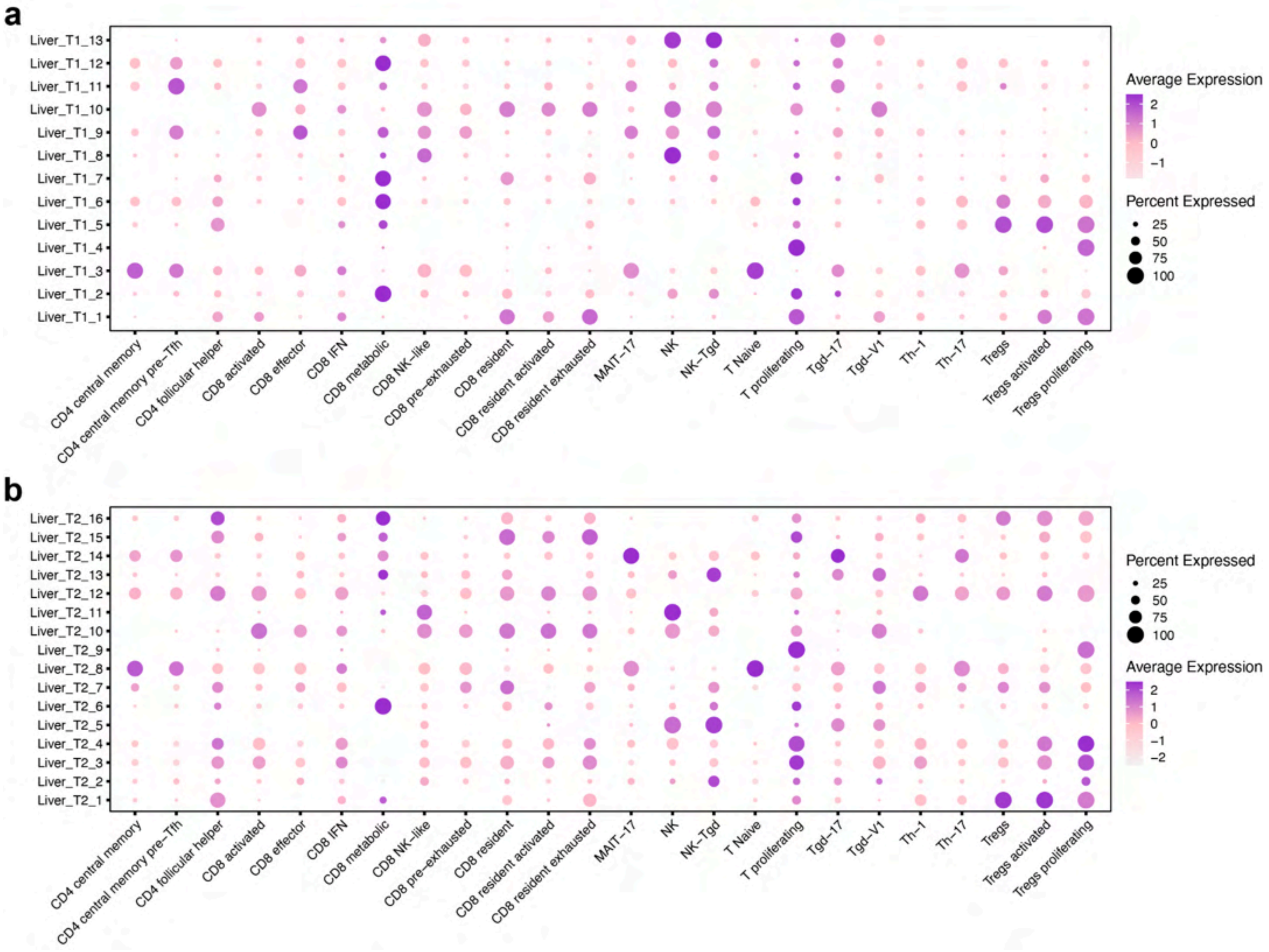


**Figure S13. Liver metastasis T-cell gene co-expression module expression levels**

Dot plots showing the single-cell gene-expression signatures (Module Eigengenes, MEs) for each of the co-expression modules pre-treatment (**a**) and post-treatment (**b**) in each cluster.

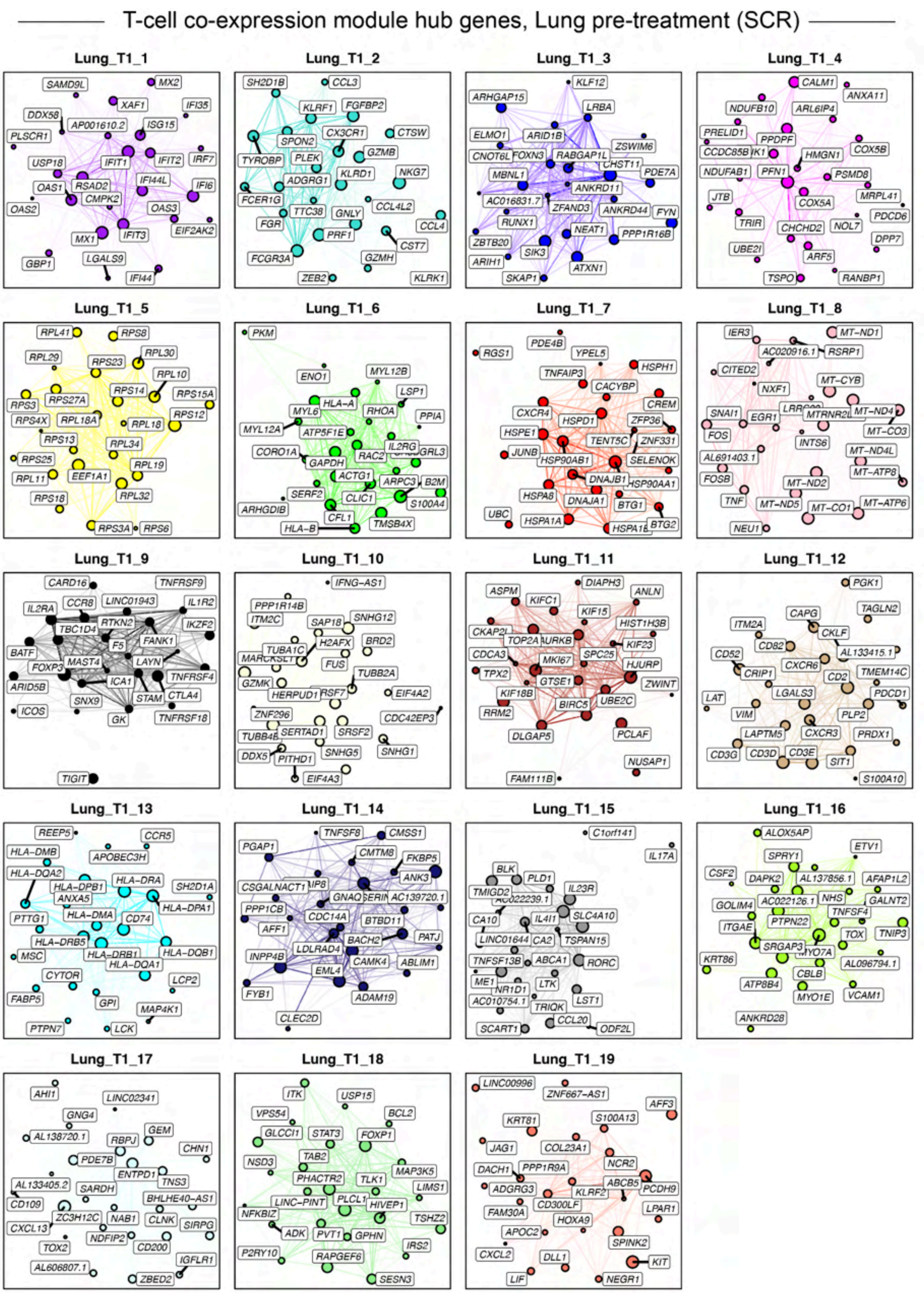


**Figure S14. T-cell gene co-expression module hub genes, lung metastasis pre-treatment**

Figure caption on the next page.

**Figure S14. T-cell gene co-expression module hub genes, lung metastasis pre-treatment**

Hub gene network for each of the 14 lung pre-treatment (SCR) T-cell lineage gene co-expression modules identified via hdWGCNA[7]. Nodes represent genes, and edges represent co-expression links (topological overlap). The top 25 hub genes ranked by eigengene-based intramodular connectivity (kME) are shown, and the top 250 edges per module are shown.

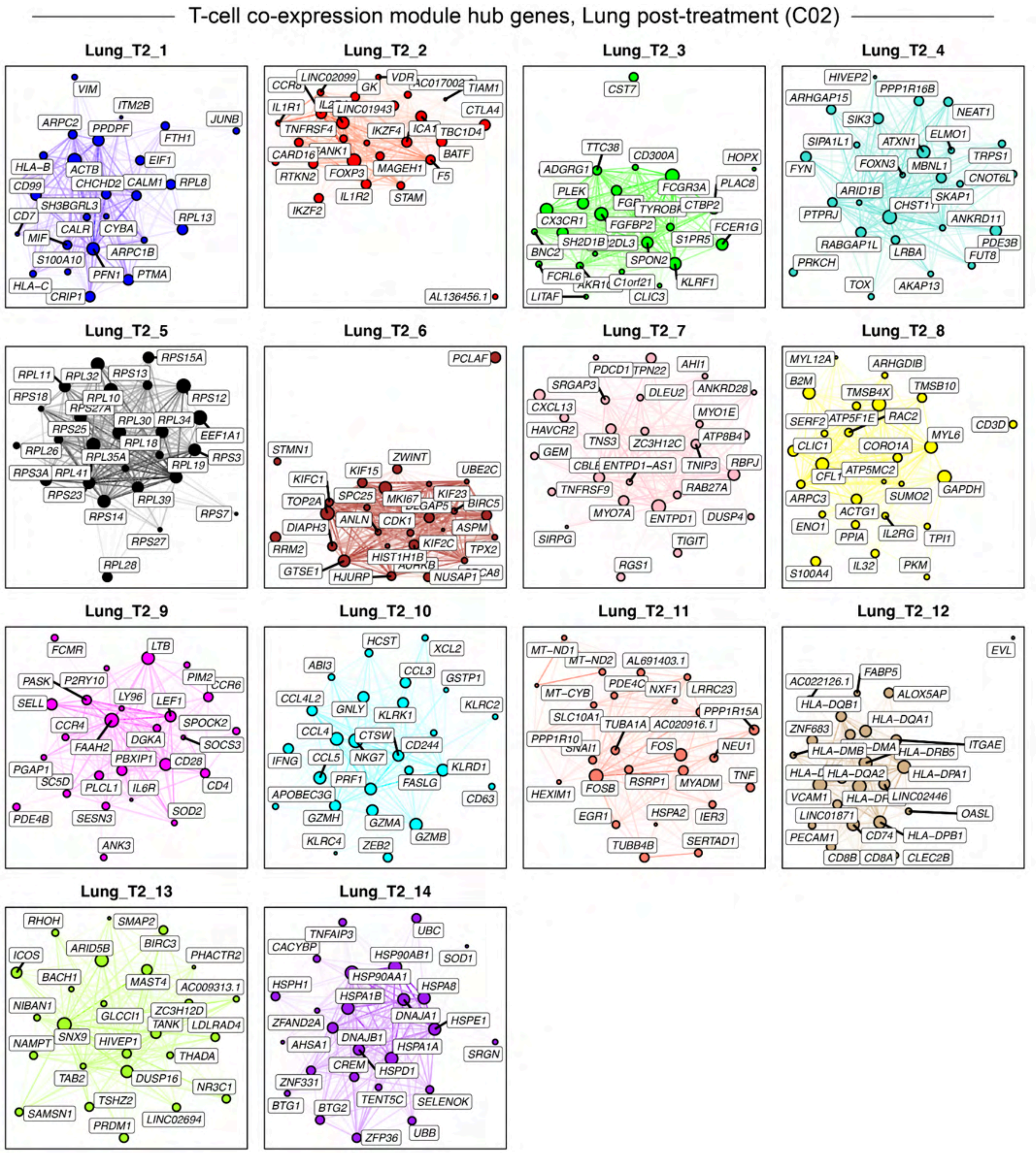


**Figure S15. T-cell gene co-expression module hub genes, lung metastasis post-treatment**

Hub gene network for each of the 14 lung post-treatment (C02) T-cell lineage gene co-expression modules identified via hdWGCNA[7]. Nodes represent genes, and edges represent co-expression links (topological overlap). The top 25 hub genes ranked by eigengene-based intramodular connectivity (kME) are shown, and the top 250 edges per module are shown.

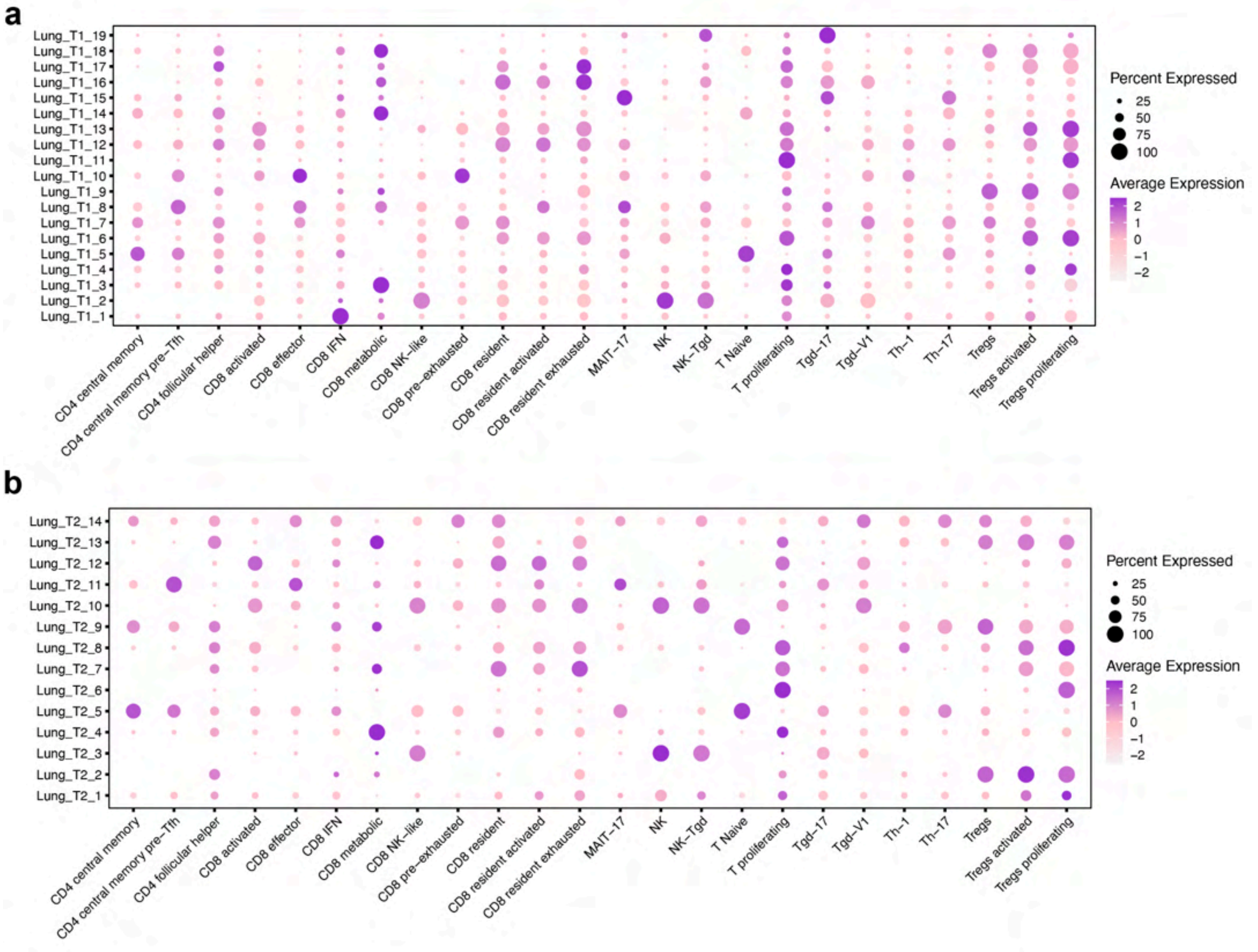


**Figure S16. Lung metastasis T-cell gene co-expression module expression levels**

Dot plots showing the single-cell gene-expression signatures (Module Eigengenes, MEs) for each of the co-expression modules pre-treatment (**a**) and post-treatment (**b**) in each cluster.

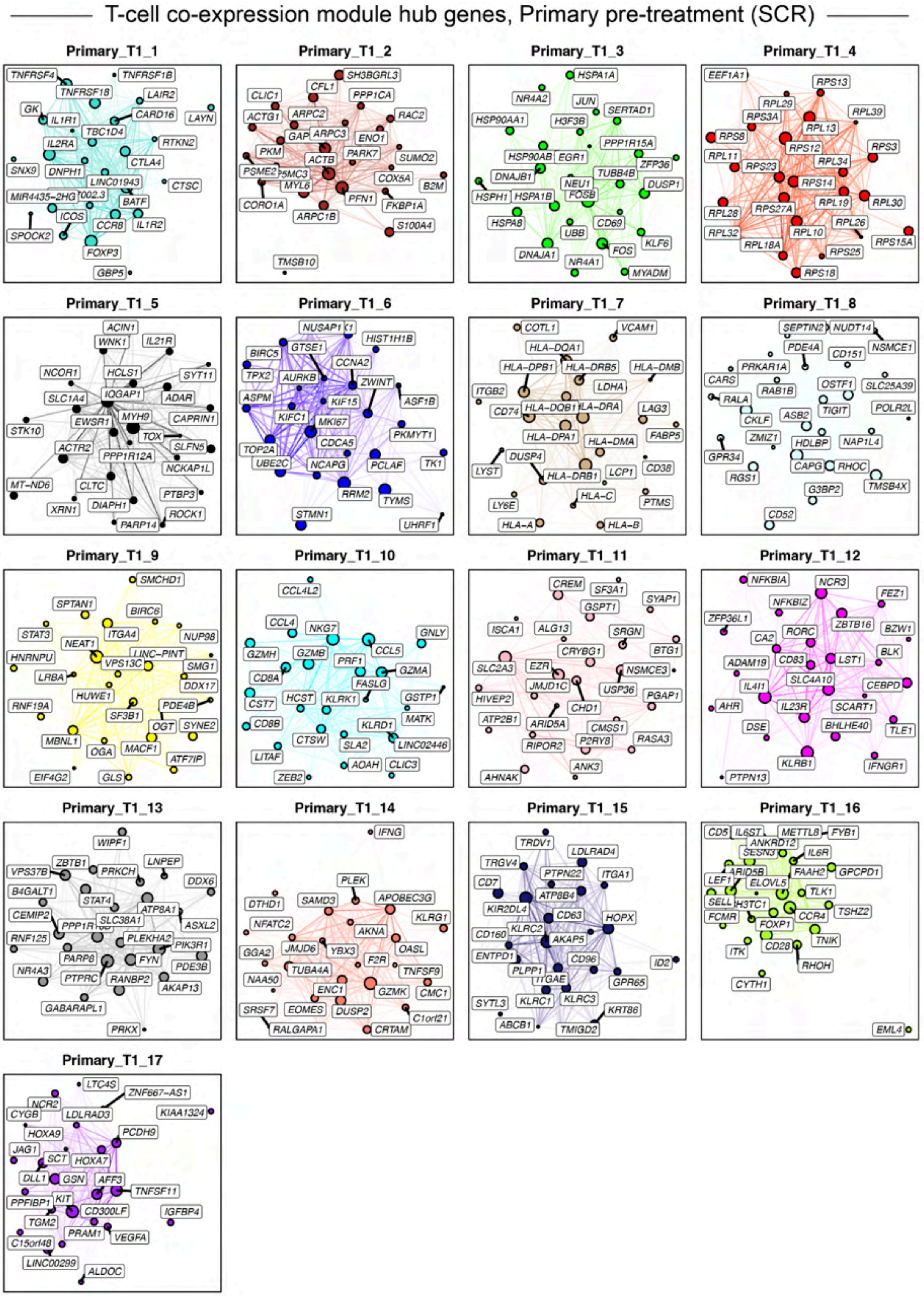


**Figure S16. T-cell gene co-expression module hub genes, primary tumor pre-treatment**

Figure caption on the next page.

**Figure S16. T-cell gene co-expression module hub genes, primary tumor pre-treatment**

Hub gene network for each of the 17 primary tumor pre-treatment T-cell lineage gene co-expression modules identified via hdWGCNA[7]. Nodes represent genes, and edges represent co-expression links (topological overlap). The top 25 hub genes ranked by eigengene-based intramodular connectivity (kME) are shown, and the top 250 edges per module are shown.

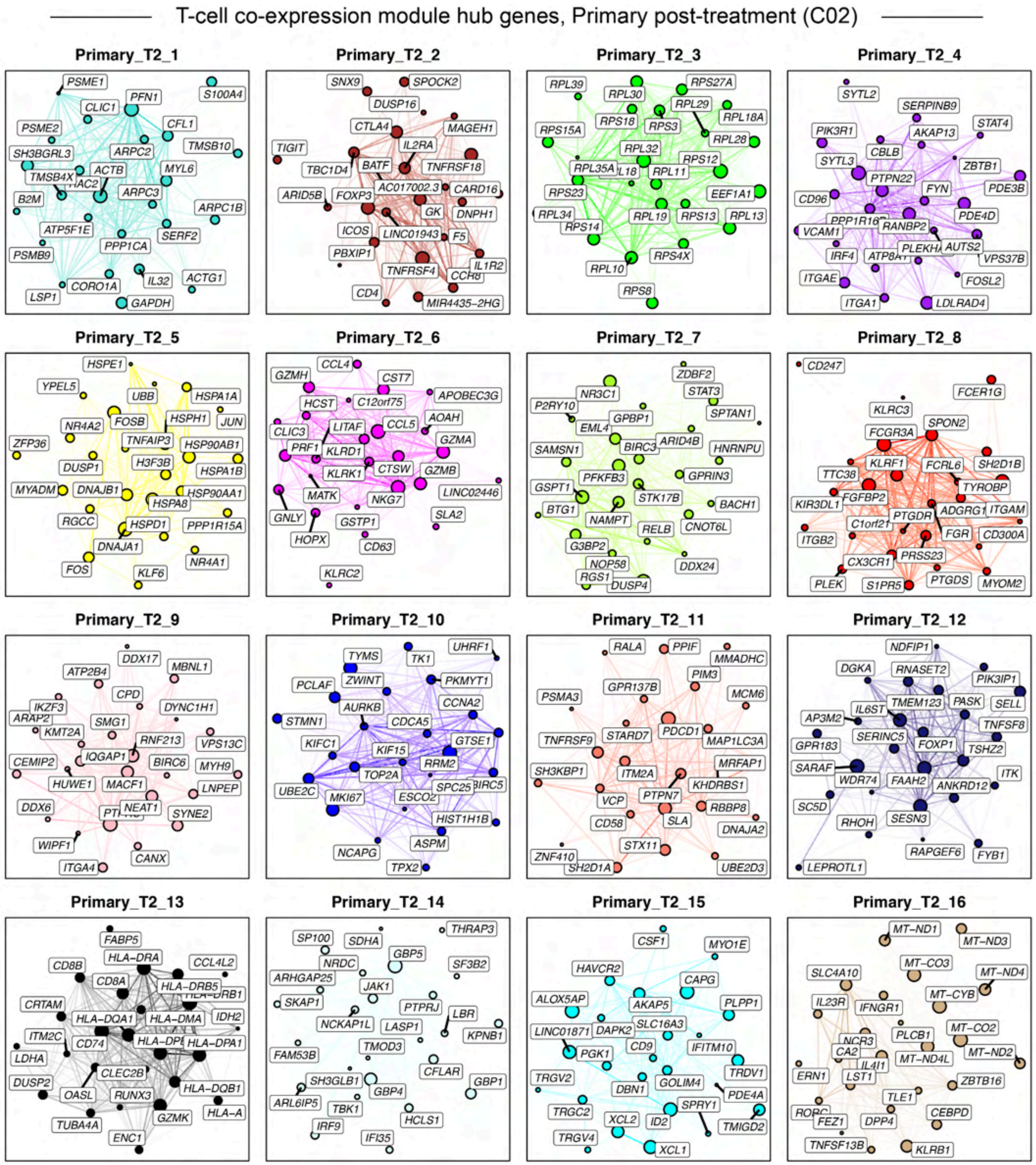


**Figure 17. T-cell gene co-expression module hub genes, primary post-treatment**

Hub gene network for each of the 16 primary tumor post-treatment T-cell lineage gene co-expression modules identified via hdWGCNA[7]. Nodes represent genes, and edges represent co-expression links (topological overlap). The top 25 hub genes ranked by eigengene-based intramodular connectivity (kME) are shown, and the top 250 edges per module are shown.

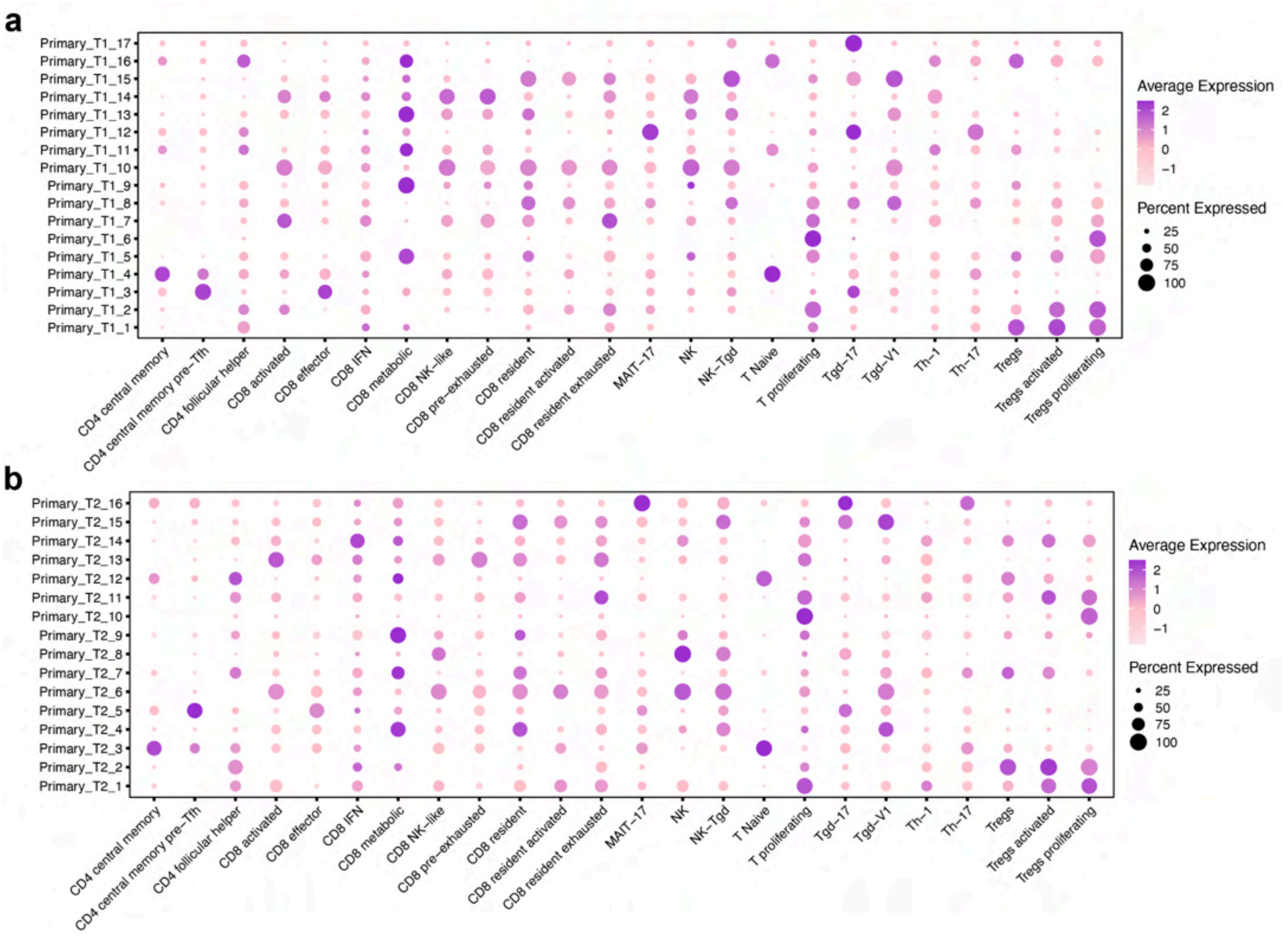


**Figure S18. Primary tumor T-cell gene co-expression module expression levels**

Dot plots showing the single-cell gene-expression signatures (Module Eigengenes, MEs) for each of the co-expression modules pre-treatment (**a**) and post-treatment (**b**) in each cluster.

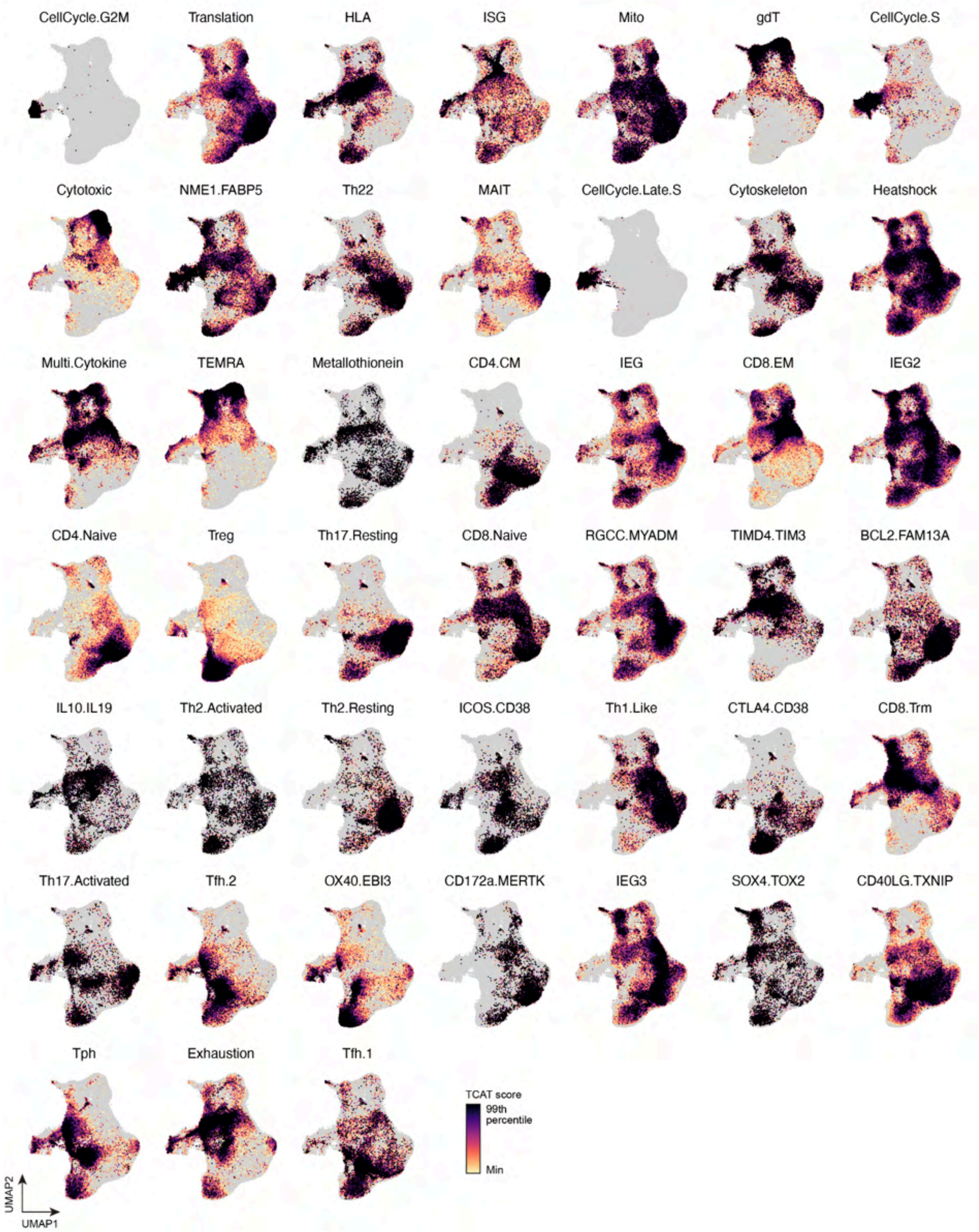


**Figure S19. Activity of T-cell consensus gene expression programs (cGEPs)**

UMAP plots showing single-cell activity scores for each of the T-cell consensus gene expression programs[8]. To avoid biasing the visualization by outliers, we cap the dynamic color range based on the 99th percentile of expression per feature.

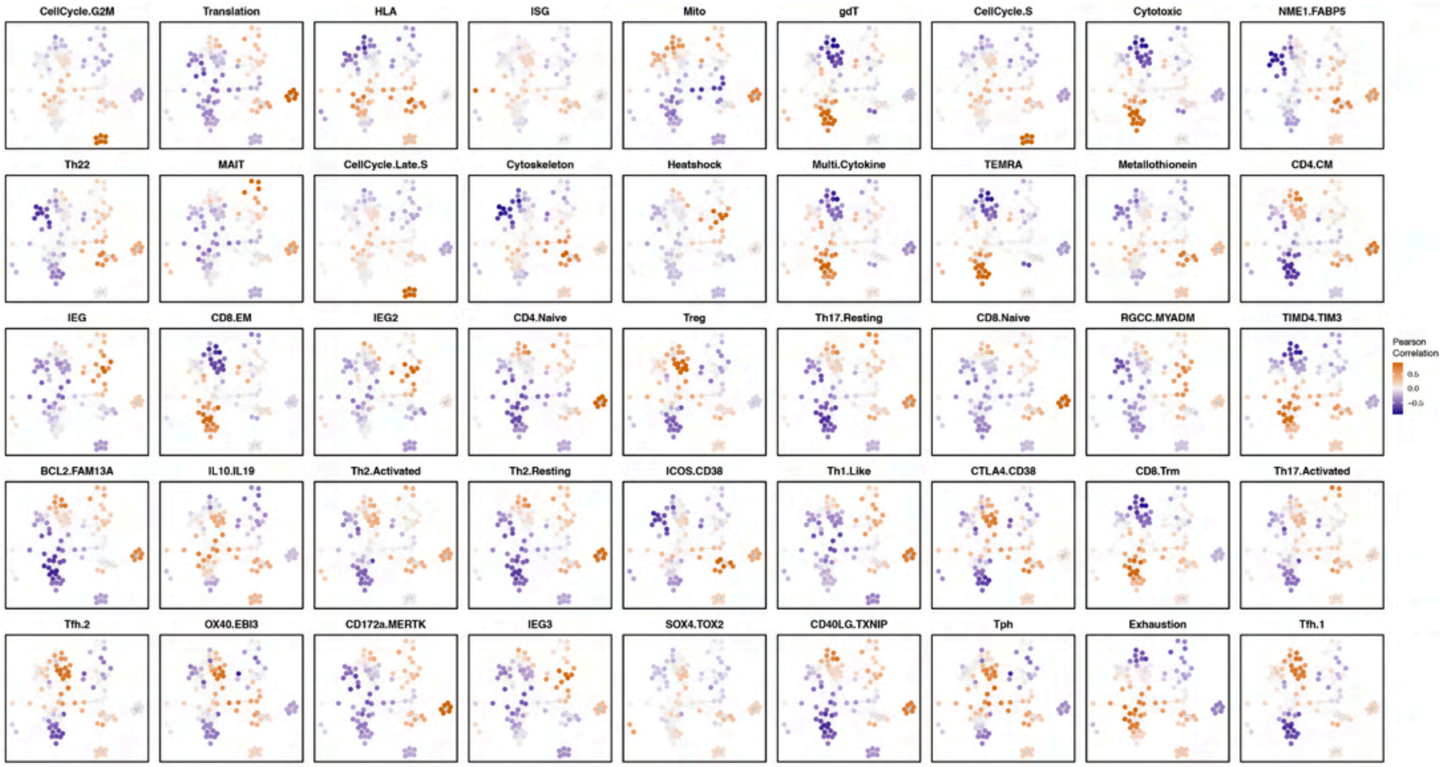


**Figure S20. Correlation of T-cell gene co-expression modules and cGEPs**

Network plots as in **Fig. 3a**, colored by gene expression correlations between co-expression modules and selected consensus gene expression programs (cGEPs)[8]. Networks show T-cell gene co-expression modules identified by hdWGCNA[7]. Nodes represent modules, and edges indicate similarity in expression profiles and gene-set overlap.

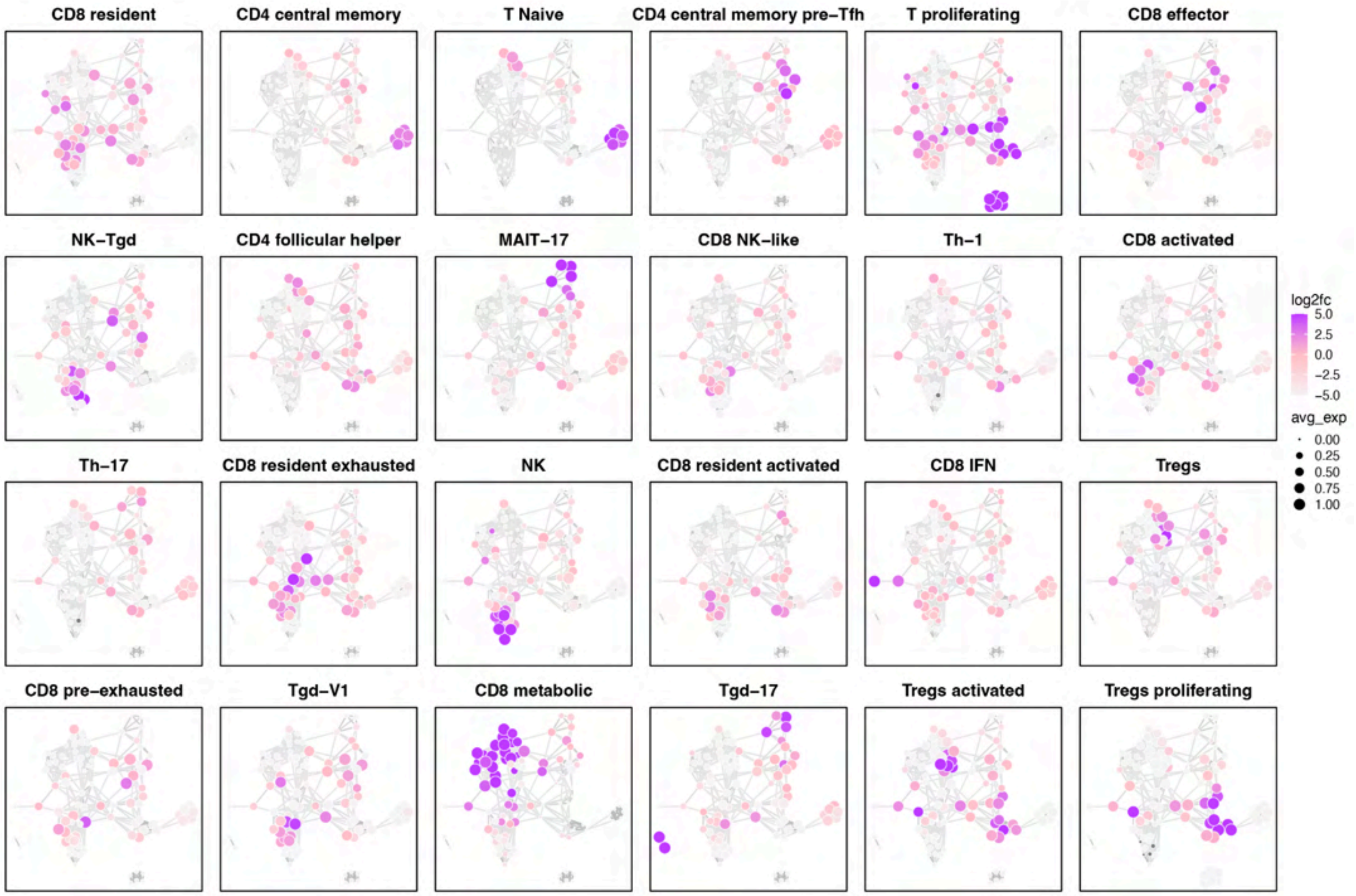


**Figure S21. Sub-type specific expression of T-cell co-expression modules**

Network plots as in **Fig. 3a**, showing the relative gene expression levels of each module for selected T-cell populations. Nodes are colored by avg. log2(fold-change) comparing one T-cell population against all others. Networks show T-cell gene co-expression modules identified by hdWGCNA[7]. Nodes represent modules, and edges indicate similarity in expression profiles and gene-set overlap. Node size is scaled by the average expression level of each module for the highlighted T-cell subtype

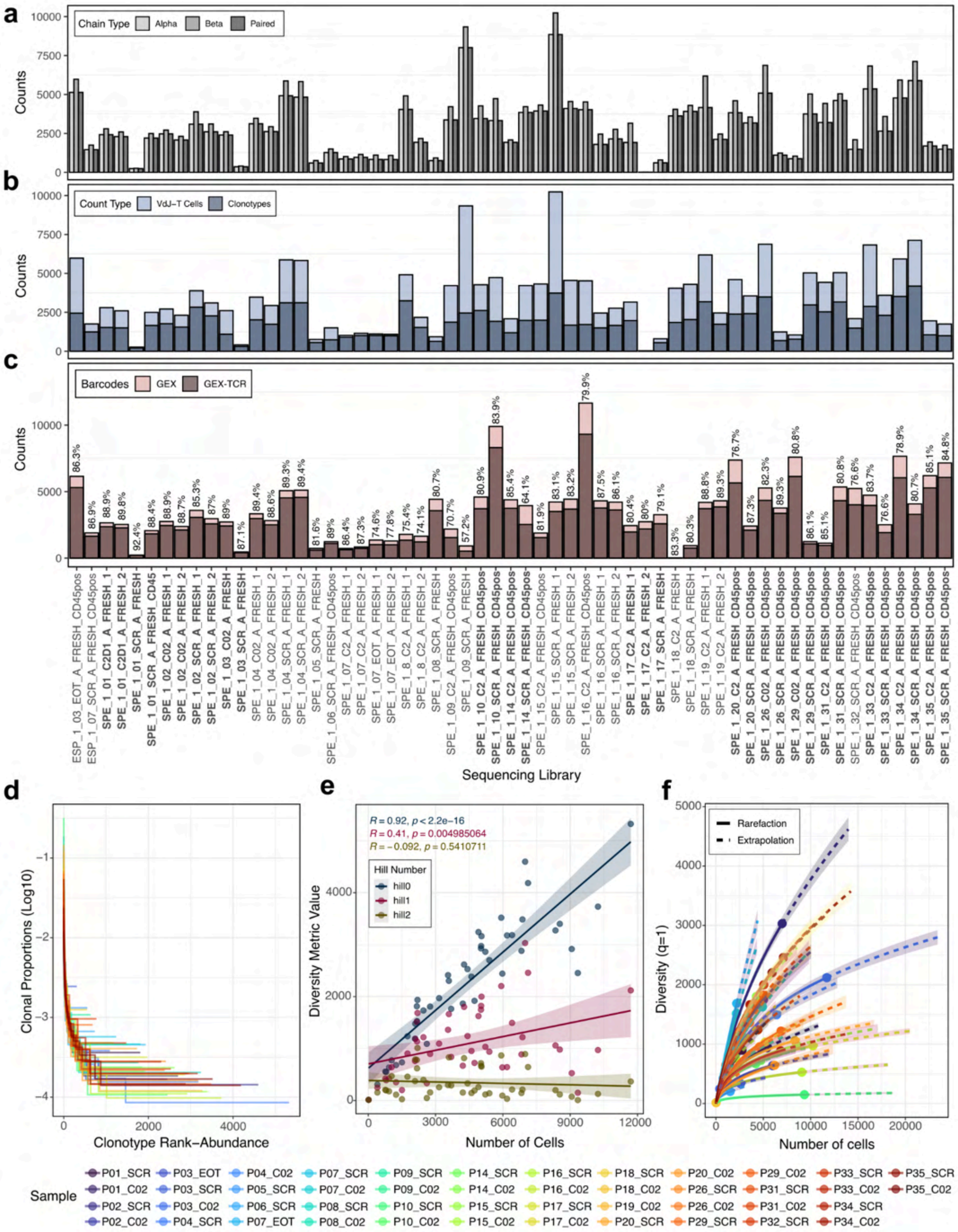
a
Chain Type
Alpha
Beta
Paired
Counts
0
2500
5000
7500
10000
b
Count Type
VdJ−T Cells
Clonotypes
Counts
0
2500
5000
7500
10000
c
Barcodes
GEX
GEX-TCR
Counts
0
5000
10000
86.3% 86.9% 88.9% 89.8% 92.4% 88.4% 88.9% 88.7% 85.3% 87% 89% 87.1% 89.4% 88.6% 89.3% 89.4% 81.6% 89% 86.4% 87.3% 74.6% 77.8% 75.4% 74.1% 80.7% 70.7% 57.2% 80.9% 83.9% 85.4% 64.1% 81.9% 83.1% 83.2% 79.9% 87.5% 86.1% 80.4% 80% 79.1% 83.3% 80.3% 88.8% 89.3% 76.7% 87.3% 82.3% 89.3% 80.8% 86.1% 85.1% 80.8% 76.6% 83.7% 76.6% 78.9% 80.7% 85.1% 84.8%
ESP_1_03_EOT_A_FRESH_CD45pos
ESP_1_07_SCR_A_FRESH_CD45pos
SPE_1_01_C2D1_A_FRESH_1
SPE_1_01_C2D1_A_FRESH_2
SPE_1_01_SCR_A_FRESH
SPE_1_01_SCR_A_FRESH_CD45
SPE_1_02_C02_A_FRESH_1
SPE_1_02_C02_A_FRESH_2
SPE_1_02_SCR_A_FRESH_1
SPE_1_02_SCR_A_FRESH_2
SPE_1_03_C02_A_FRESH
SPE_1_03_SCR_A_FRESH
SPE_1_04_C02_A_FRESH_1
SPE_1_04_C02_A_FRESH_2
SPE_1_04_SCR_A_FRESH_1
SPE_1_04_SCR_A_FRESH_2
SPE_1_05_SCR_A_FRESH
SPE_1_06_SCR_A_FRESH_CD45pos
SPE_1_07_C2_A_FRESH_1
SPE_1_07_C2_A_FRESH_2
SPE_1_07_EOT_A_FRESH_1
SPE_1_07_EOT_A_FRESH_2
SPE_1_8_C2_A_FRESH_1
SPE_1_8_C2_A_FRESH_2
SPE_1_08_SCR_A_FRESH
SPE_1_09_C2_A_FRESH_CD45pos
SPE_1_09_SCR_A_FRESH
SPE_1_10_C2_A_FRESH_CD45pos
SPE_1_10_SCR_A_FRESH_CD45pos
SPE_1_14_C2_A_FRESH_CD45pos
SPE_1_14_SCR_A_FRESH_CD45pos
SPE_1_15_C2_A_FRESH_CD45pos
SPE_1_15_SCR_A_FRESH_1
SPE_1_15_SCR_A_FRESH_2
SPE_1_16_C2_A_FRESH_CD45pos
SPE_1_16_SCR_A_FRESH_1
SPE_1_16_SCR_A_FRESH_2
SPE_1_17_C2_A_FRESH_1
SPE_1_17_C2_A_FRESH_2
SPE_1_17_SCR_A_FRESH
SPE_1_18_C2_A_FRESH
SPE_1_18_SCR_A_FRESH
SPE_1_19_C2_A_FRESH_1
SPE_1_19_C2_A_FRESH_2
SPE_1_20_C2_A_FRESH_CD45pos
SPE_1_20_SCR_A_FRESH_CD45pos
SPE_1_26_C02_A_FRESH_CD45pos
SPE_1_26_SCR_A_FRESH_CD45pos
SPE_1_29_C02_A_FRESH_CD45pos
SPE_1_29_SCR_A_FRESH_CD45pos
SPE_1_31_C2_A_FRESH_CD45pos
SPE_1_31_SCR_A_FRESH_CD45pos
SPE_1_32_SCR_A_FRESH_CD45pos
SPE_1_33_C2_A_FRESH_CD45pos
SPE_1_33_SCR_A_FRESH_CD45pos
SPE_1_34_C2_A_FRESH_CD45pos
SPE_1_34_SCR_A_FRESH_CD45pos
SPE_1_35_C2_A_FRESH_CD45pos
SPE_1_35_SCR_A_FRESH_CD45pos
Sequencing Library
d
Clonal Proportions (Log10)
−1
−2
−3
−4
0
2000
4000
Clonotype Rank−Abundance
e
R = 0.92, p < 2.2e−16
R = 0.41, p = 0.004985064
R = −0.092, p = 0.5410711
Hill Number
hill0
hill1
hill2
Diversity Metric Value
0
2000
4000
0
3000
6000
9000
12000
Number of Cells
f
Rarefaction
Extrapolation
Diversity (q=1)
0
1000
2000
3000
4000
5000
0
5000
10000
15000
20000
Number of cells
Sample
P01_SCR P01_C02 P02_SCR P02_C02
P03_EOT P03_SCR P03_C02 P04_SCR
P04_C02 P05_SCR P06_SCR P07_EOT
P07_SCR P07_C02 P08_SCR P08_C02
P09_SCR P09_C02 P10_SCR P10_C02
P14_SCR P14_C02 P15_SCR P15_C02
P16_SCR P16_C02 P17_SCR P17_C02
P18_SCR P18_C02 P19_C02 P20_SCR
P20_C02 P26_SCR P26_C02 P29_SCR
P29_C02 P31_SCR P31_C02 P32_SCR
P33_SCR P33_C02 P34_SCR P34_C02
P35_SCR P35_C02

**Figure S22. Quality control and depth-dependent bias assessment of tumor 10x Genomics VDJ-T sequencing and TCR repertoire diversity**

**a**, Number of cells with a recovered TCR alpha chain (light gray), beta chain (gray), or both paired chains (dark gray) for each sequencing library. Library names in bold (x-axis, shared across **a-c**) indicate libraries belonging to the MSS CRC liver/lung metastasis cohort used in downstream analyses. **b**, Number of cells with a recovered beta chain, used to define clonotypes (light blue), and the corresponding number of unique clonotypes (dark blue) per sequencing library. **c**, For paired GEX-TCR libraries, total number of barcodes annotated as T cells by gene expression (GEX, pink) and the subset of these with matched TCR information (GEX-TCR, brown). Percentages above each bar indicate the percentage of GEX-annotated T cells with a recovered TCR. **d**, Clonal proportion (rank-abundance) curves showing the normalized frequency of each clonotype (log10 scale) ranked by abundance, per biological sample. Technical replicate libraries were aggregated into a single biological sample prior to this and all subsequent analyses (**e, f**). **e**, Spearman correlation between the number of cells recovered per sample and raw diversity, computed at Hill number orders q=0 (Species richness), q=1 (Shannon diversity: exponential of Shannon entropy), and q=2 (Simpson diversity: inverse Simpson index), illustrating the dependence of diversity estimates on sample size and justifying the use of bootstrapped downsampling to a common depth in subsequent diversity analyses (see **Methods**). Spearman's ρ and *p*-values are shown for each diversity metric. **f**, Sample-level rarefaction (solid lines) and extrapolation (dashed lines) curves of TCR repertoire diversity (Shannon diversity, q = 1) as a function of the number of sampled cells. Shaded areas represent 95% confidence intervals; dots indicate the observed sample size and corresponding diversity value. Sample color legend applies to panels **d–f**; sample codes indicate patient ID and timepoint (SCR = screening/pre-treatment, C02 = on-treatment cycle 2, EOT = end of treatment).

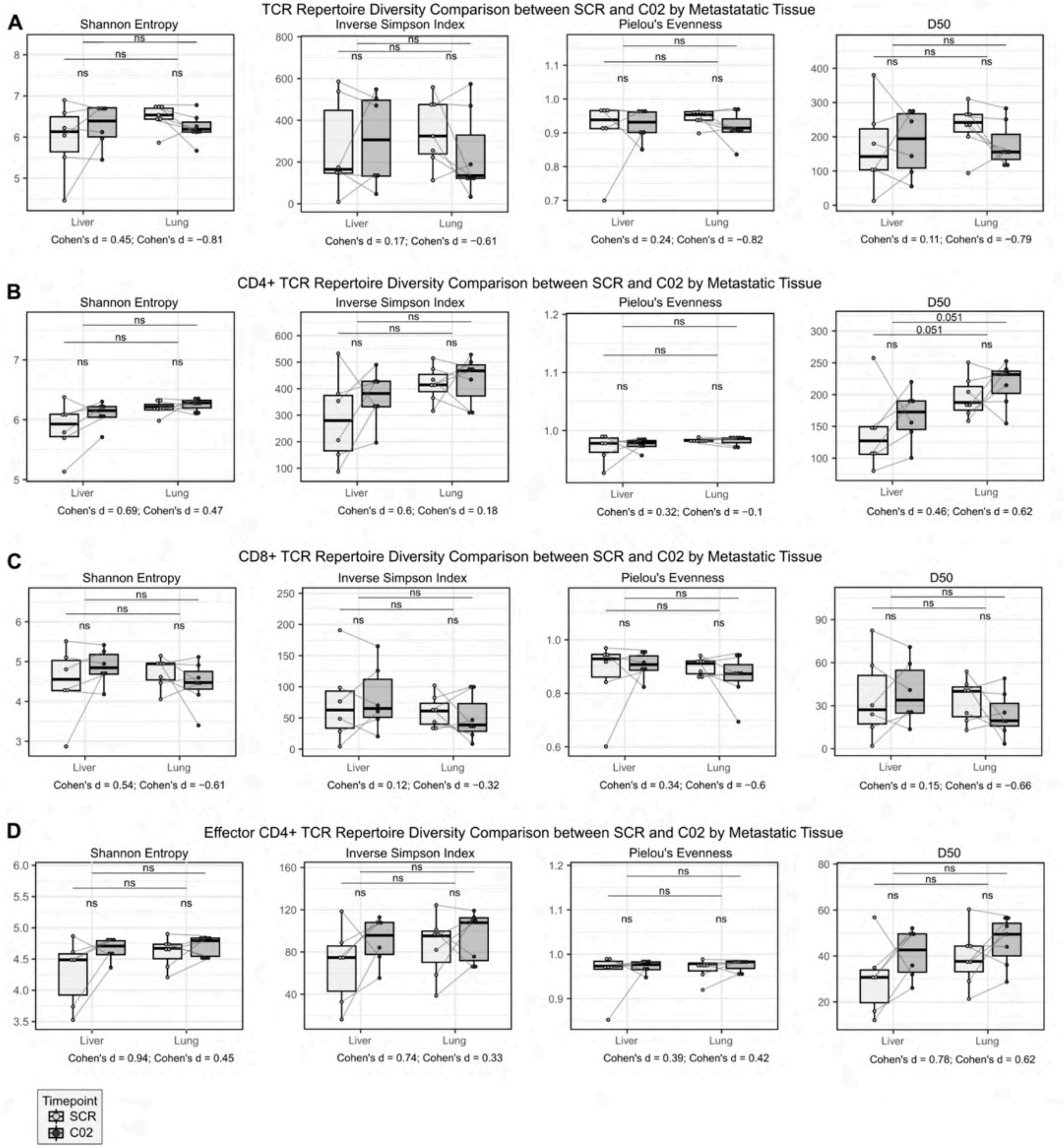


**Figure S23. Extended TCR repertoire diversity comparison between SCR and C02 timepoints split by metastatic site**

Figure caption on the next page.

**Figure S23. Tumor TCR repertoire diversity comparison between SCR and C02 timepoints split by metastatic site**

**A-D,** Comparison of TCR repertoire diversity metrics (Shannon entropy, Inverse Simpson Index, Pielou's Evenness, and the number of top clones making up 50% of the library [D50]) between SCR (screening, pre-treatment) and C02 (2nd cycle on treatment, post-treatment) timepoints across all TCR+ cells (**A**), CD4+ TCR+ cells (**B**), CD8+ TCR+ cells (**C**), and effector non-Treg CD4+ TCR+ cells (**D**), and stratified by liver (n=6) and lung metastases (n=7). Each dot represents the mean diversity metric per patient with lines connecting the same patient across time points, calculated from 1,000 bootstrap iterations with downsampling to a common depth (10th percentile of sample sizes [1,667 cells (**A**); 743 cells (**B**); 431 cells (**C**); 651 cells (**D**)]). Effect sizes are reported as Cohen's d for each paired comparison; p-values from two-sided Wilcoxon signed-rank tests from paired (SCR vs. C02), and from two-sided Wilcoxon rank-sum tests from timepoint-specific unpaired (liver vs. lung) are shown. Non-significant comparisons are labeled "ns"; comparisons with $p < 0.1$ display the exact p-value; significant comparisons ($p < 0.05$) are shown in bold.

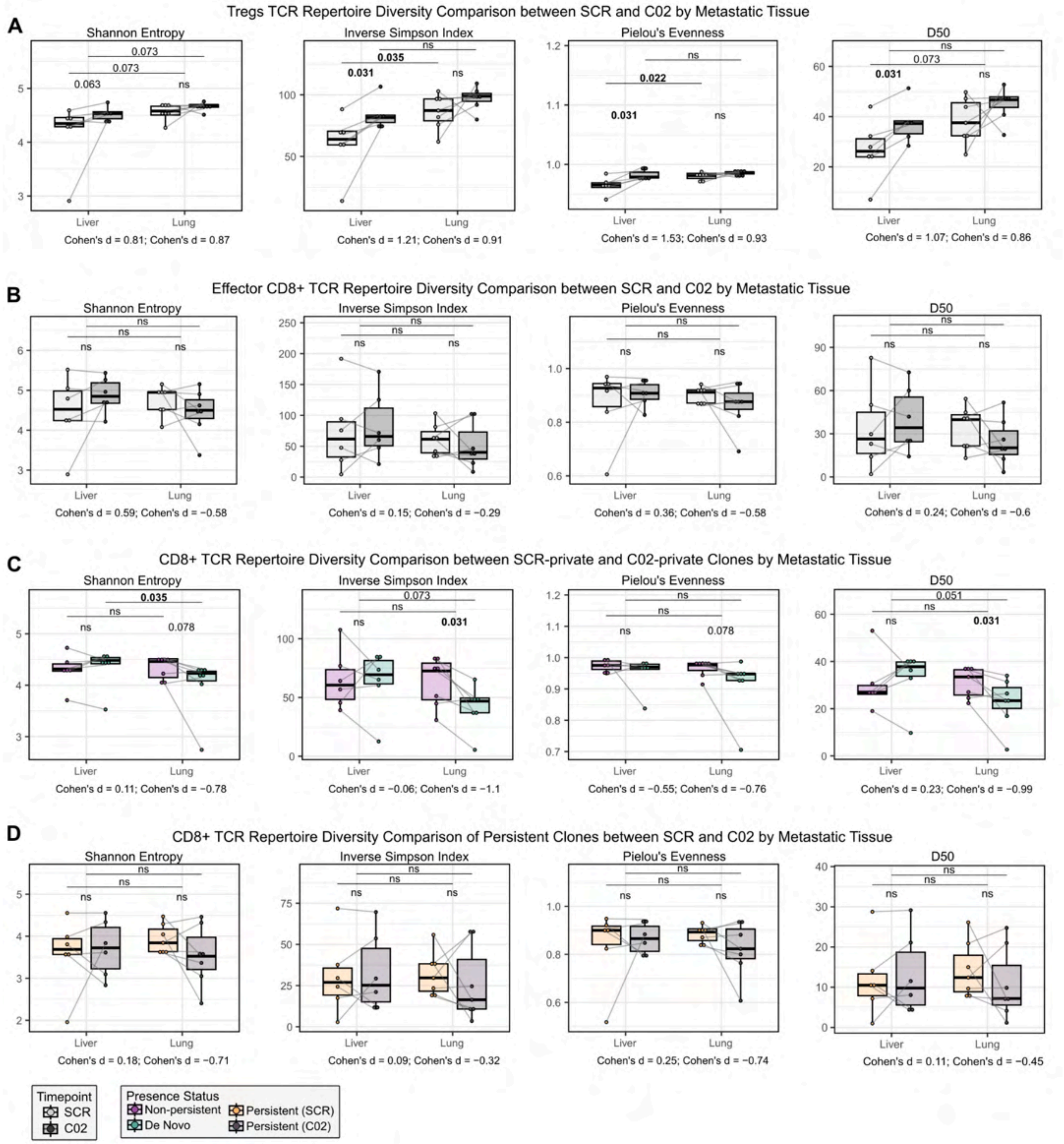


**Figure S24. Extended TCR repertoire diversity comparison between SCR and C02 timepoints split by metastatic site**

Figure caption on the next page.

**Figure S24. Extended TCR repertoire diversity comparison between SCR and C02 timepoints split by metastatic site**

**A-D,** Comparison of TCR repertoire diversity metrics (Shannon entropy, Inverse Simpson Index, Pielou's Evenness, and the number of top clones making up 50% of the library [D50]) between SCR (screening, pre-treatment) and C02 (2nd cycle on treatment, post-treatment) timepoints across Treg TCR+ cells (**A**), effector CD8+ TCR+ cells (**B**), timepoint-private CD8+ TCR+ cells (non-persistent vs. *de novo*; **C**), and persistent CD8+ TCR+ cells (**D**), and stratified between liver (n=6) and lung metastases (n=7). Each dot represents the mean diversity metric per patient with lines connecting the same patient across time points, calculated from 1,000 bootstrap iterations with downsampling to a common depth (10th percentile of sample sizes [139 cells (**A**); 431 cells (**B**); 130 cells (**C**); 239 cells (**D**)]). Effect sizes are reported as Cohen's d for each paired comparison; p-values from two-sided Wilcoxon signed-rank tests from paired (SCR vs. C02), and from two-sided Wilcoxon rank-sum tests from timepoint-specific unpaired (liver vs. lung) are shown. Non-significant comparisons are labeled "ns"; comparisons with $p < 0.1$ display the exact p-value; significant comparisons ($p < 0.05$) are shown in bold.

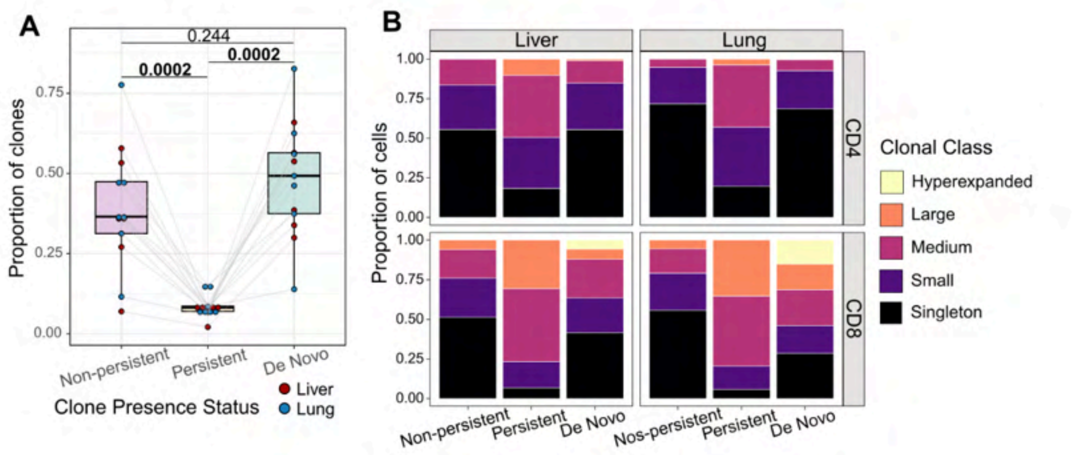


**Figure S25. Extended characterization of tumor TCR repertoire clonal structure in liver and lung metastases**

**a**, Patient-level proportions of tumor non-persistent, persistent and *de novo* clones within their paired SCR-C02 TCR repertoires. Dots are colored by metastatic sites (liver = red, lung = blue) with lines connecting the same patient across presence statuses. Pairwise two-sided Wilcoxon signed-rank tests p-values are shown. **b**, Stacked barplots showing the global proportions of clonal expansion classes across non-persistent, persistent at SCR, persistent at C02, and *de novo* clones faceted by metastatic site and T-cell lineage.

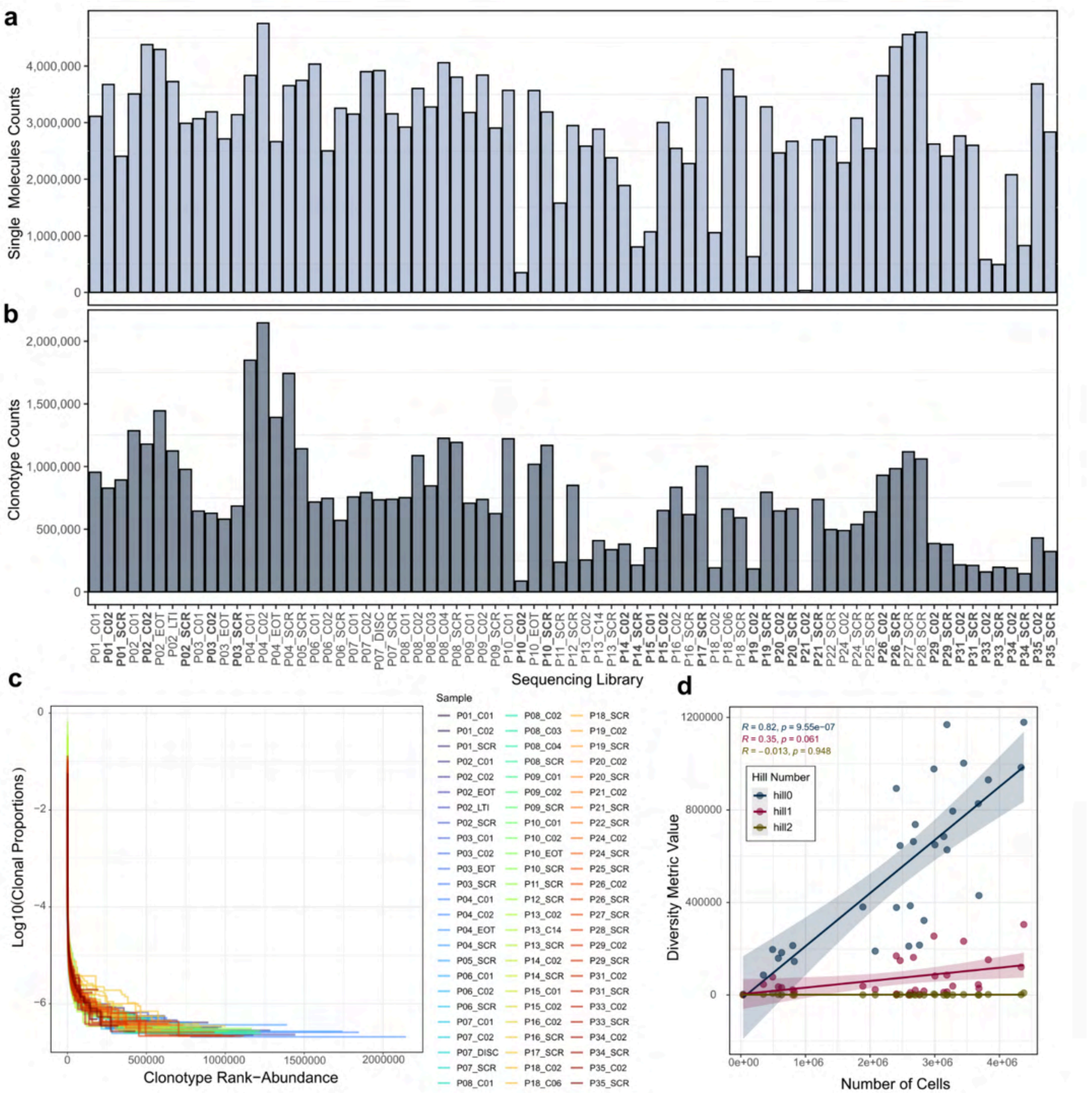


**Figure S26. Quality control and depth-dependent bias assessment of blood OS-TCR VDJ-T sequencing and TCR repertoire diversity**

Figure caption on the next page.

**Figure S26. Quality control and depth-dependent bias assessment of blood OS-TCR VDJ-T sequencing and TCR repertoire diversity**

**a**, Number of sequenced single molecules per OS-TCR sequencing library. **b**, Number of sequenced clonotypes per OS-TCR sequencing library. **c**, Clonal proportion (rank-abundance) curves showing the normalized frequency of each clonotype (log10 scale) ranked by abundance, per library. **d**, Spearman correlation between the number of cells recovered per samples used in downstream analyses and raw diversity, computed at Hill number orders q=0 (Species richness), q=1 (Shannon diversity: exponential of Shannon entropy), and q=2 (Simpson diversity: inverse Simpson index). Spearman's ρ and *p*-values are shown for each diversity metric.

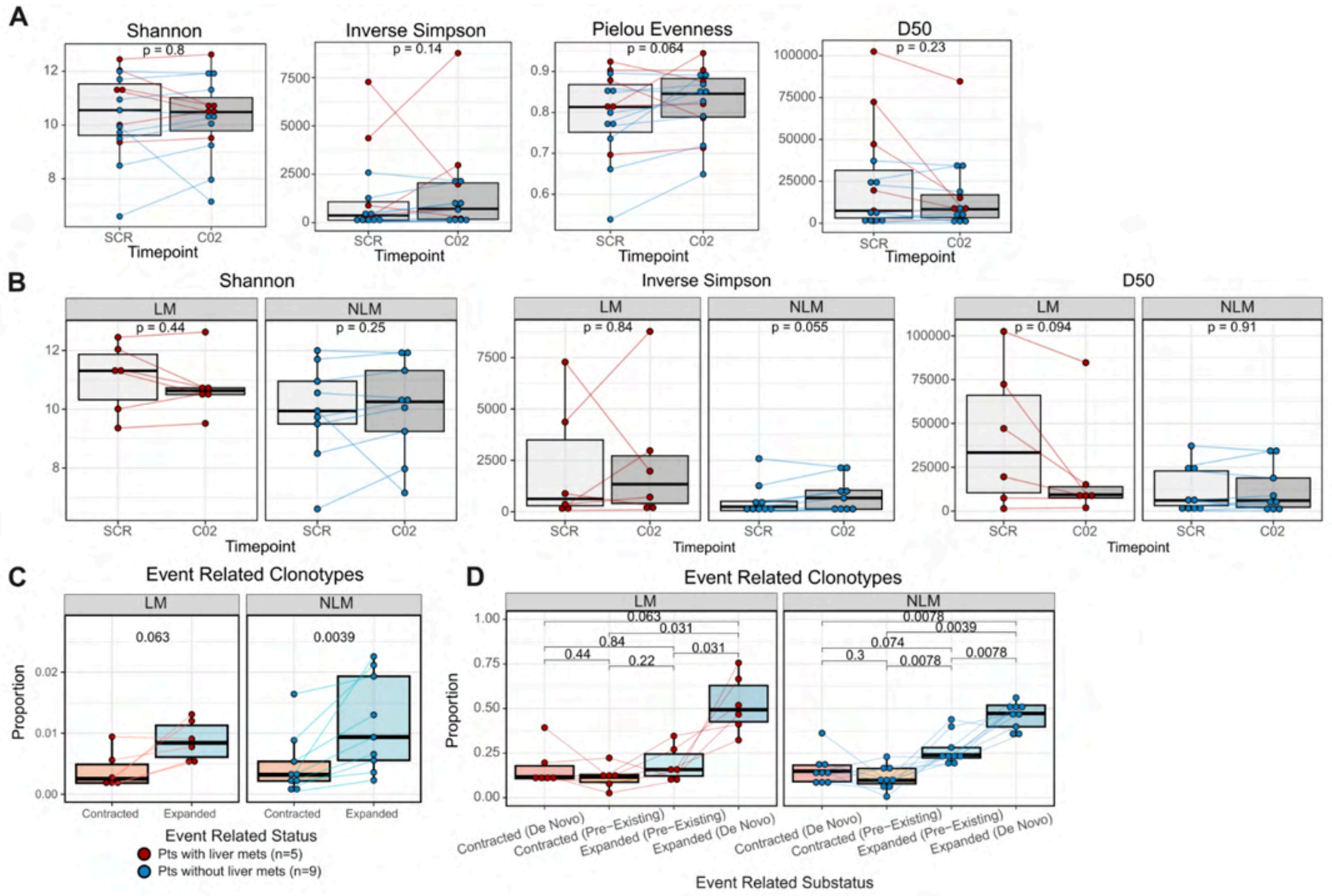


**Figure S27. Extended peripheral blood ICI-induced clonal dynamics**

**A,B,** Comparison of peripheral blood OS-TCR repertoire diversity metrics [Shannon Entropy (Shannon), Inverse Simpson Index (Inverse Simpson), Pielou's Evenness (Pielou Evenness), and the number of top clones making up 50% of the library (D50)] between SCR (screening, pre-treatment) and C02 (2nd cycle on treatment, post-treatment) timepoints at the global level (**A**) and split by patients with (n=6) and without (n=9) liver metastases (**B**). Each dot represents the diversity metric per patient, with lines connecting the same patient across time points. Dots are colored by whether patients have liver (red) or lung metastases (blue). P-values from two-sided Wilcoxon signed-rank tests are shown. **C,** Proportion of event-related clonotypes (contracted, expanded) within the peripheral blood OS-TCR repertoires computed with a proprietary noise model (Omniscope Inc., Spain) using a $\log_2$ fold change ≥ 2 between time points to assess event relatedness. Each dot represents the proportion for each patient, with lines connecting the same patient across time points, and are colored by whether they have liver (red) metastases or not (and have lung metastases; blue). Boxplots are split between patients with (n=6) and without liver metastases (n=9). P-values from two-sided Wilcoxon signed-rank tests are shown. **D,** Proportion of pre-existing and *de novo* event-related clonotypes within the peripheral blood OS-TCR event-related pool. Each dot represents the proportion for each patient, and are colored by whether they have liver metastases (red) or not (and have lung metastases; blue). Boxplots are split between patients with (n=6) and without liver metastases (n=9). P-values from two-sided Wilcoxon signed-rank tests are shown.

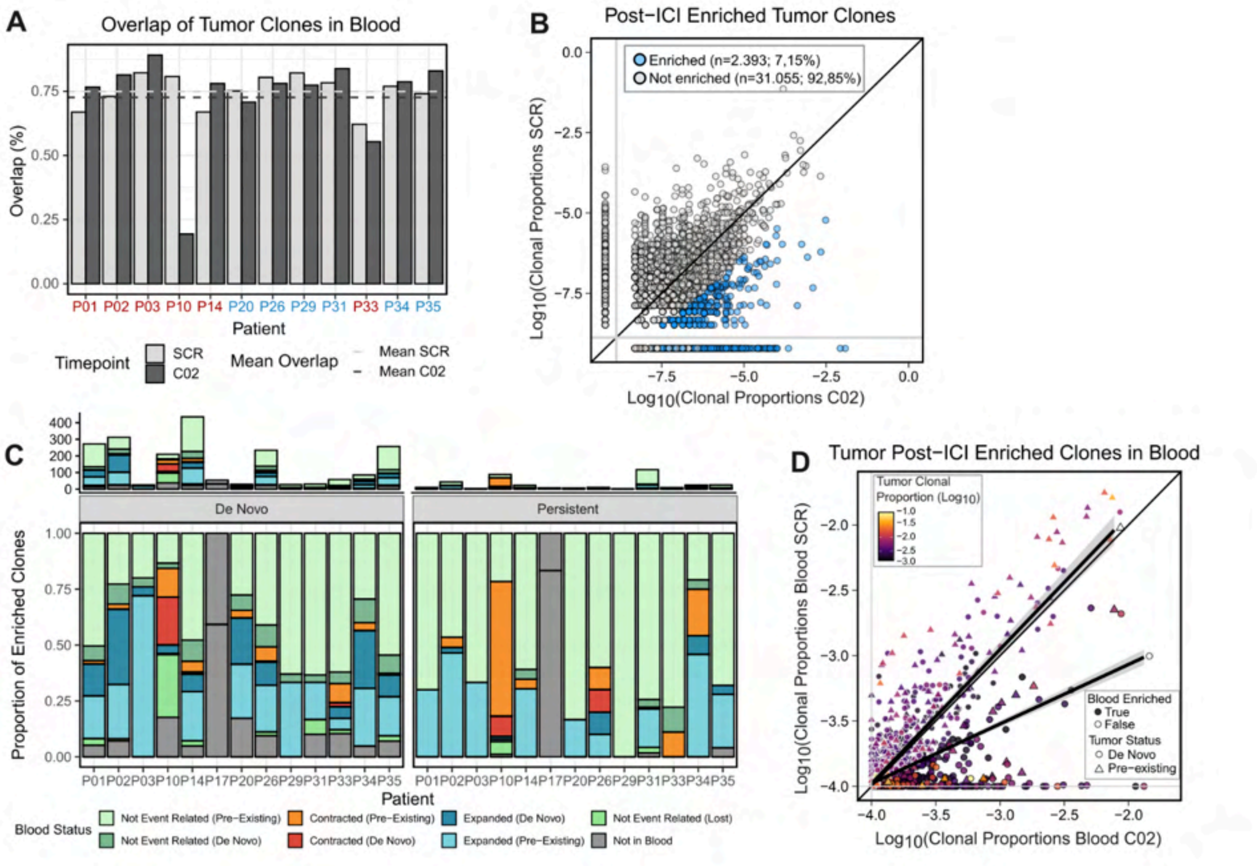


**Figure S28. Extended systems tracking of therapy-specific clones**

**A,** Barplot showing the timepoint-specific overlap (in %) between tumor and blood TCR repertoires (SCR, pre-treatment in light gray; C02, post-treatment in dark gray) for each patient with matched 10x tumor and blood OS-TCR SCR and C02 data. The mean for each timepoint is represented as a dashed horizontal line. Patients with liver metastases are colored in red and patients without liver but with lung metastases are colored in blue. **B,** Clonal scatter of tumor pre-treatment (SCR, y axis) and post-treatment (C02, x axis) clonotype expansion levels (clonal proportions in log10 scale). Each dot represents a clonotype and is colored by whether it is enriched post-treatment (post-ICI; blue) or not (gray). Enrichment is determined by whether the clonotype has a clonal proportion with a $\log_2$ fold change ≥ 2 for persistent clones, and by whether the clonotype has a clonal proportion greater than the 90th percentile of the patient's-specific non-persistent pool for *de novo* clones. **C,** Frequency (top) and proportions stacked barplots (bottom) showing blood event related status in blood of tumor post-ICI clones at the patient level (x axis) split by presence status in the tumor (*de novo* and persistent [Pre-Existing]). **D,** Clonal scatter of blood pre-treatment (SCR, y axis) and post-treatment (C02, x axis) clonotype expansion levels (clonal proportions in log10 scale) of tumor post-ICI enriched clonotypes. Each dot is a tumor clonotype that is matched in peripheral blood in both SCR and CO2 timepoints, and is colored by clonal expansion in the tumor post-treatment (clonal proportions in log10 scale), being yellow greater expansion and dark purple lower expansion. Triangle-shaped dots clonotypes represent tumor persistent clones (pre-existing) and circle-shaped clonotypes represent tumor *de novo* clones. Black-edged clonotypes represent tumor post-ICI enriched clones and white-edged clonotypes represent non-enriched tumor clones.

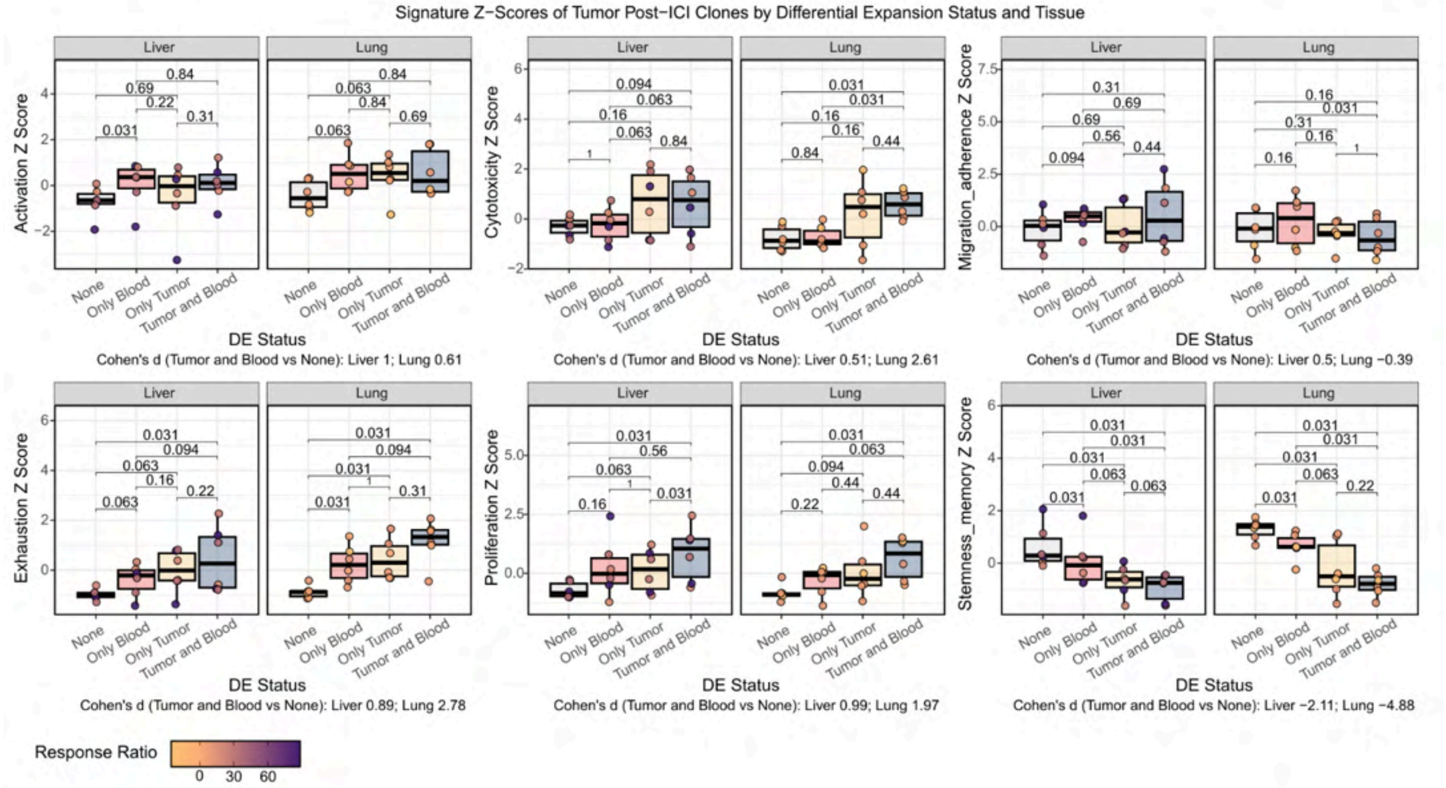


**Figure S29. T-cell function gene set signatures assessment in tumor post-ICI clones based on their differential expansion status in tumor and peripheral blood compartments**

Z-scores of gene signatures computed with AUCell on post-ICI (C02) tumor TRC+ cells stratified by their differential expansion status (DE status) in tumor and peripheral blood (Tumor and Blood: differentially expanded (DE) in both tumor and blood compartments; Only Tumor: DE in the tumor but not in blood; Only Blood: DE in blood but not in the tumor; None: not DE in any compartment) and split by liver (n=6) and lung (n=6) metastatic tissues. Each dot corresponds to each patient's mean Z-scaled AUCell score (scores are scaled across patients and DE Status groups) and is colored by tumor growth after treatment (response ratio), being dark purple the worst response and light orange the best response. P-values of pairwise paired statistical significance assessment through two-sided Wilcoxon signed-rank tests across DE Status groups are shown.

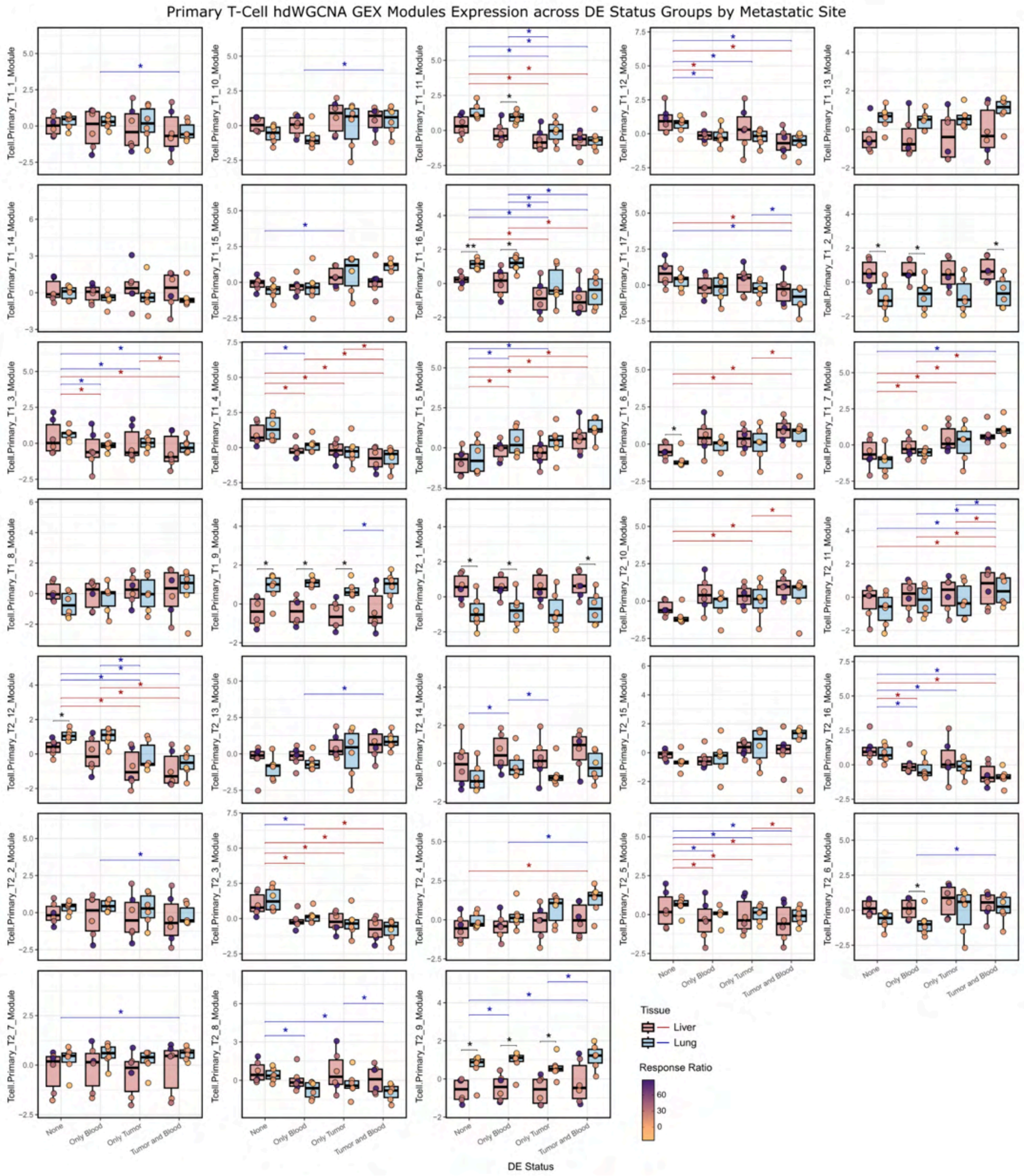


**Figure S30. Primary T-cell co-expression modules expression on tumor post-ICI TCR+ cells stratified by differential expansion status groups and split by metastatic site**

Figure caption on the next page.

**Figure S30. Primary T-cell co-expression modules expression on tumor post-ICI TCR+ cells stratified by differential expansion status groups and split by metastatic site**

Primary-derived T-cell modules ULM Z-Scores on patient-sample pseudobulks of post-ICI tumor TCR+ cells and stratified by tumor and blood differential expansion (DE) status (Tumor and Blood: differentially expanded (DE) in both tumor and blood compartments; Only Tumor: DE in the tumor but not in blood; Only Blood: DE in blood but not in the tumor; None: not DE in any compartment) and metastatic site (liver in red, lung in blue). Patient's dots are colored by tumor growth (response ratio) where orange indicates best responses and purple worse responses. Unpaired inter-tissue differences are tested through two-sided Wilcoxon rank-sum tests for each DE status group, and significant differences ($p < 0.05$) are shown with an asterisk (*). Paired intra-tissue differences are tested through two-sided Wilcoxon signed-rank tests for each metastatic site, and significant paired differences ($p < 0.05$) are additionally shown for both metastatic sites (blue lines and asterisks for lung mets, red lines and asterisk for liver mets).

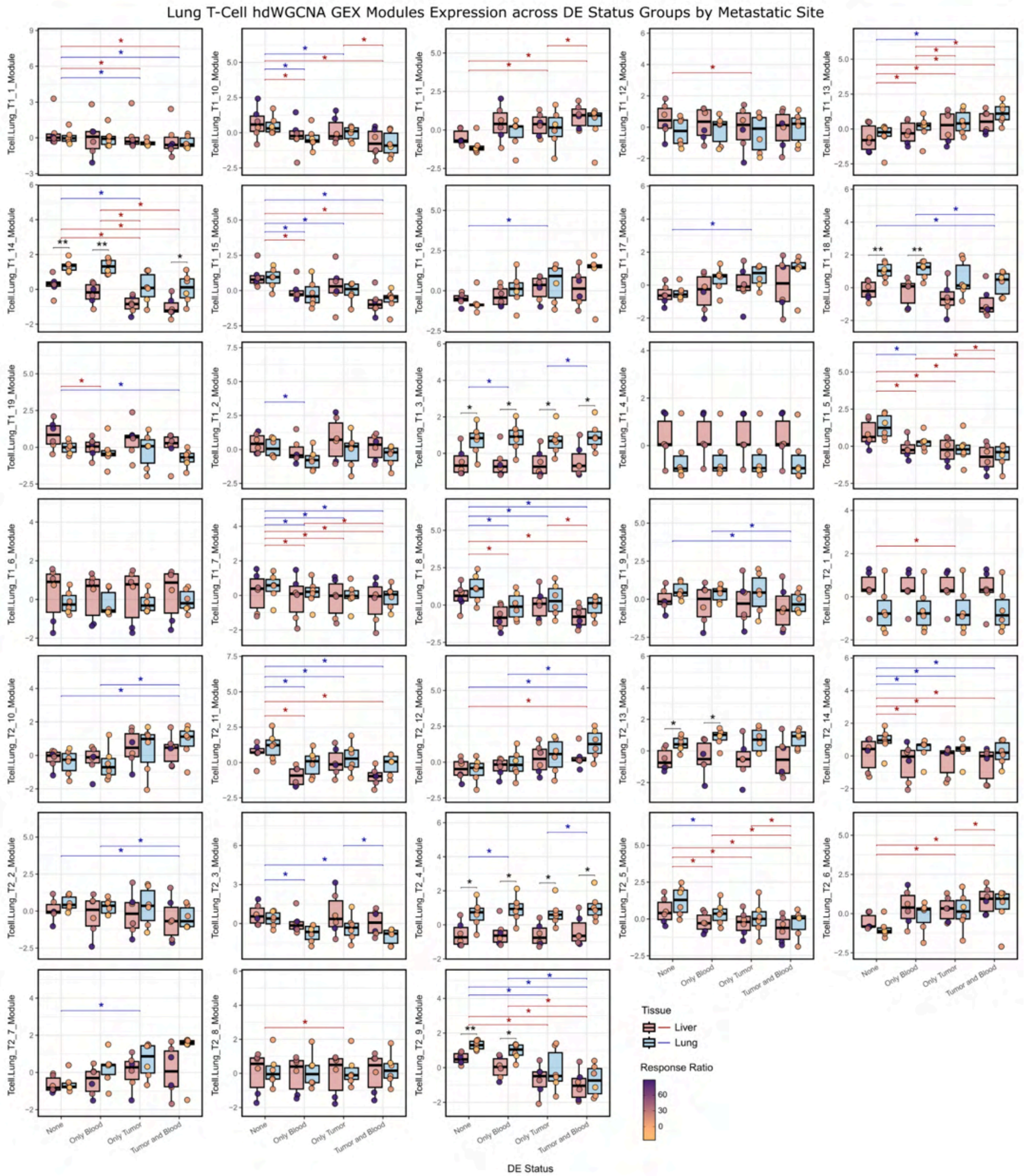


**Figure S31. Lung T-cell co-expression modules expression on tumor post-ICI TCR+ cells stratified by differential expansion status groups and split by metastatic site**

Figure caption on the next page.

**Figure S31. Lung T-cell co-expression modules expression on tumor post-ICI TCR+ cells stratified by differential expansion status groups and split by metastatic site**

Lung-derived T-cell modules ULM Z-Scores on patient-sample pseudobulks of post-ICI tumor TCR+ cells and stratified by tumor and blood differential expansion (DE) status (Tumor and Blood: differentially expanded (DE) in both tumor and blood compartments; Only Tumor: DE in the tumor but not in blood; Only Blood: DE in blood but not in the tumor; None: not DE in any compartment) and metastatic site (liver in red, lung in blue). Patient's dots are colored by tumor growth (response ratio) where orange indicates best responses and purple worse responses. Unpaired inter-tissue differences are tested through two-sided Wilcoxon rank-sum tests for each DE status group, and significant differences ($p < 0.05$) are shown with an asterisk (*). Paired intra-tissue differences are tested through two-sided Wilcoxon signed-rank tests for each metastatic site, and significant paired differences ($p < 0.05$) are additionally shown for both metastatic sites (blue lines and asterisks for lung mets, red lines and asterisk for liver mets.

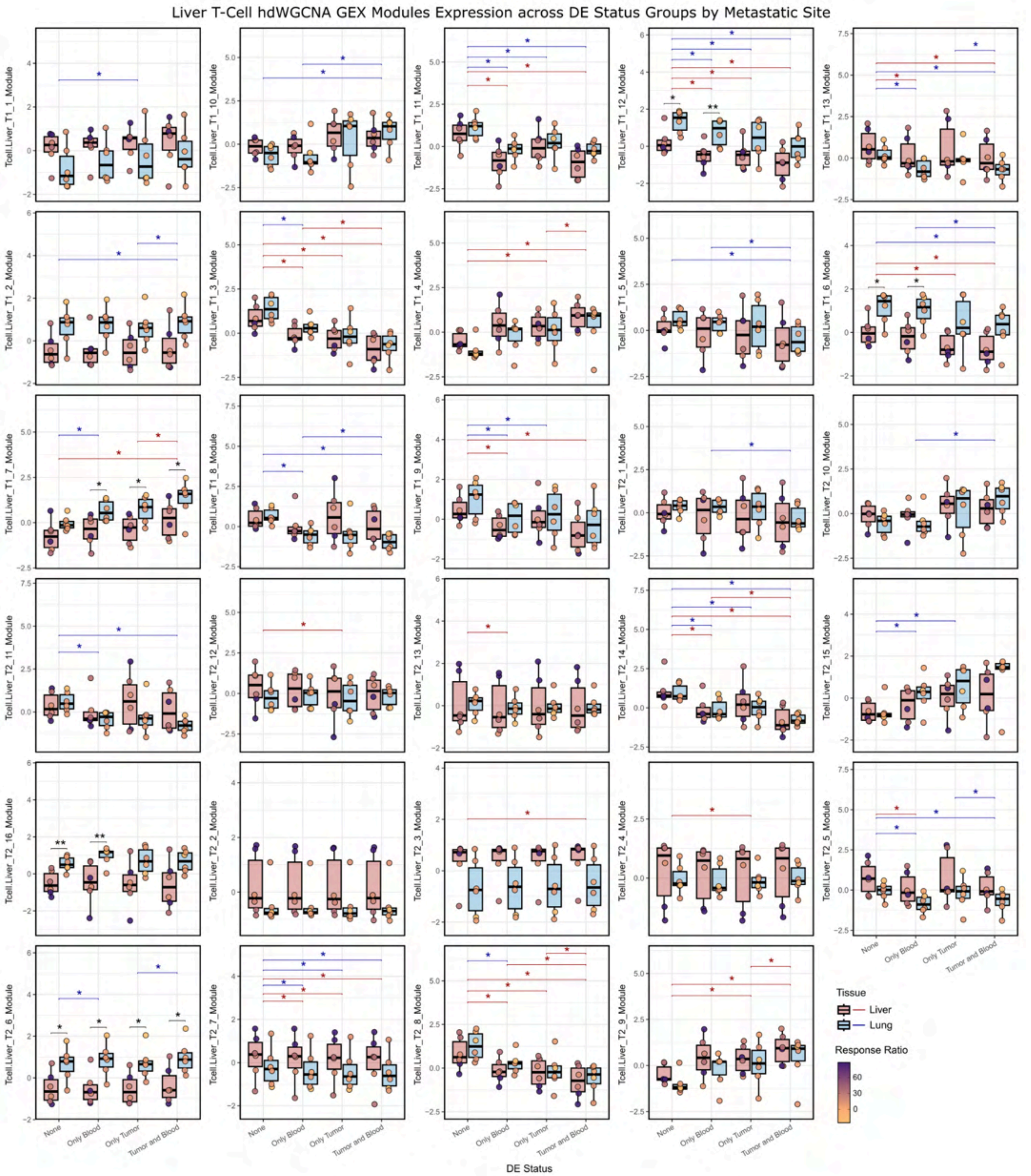


**Figure S32. Liver T-cell co-expression modules expression on tumor post-ICI TCR+ cells stratified by differential expansion status groups and split by metastatic site**

Figure caption on the next page.

**Figure S32. Liver T-cell co-expression modules expression on tumor post-ICI TCR+ cells stratified by differential expansion status groups and split by metastatic site**

Liver-derived T-cell modules ULM Z-Scores on patient-sample pseudobulks of post-ICI tumor TCR+ cells and stratified by tumor and blood differential expansion (DE) status (Tumor and Blood: differentially expanded (DE) in both tumor and blood compartments; Only Tumor: DE in the tumor but not in blood; Only Blood: DE in blood but not in the tumor; None: not DE in any compartment) and metastatic site (liver in red, lung in blue). Patient's dots are colored by tumor growth (response ratio) where orange indicates best responses and purple worse responses. Unpaired inter-tissue differences are tested through two-sided Wilcoxon rank-sum tests for each DE status group, and significant differences ($p < 0.05$) are shown with an asterisk (*). Paired intra-tissue differences are tested through two-sided Wilcoxon signed-rank tests for each metastatic site, and significant paired differences ($p < 0.05$) are additionally shown for both metastatic sites (blue lines and asterisks for lung mets, red lines and asterisk for liver mets.

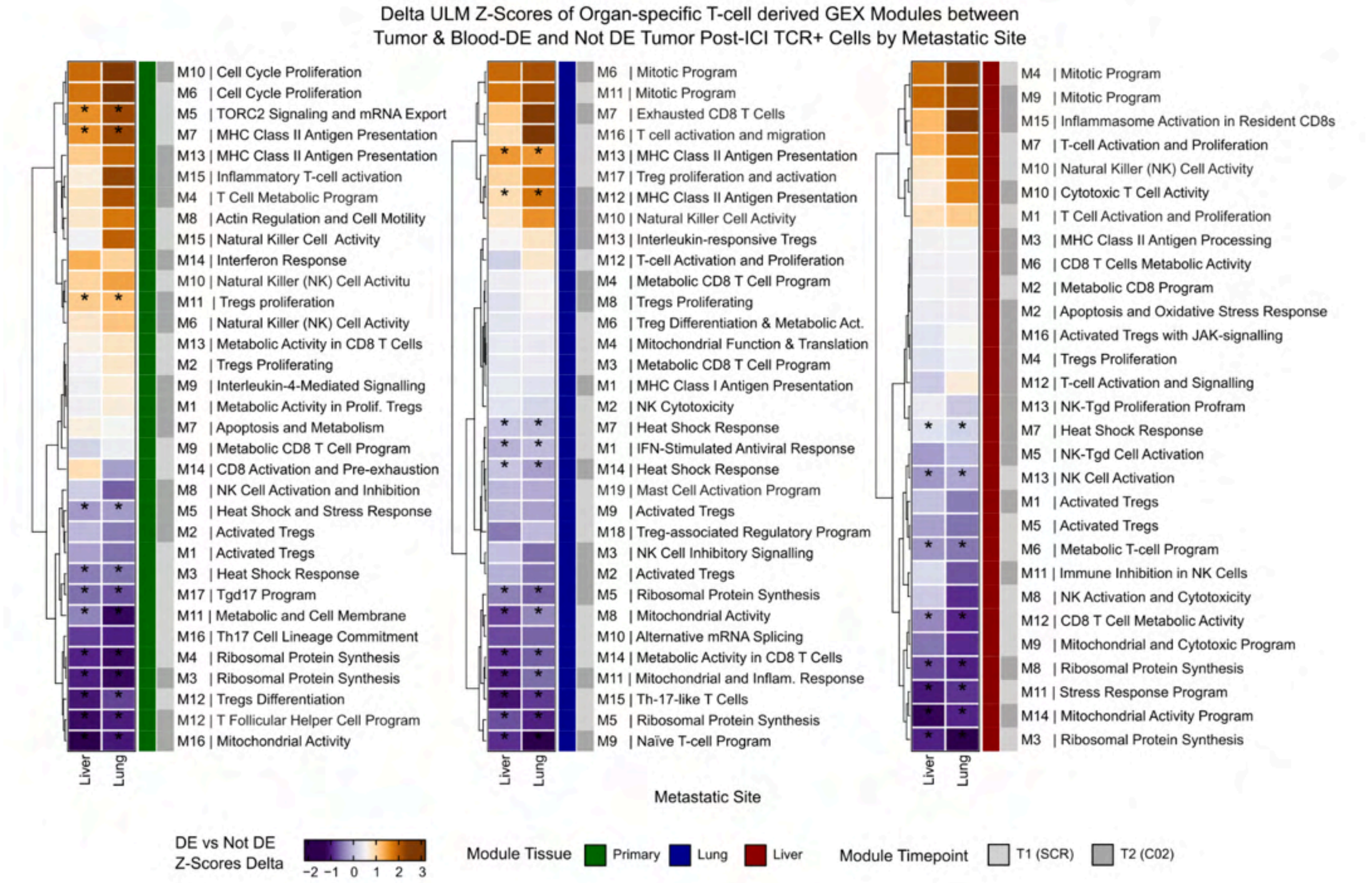


**Figure S33. Delta ULM Z-Scores of Organ-specific T-cell derived GEX Modules between Tumor and Blood DE and Not DE tumor post-ICI TCR+ cells by metastatic site**

Heatmaps showing the delta between Tumor-and-Blood DE and Not DE Status groups of ULM Z-scaled scores of Organ-derived (Primary CRC in green, Lung mets in blue, Liver mets in red) and timepoint-specific (SCR in light gray, C02 in dark gray) GEX modules computed with hdWGCNA from the T-cell scRNA-seq compartment on the tumor post-ICI (C02) TCR+ cells split by metastatic site (liver and lung; x axis). Positive Z-scores delta values shown in orange indicate higher expression of the module in the Tumor and Blood DE Status group, while negative values shown in purple indicate higher expression in the None DE Status group. Modules are hierarchically clustered based on their deltas across liver and lung mets (y axis). Modules are functionally annotated with *Llegir* (see **Methods**). Statistical significance of paired differences between both DE Status groups was assessed through two-sided Wilcoxon signed-rank tests and corrected for multiple testing via the Bonferroni-Hochberg method. The significance threshold was set at 0.05 and significant differences for each module and tissue are shown with an asterisk (*).

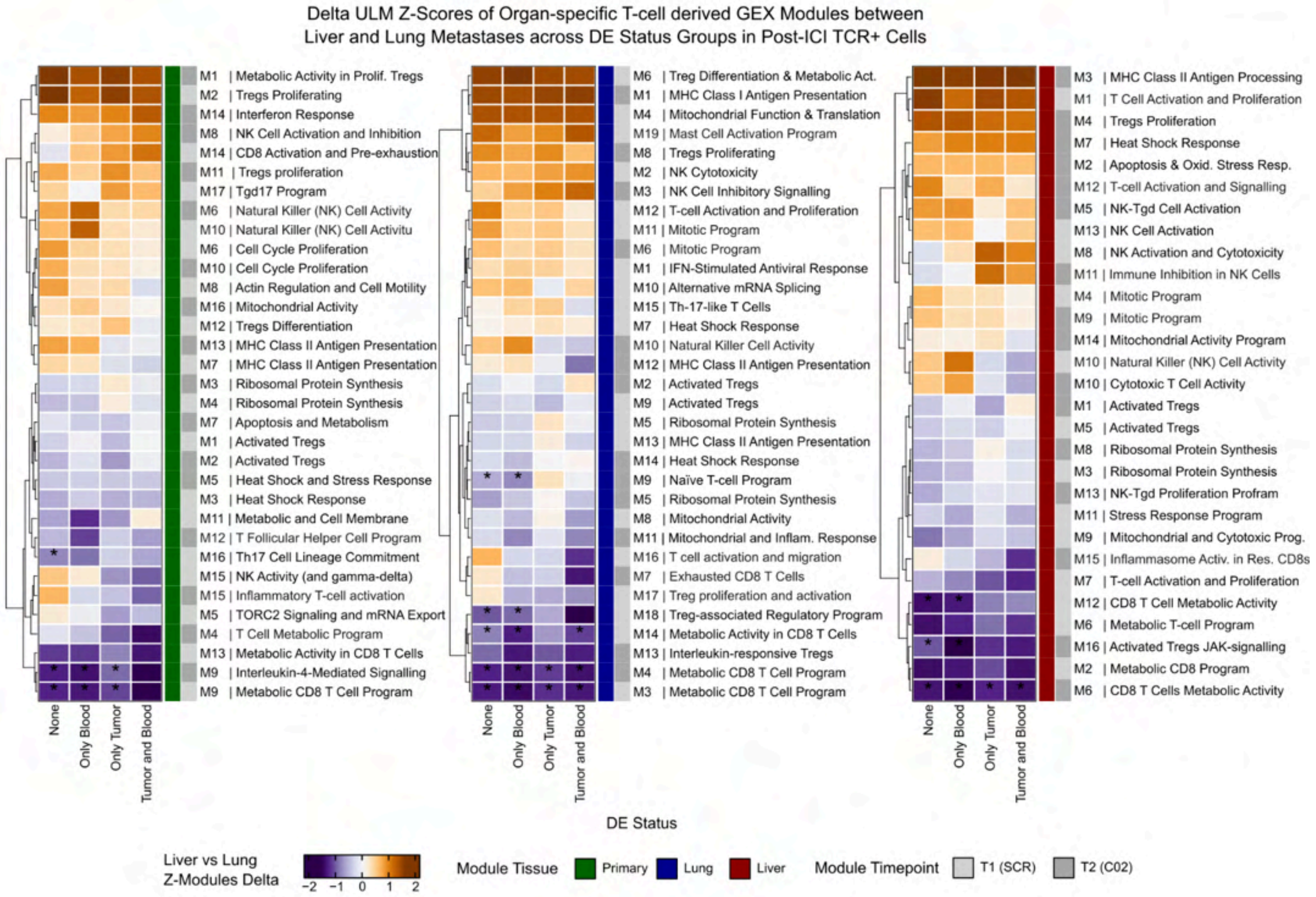


**Figure S34. Delta ULM Z-Scores of Organ-specific T-cell derived GEX Modules between liver and lung metastases tumor post-ICI TCR+ cells by differential expansion group**

Heatmaps showing the delta between liver and lung metastases of ULM Z-scaled scores of Organ-derived (Primary CRC in green, Lung mets in blue, Liver mets in red) and timepoint-specific (SCR in light gray, C02 in dark gray) GEX modules computed with hdWGCNA from the T-cell scRNA-seq compartment on the tumor post-ICI (C02) TRC+ cells split by DE Status groups. Positive Z-scores delta values shown in orange indicate higher expression of the module in liver metastases, while negative values shown in purple indicate higher expression in lung metastases. Modules and DE Status groups are hierarchically clustered based on their deltas across DE groups and modules, respectively. Modules are functionally annotated with *Llegir* (see **Methods**). Statistical significance of unpaired differences between metastatic sites was assessed through two-sided Wilcoxon rank-sum tests and corrected for multiple testing via the Bonferroni-Hochberg method. The significance threshold was set at 0.05 and significant differences for each module and tissue are shown with an asterisk (*).

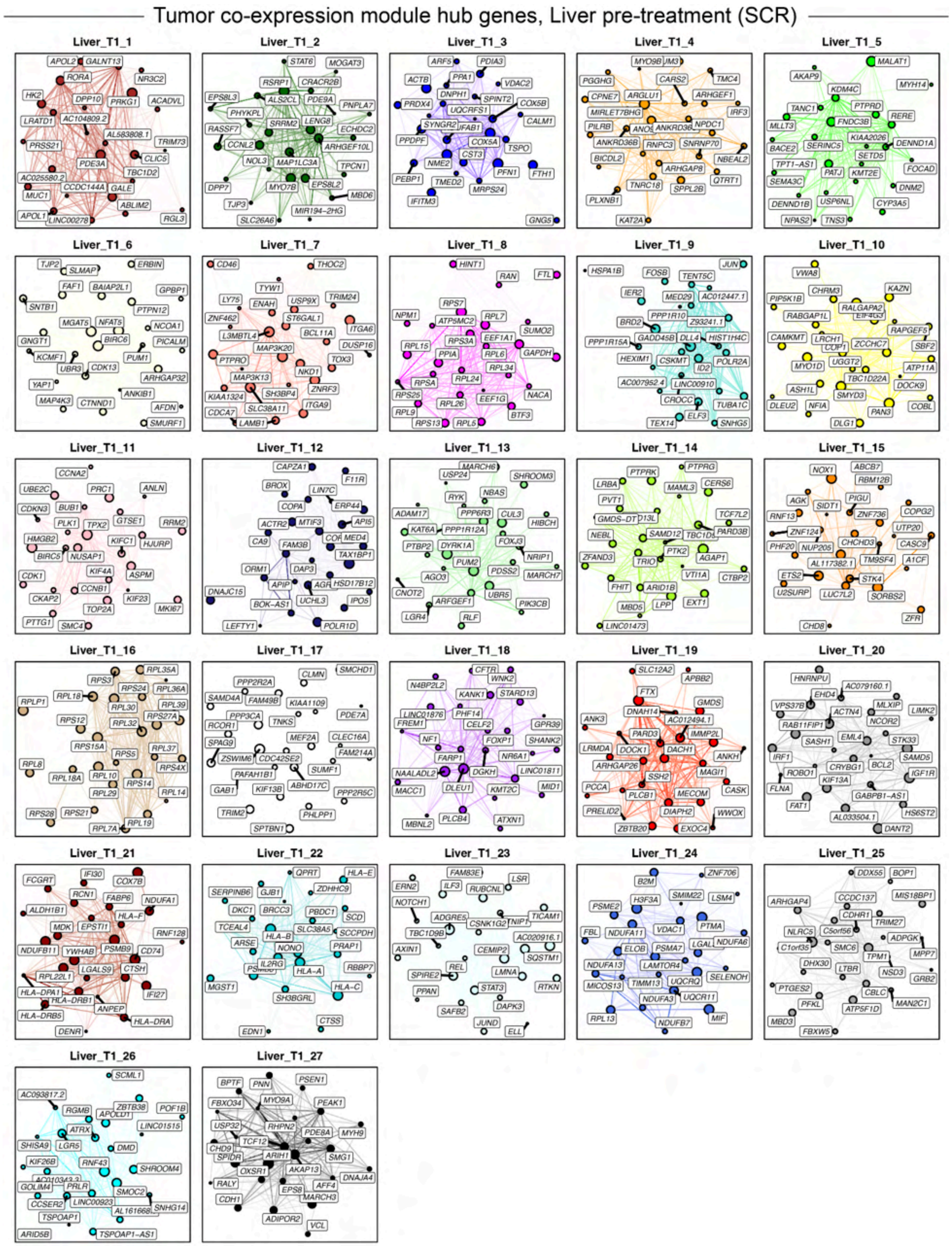


**Figure S35. Tumor gene co-expression module hub genes, liver pre-treatment**

Figure caption on the next page.

**Figure S35. Tumor gene co-expression module hub genes, liver pre-treatment**

Hub gene network for each of the 27 liver pre-treatment tumor gene co-expression modules identified via hdWGCNA[7]. Nodes represent genes, and edges represent co-expression links (topological overlap). The top 25 hub genes ranked by eigengene-based intramodular connectivity (kME) are shown, and the top 250 edges per module are shown.

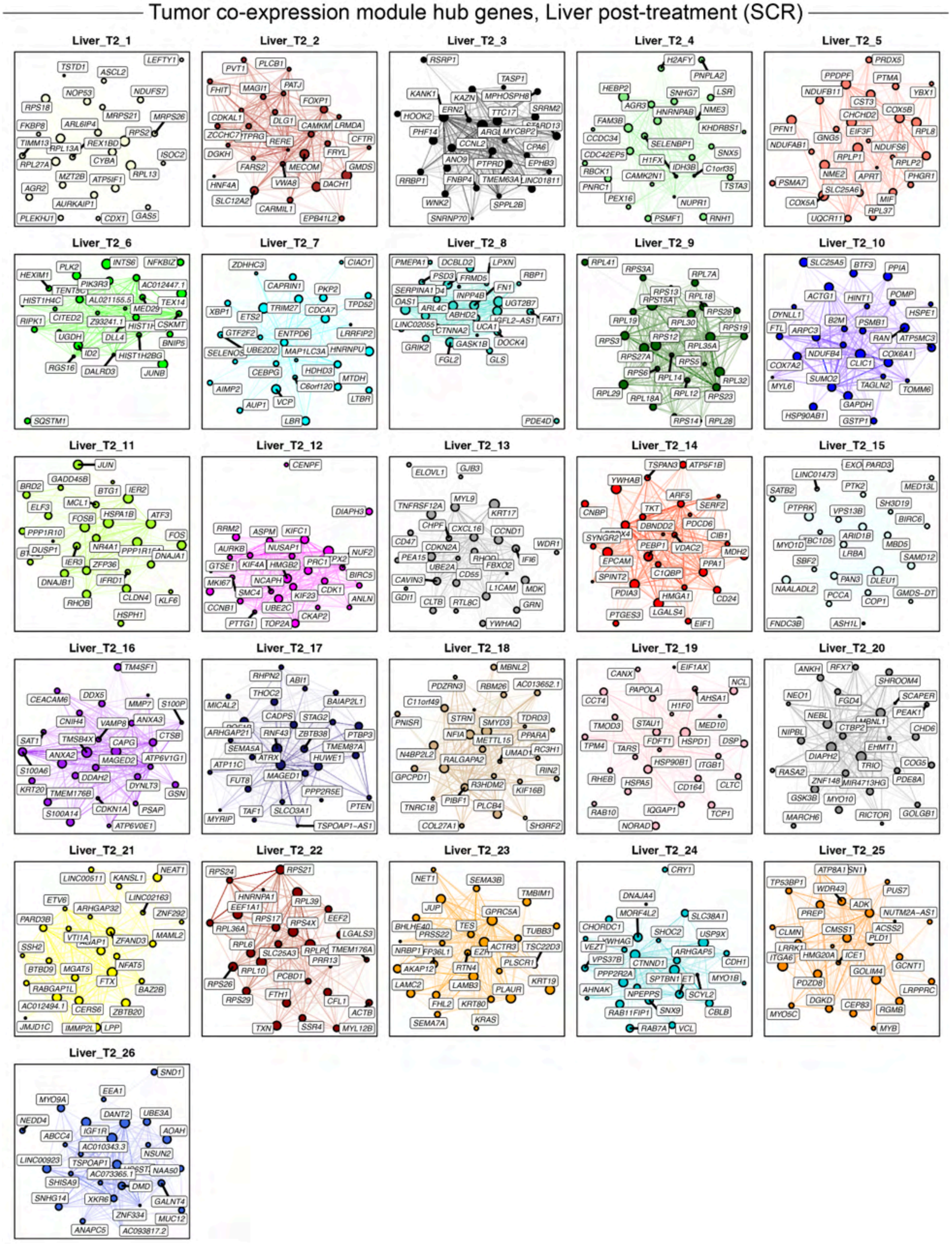


**Figure S36. Tumor gene co-expression module hub genes, liver post-treatment**

Figure caption on the next page.

**Figure S36. Tumor gene co-expression module hub genes, liver post-treatment**

Hub gene network for each of the 26 liver post-treatment tumor gene co-expression modules identified via hdWGCNA[7]. Nodes represent genes, and edges represent co-expression links (topological overlap). The top 25 hub genes ranked by eigengene-based intramodular connectivity (kME) are shown, and the top 250 edges per module are shown.

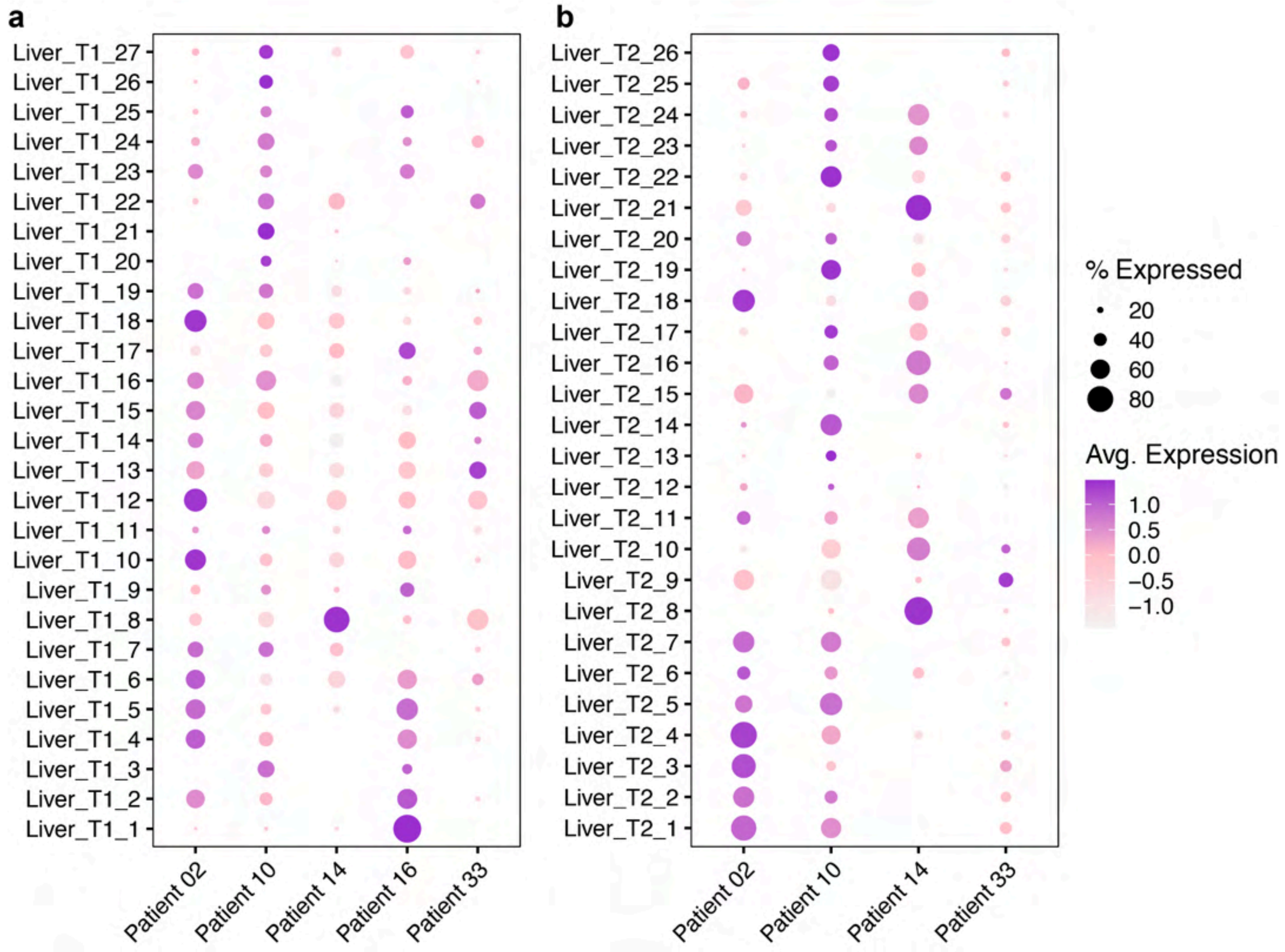


**Figure S37. Liver metastasis tumor compartment gene co-expression module expression levels**
Dot plots showing the single-cell gene-expression signatures (Module Eigengenes, MEs) for each of the co-expression modules pre-treatment (**a**) and post-treatment (**b**) in each patient.

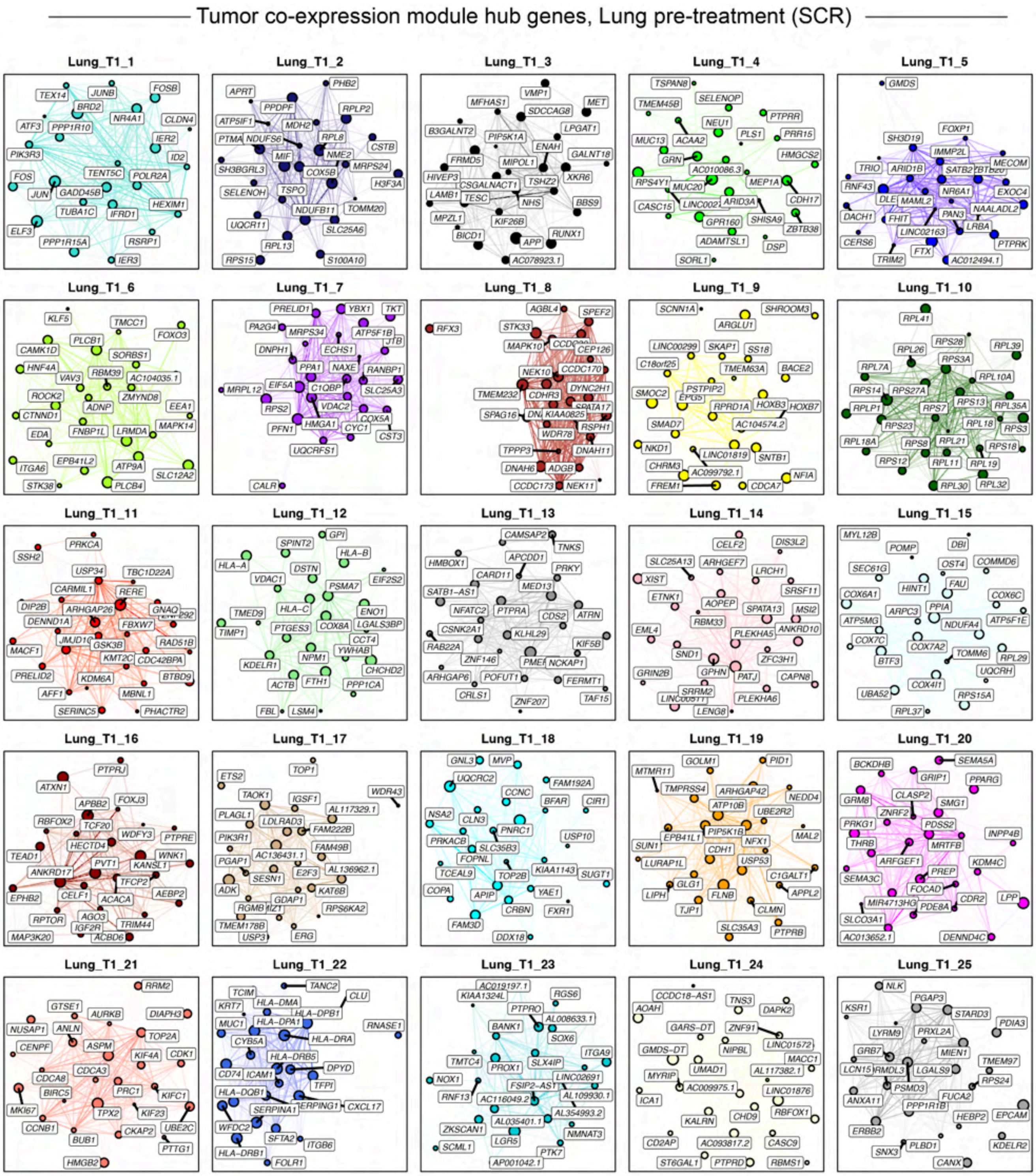


**Figure S38. Tumor gene co-expression module hub genes, lung pre-treatment**

Hub gene network for each of the 25 lung pre-treatment tumor gene co-expression modules identified via hdWGCNA[7]. Nodes represent genes, and edges represent co-expression links (topological overlap). The top 25 hub genes ranked by eigengene-based intramodular connectivity (kME) are shown, and the top 250 edges per module are shown.

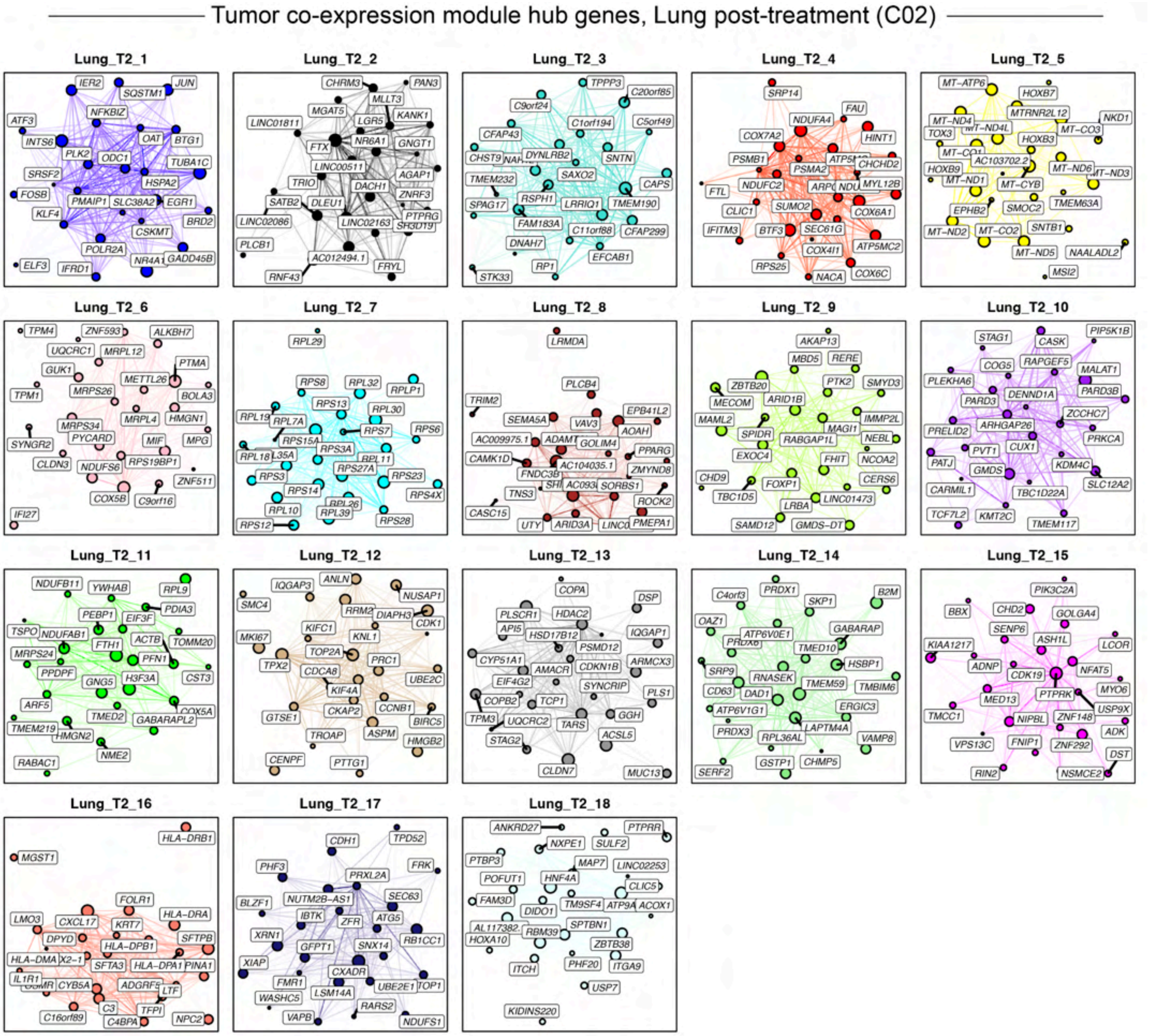


**Figure S39. Tumor gene co-expression module hub genes, lung post-treatment**

Hub gene network for each of the 18 lung post-treatment tumor gene co-expression modules identified via hdWGCNA[7]. Nodes represent genes, and edges represent co-expression links (topological overlap). The top 25 hub genes ranked by eigengene-based intramodular connectivity (kME) are shown, and the top 250 edges per module are shown.

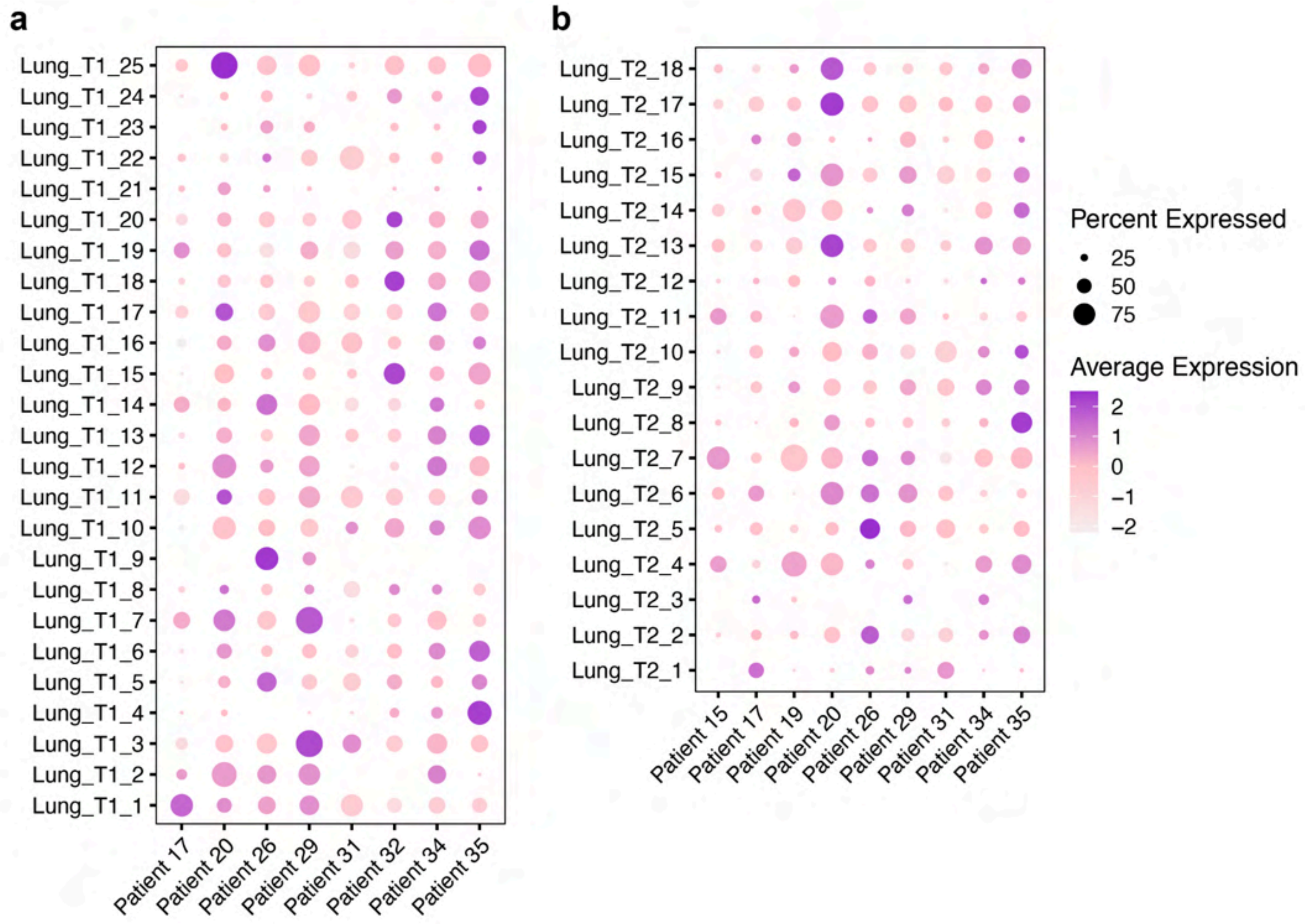


**Figure S40. Lung metastasis tumor compartment gene co-expression module expression levels**

Dot plots showing the single-cell gene-expression signatures (Module Eigengenes, MEs) for each of the co-expression modules pre-treatment (**a**) and post-treatment (**b**) in each patient.

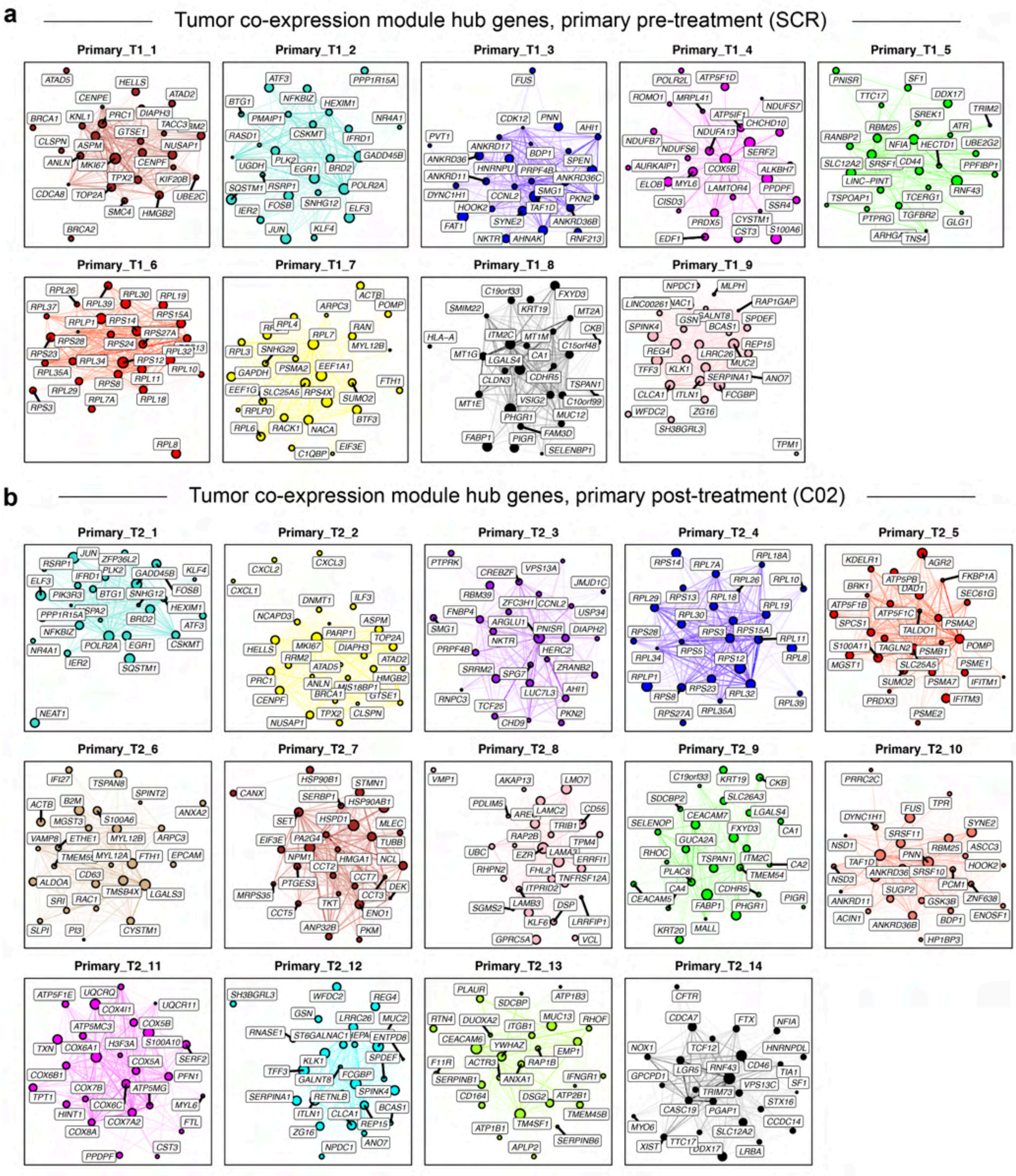


**Figure S41. Tumor gene co-expression module hub genes, lung post-treatment**

Hub gene network for each of the Primary tumor pre- (**a**) and post-treatment (**b**) gene co-expression modules identified via hdWGCNA[7]. Nodes represent genes, and edges represent co-expression links (topological overlap). The top 25 hub genes ranked by eigengene-based intramodular connectivity (kME) are shown, and the top 250 edges per module are shown.

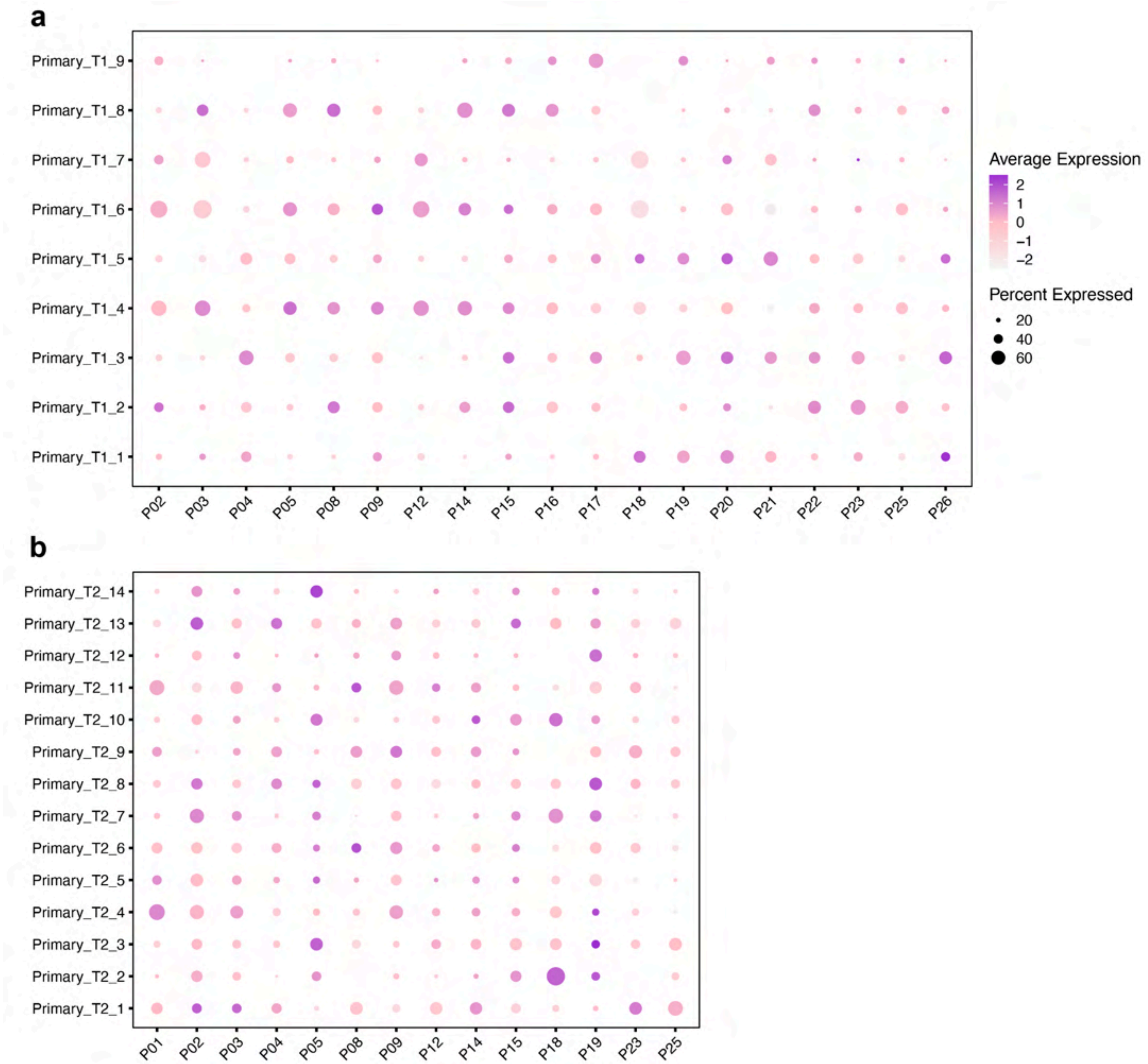


**Figure S42. Primary CRC tumor compartment gene co-expression module expression levels**

Dot plots showing the single-cell gene-expression signatures (Module Eigengenes, MEs) for each of the co-expression modules pre-treatment (**a**) and post-treatment (**b**) in each patient.

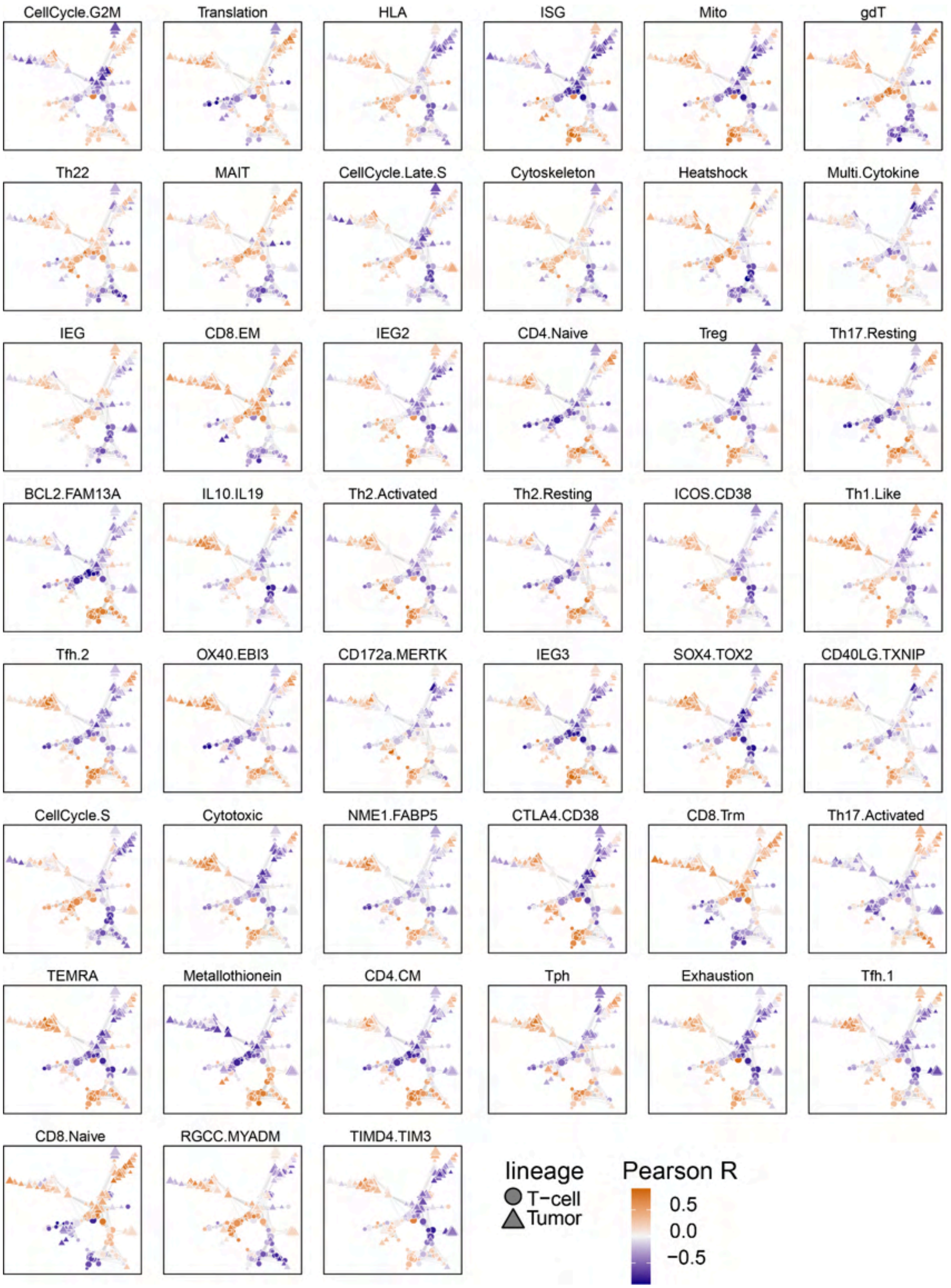


**Figure S43. Correlation of co-expression module dynamics and T-cell cGEPs**

Coordination network from panel **Fig. 5e**, colored by Pearson correlation between the treatment-induced dynamics of each module (Δ C02 - SCR) and that of the each TCAT consensus gene expression program (cGEP[8]).

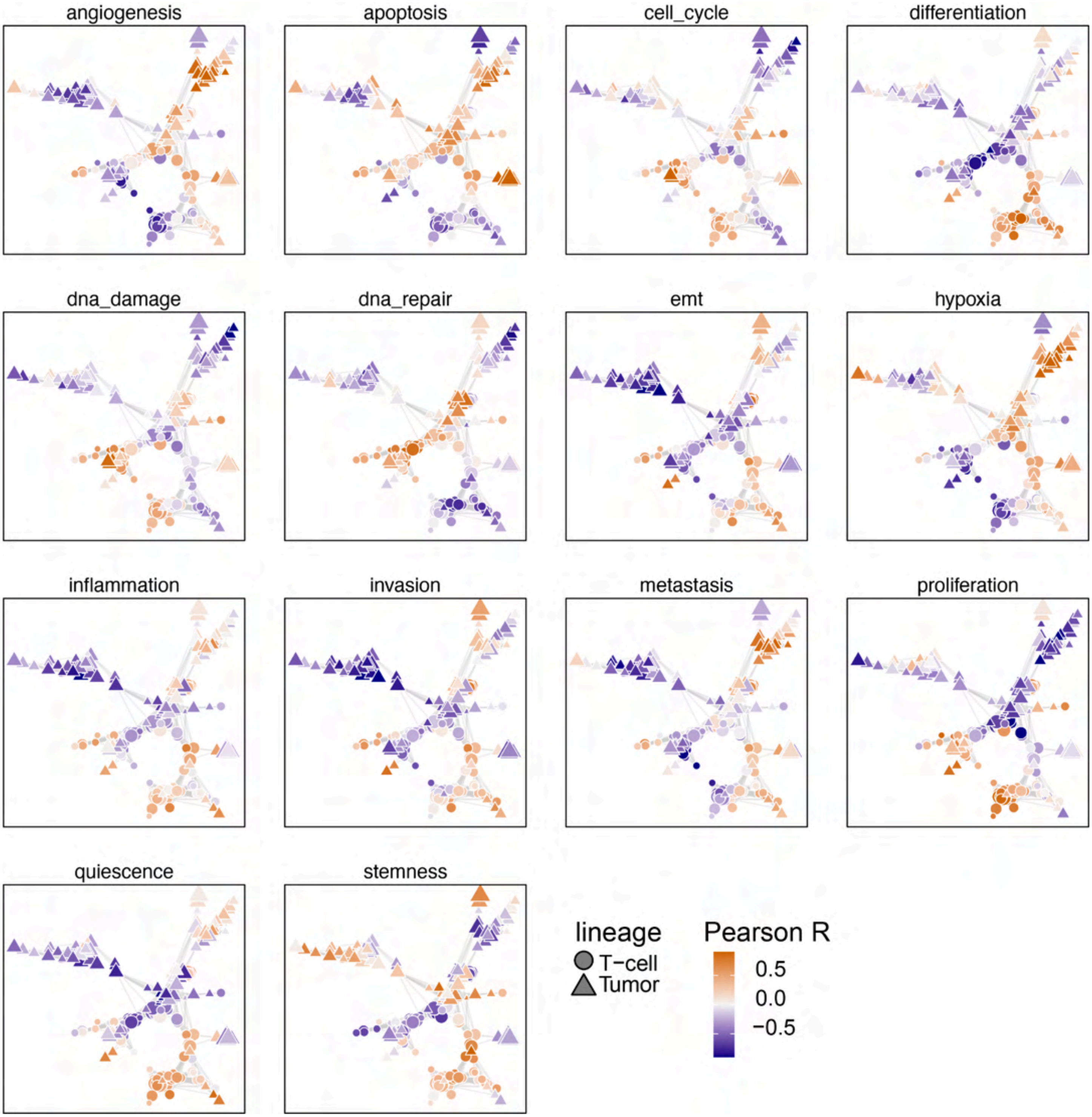


**Figure S44. Correlation of co-expression module dynamics and CancerSEA**

Coordination network from panel **Fig. 5e**, colored by Pearson correlation between the treatment-induced dynamics of each module (Δ C02 - SCR) and that of the 14 signatures from CancerSEA[9].
